# Transcriptomic and Machine-Learning Prioritization of OCEL1 in Low Bone Mineral Density With Multivariable cis-MR

**DOI:** 10.64898/2026.09.16.26363200

**Authors:** Jiaheng Yao, Qiong Zhong

**Affiliations:** School of Physical Science and Engineering, College of Life Sciences and Bioengineering, Beijing Jiaotong University, Beijing 100044, China; Beijing Weiguang Genome Technology Co., Ltd., Beijing, China

**Author notes:** **Corresponding author Correspondence Jiaheng Yao** School of Physical Science and Engineering, College of Life Sciences and Bioengineering, Beijing Jiaotong University, Beijing 100044, China.

**Keywords:** low bone mineral density, OCEL1, transcriptomics, machine learning, cis-Mendelian randomization

## Abstract

**Introduction:** Low bone mineral density (BMD) is associated with altered bone remodeling and osteoimmune regulation. We aimed to prioritize transcript-level signals supported across human expression cohorts and evaluate their attribution within local cis-regulatory architecture.

**Materials and Methods:** Nominal differential-expression signals from GSE56815 were restricted with a predefined 25-term MSigDB thematic library. Features measurable in GSE56815 and GSE2208 underwent linear SVM ranking, LASSO, and XGBoost selection, followed by cross-cohort transcriptomic assessment. OCEL1 was examined in a four-exposure locus-aware CisMRBEEX model with NR2F6, MRPL34, BABAM1, eQTLGen cis-eQTLs, heel eBMD GWAS statistics, and UKBB337K LD.

**Results:** The expression screen identified 2,568 genes at nominal P<0.05; thematic restriction retained 190 candidates and 107 common features entered machine learning. Linear SVM, LASSO, and XGBoost retained 24, 18, and 9 genes and converged on CPNE1, EZR, and OCEL1. Only OCEL1 showed concordant expression direction with P<0.05 in both cohorts. In the primary 268-variant four-exposure model, OCEL1 was positively associated with heel eBMD (β=0.010293, SE=0.004107, 95% CI 0.002244-0.018342, P=0.01220, PIP=0.4001; conditional coefficient on the standardized analysis scale); BABAM1 retained an oppositely directed component. The no-palindromic reconstruction remained positive with wider uncertainty (β=0.007652, P=0.06739).

**Conclusion:** Convergent transcriptomic evidence prioritized OCEL1, whose conditional genetic-expression component was positively associated with heel eBMD after local shared cis regulation was modeled.

## Introduction

Osteoporosis is characterized by reduced bone mass and impaired bone strength, resulting in greater fragility-fracture risk [1]. Bone remodeling depends on the balance between osteoblast-mediated formation and osteoclast-mediated resorption, with immune signaling contributing to this balance [2]. Osteoclasts arise from monocyte-macrophage lineages, and nuclear factor-kappaB (NF-κB) signaling contributes to osteoclast differentiation, survival, and bone-resorptive activity [3,4]. Circulating monocyte-lineage transcriptomes therefore provide a human molecular perspective on osteoimmune processes relevant to BMD variation.

Public transcriptomic datasets can identify reproducible expression differences, but their dimensionality creates a candidate-prioritization problem. A permissive expression screen retains potential signals, whereas biologically constrained filtering and complementary machine-learning procedures apply distinct feature-prioritization criteria. Cross-cohort transcriptomic support can then determine whether convergent candidates retain consistent expression behavior outside the discovery cohort. In this study, machine learning provided a structured layer for integrating complementary feature-selection criteria.

Genetic follow-up poses a separate attribution challenge because cis-regulatory loci often influence multiple neighboring transcripts. Consequently, a single-exposure genetic association can reflect shared local regulation rather than a uniquely attributable transcript. Molecular-QTL prioritization frameworks emphasize local transcript attribution within correlated regulatory architecture [5], and multivariable cis-MR can estimate conditional transcript-level components while retaining correlated cis instruments and addressing weak-instrument structure [6,7]. OCEL1 is of interest in this setting because experimental work has identified human OCEL1 as a regulator of inflammatory responses linked to NF-κB signaling during bacterial infection [8], providing biological context for a potential link between OCEL1-associated inflammatory signaling and osteoimmune regulation.

We used a stage-wise analytic design in which transcriptomic discovery and cross-cohort support identified the focal gene, followed by locus-aware multivariable cis-MR of its genetic-expression component in relation to heel estimated BMD (heel eBMD). The analysis integrated GSE56815 discovery, GSE2208 cross-cohort transcriptomic support, eQTLGen blood cis-eQTL summary statistics, heel eBMD GWAS data, and UK Biobank-derived LD. The thematic candidate library preceded machine-learning selection; local co-exposures were defined exclusively from exposure-side cis architecture; and complementary sensitivity analyses assessed statistical-specification and exposure-set stability (Fig. 1).

**Fig. 1.**
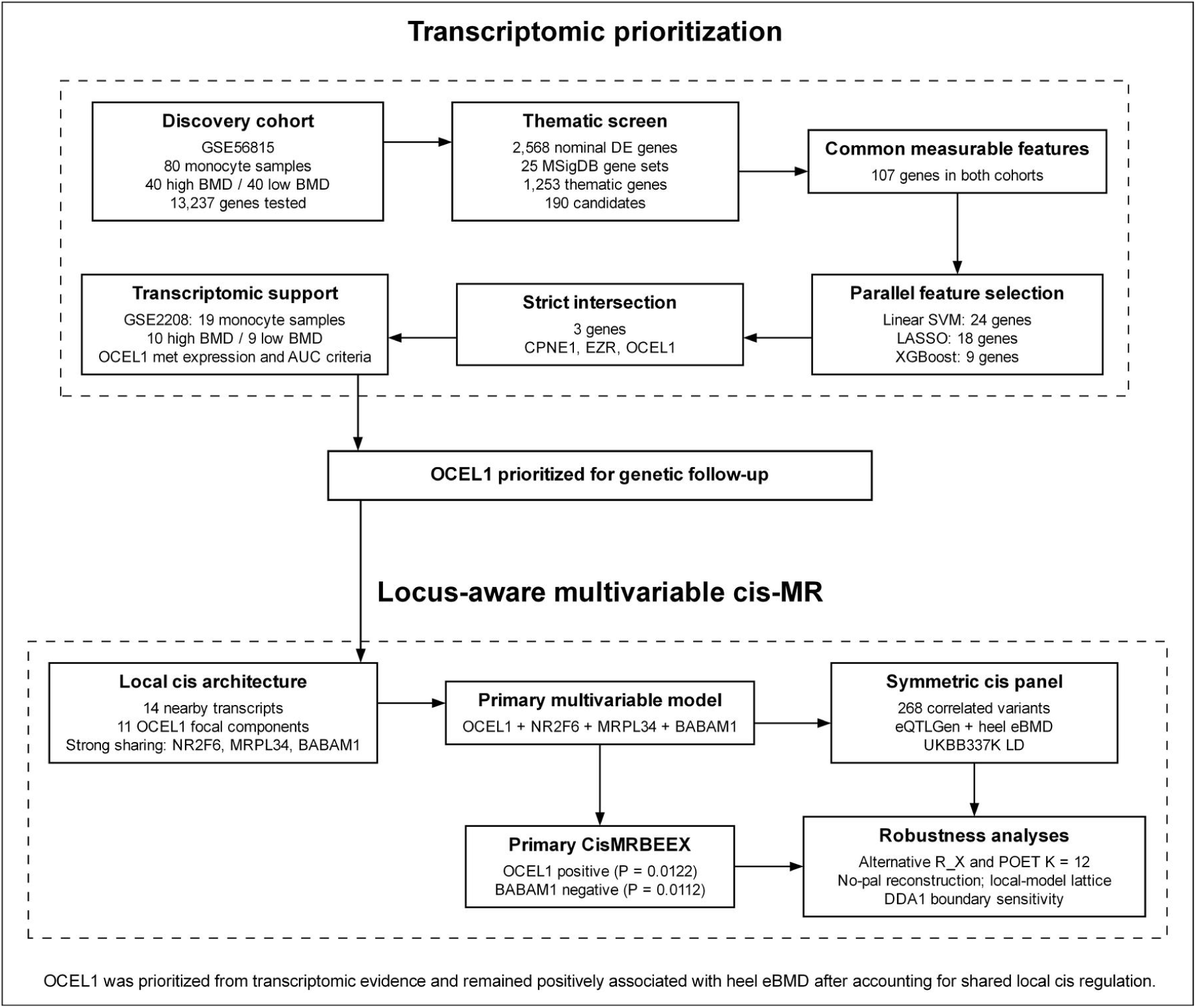
Overall study design and evidence flow. Figure 1 depicts the analytical sequence from GSE56815 transcriptomic discovery and thematic screening through common-feature definition across cohorts, parallel linear-SVM ranking, LASSO and XGBoost selection, the strict three-gene intersection, transcriptomic support in GSE2208, and OCEL1 prioritization. The genetic module summarizes local cis-architecture analysis, construction of the symmetric 268-variant cis panel for the four-exposure model, primary CisMRBEEX estimates, and robustness analyses

Mendelian randomization uses inherited genetic variation as an instrumental-variable framework intended to estimate causal exposure-outcome effects under defined assumptions.

## Materials and Methods

### Study design and public data resources

The study used a stage-wise design: transcriptomic discovery and biological restriction established a common candidate space; three complementary machine-learning procedures identified convergent genes; an independent monocyte cohort provided cross-cohort transcriptomic support; and locus-aware multivariable cis-MR evaluated the focal genetic-expression signal in its local regulatory context. The focal transcript was selected before genetic outcome analysis. Primary and sensitivity analyses were reported separately in accordance with STROBE-MR reporting principles [9]. GSE56815 was the discovery cohort and comprised 80 female peripheral-blood monocyte samples selected from high- and low-hip-BMD extremes (40 high BMD and 40 low BMD), with premenopausal and postmenopausal participants balanced within each BMD group [10,11]. GSE2208 served as the independent transcriptomic support cohort and comprised 19 monocyte samples (10 high BMD and 9 low BMD) [10,12]. For both cohorts, expression values were obtained from the GEO Series Matrix and phenotype assignments were recovered from the corresponding family SOFT metadata. Probes with entirely missing expression were removed, and isolated missing values were imputed using the probe-wise median. Gene annotations were harmonized across cohorts before defining common measurable features. GPL96 annotation was used for probe-to-gene mapping; when multiple probes represented one gene, the probe with the highest mean expression was retained. This procedure yielded 13,237 gene-symbol rows in GSE56815 and 6,271 in GSE2208. Of the 190 thematic candidates, 107 were measurable in both cohorts and entered machine-learning analyses.

Genetic associations with blood transcript expression were obtained from eQTLGen Phase I cis-eQTL summary statistics [13]. This resource meta-analyzed up to 31,684 blood or peripheral-blood mononuclear-cell samples from 37 contributing datasets, including 25,482 whole-blood and 6,202 PBMC samples; most participants were of European ancestry. Association-specific sample sizes from eQTLGen were used in standardization of the xQTL statistics. Heel eBMD associations were taken from GWAS Catalog study GCST006979, based on 426,824 White British UK Biobank participants with valid heel quantitative-ultrasound measurements (233,185 women and 193,639 men) [14]. The source GWAS used BOLT-LMM and included age, sex, genotyping array, assessment center, and ancestry principal components 1-20 as fixed effects. We retained the adjustment models of the contributing studies. UKBB337K European-ancestry LD supported variant mapping, LD pruning, component-sharing calculations, and construction of local LD matrices. The exposure and outcome resources were considered sufficiently ancestry-aligned for two-sample analysis because eQTLGen was predominantly of European ancestry and the heel eBMD GWAS was restricted to White British participants; European-ancestry UKBB337K LD was used throughout the local genetic analysis. Exposure and heel eBMD summary associations were transformed to the standardized summary-statistic scale. For exposure associations, β*=Z/√N and SE*=1/√N were calculated using variant-transcript-specific eQTLGen sample sizes; heel eBMD associations were transformed analogously using N=426,824.

No a priori power or sample-size calculation was undertaken; the analyzed sample sizes were determined by the available public source datasets.

### Ethics and informed consent

Only publicly available, de-identified expression data and published summary statistics were analyzed. The original GSE56815 and GSE2208 studies reported institutional ethical approval and participant informed consent [11,12]. The contributing eQTLGen cohorts and the UK Biobank study likewise obtained study-specific ethical approval and participant informed consent as described in their source publications [13,14]. No participants were newly recruited, and no identifiable individual-level data were accessed.

### Thematic candidate library and differential-expression screening

A predefined 25-gene-set MSigDB thematic library represented biological domains relevant to BMD and osteoimmune regulation, including bone remodeling and osteoclastogenesis, vitamin D/VDR-related regulation, NF-κB signaling, monocyte differentiation and activation, and monocyte/macrophage state or perturbation signatures [15,16]. Online Resource 1, Table S1 provides the complete gene-set list and MSigDB collection provenance. The union comprised 1,253 unique genes. Differential expression in GSE56815 was assessed with limma using a two-group no-intercept design and a low-BMD minus high-BMD contrast [17]. Following the nominal differential-expression strategy used in the source-linked monocyte/BMD study, nominal P<0.05 defined the permissive first-stage screen before thematic intersection [11]. FDR-adjusted results were also calculated for descriptive reporting.

### Common measurable feature space and machine-learning selection

Intersecting the nominal differential-expression screen with the 1,253-gene thematic library yielded 190 candidates. Candidates measurable in both the GSE56815 and GSE2208 Series Matrices were retained for cross-cohort evaluation, yielding 107 common features. All three machine-learning procedures were trained exclusively in GSE56815.

Three complementary feature-selection procedures were applied in parallel. For linear SVM weight-based ranking, a linear SVM was fitted to the 107 discovery features with internal predictor scaling, and genes were ranked once by the absolute magnitude of the fitted linear-SVM weights. Fixed top-ranked subsets of 2-40 genes were evaluated by three repeats of 10-fold cross-validation. The maximum internal subset-selection accuracy was 0.9833 at both 24 and 26 features; the smaller tied subset of 24 genes was retained [18]. Logistic LASSO used 10-fold cross-validation with the lambda.1se rule and retained 18 nonzero genes [19]. XGBoost used a gbtree binary-classification model with 5-fold cross-validation and early stopping; occurrence frequency summarized feature importance, and selection extended through the first rank at which cumulative frequency reached or exceeded 0.50, yielding 9 genes [20–22]. The strict intersection of the three selected sets defined the convergent candidate set (Fig. 2; Online Resource 1, Table S2).

**Fig. 2.**
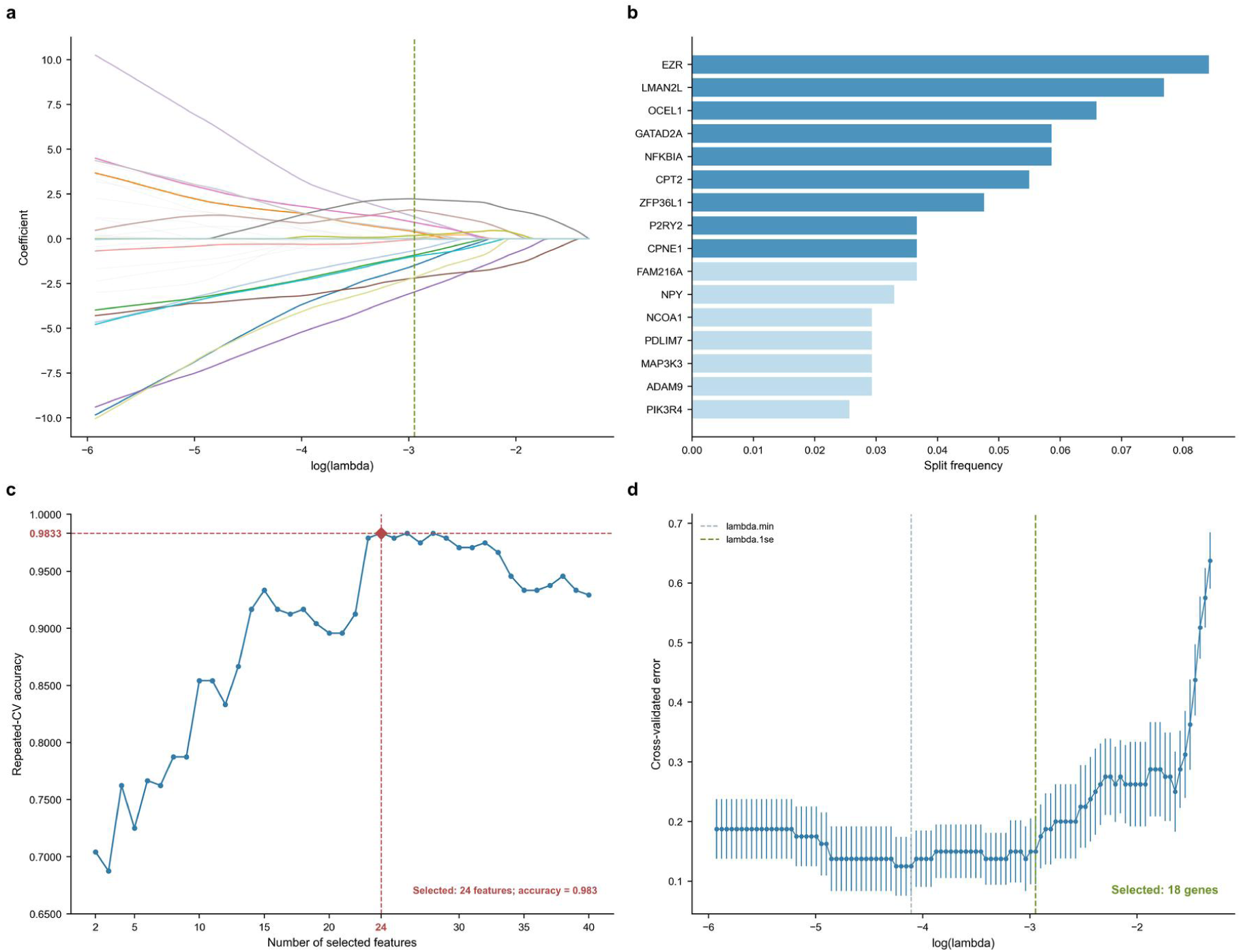
Machine-learning selection. (a) LASSO coefficient paths over the regularization sequence, with the selected lambda indicated. (b) XGBoost feature-occurrence importance and the cumulative-frequency threshold used for selection. (c) Performance of top-ranked linear-SVM subsets across candidate subset sizes; the 24-feature subset was the smaller solution tied for the highest repeated cross-validation accuracy. (d) LASSO cross-validation profile, showing lambda.min and the lambda.1se value used for feature selection

### Cross-cohort transcriptomic support

Genes in the strict three-model intersection were evaluated independently in GSE56815 and GSE2208. For each gene and cohort, we calculated the low-BMD minus high-BMD expression difference, a two-sided Wilcoxon P value, and a single-gene ROC/AUC using pROC [23]. Discrimination was summarized as max(AUC, 1-AUC), whereas expression direction was assessed from the signed group difference. AUC support required a value of at least 0.60 in both cohorts [24]. Expression support required nonzero group differences in the same direction and P<0.05 in both cohorts. Genes meeting both criteria were retained for genetic follow-up; cross-cohort metrics for the three genes are reported in Online Resource 1, Table S3.

### Local co-exposure definition for multivariable cis-MR

OCEL1 was selected as the focal transcript before the MR stage, and local co-exposures were defined exclusively from exposure-side cis architecture. Eleven independently selected OCEL1 cis components served as focal components. Candidate discovery considered non-OCEL1 chromosome 19 transcripts within ±1 Mb of OCEL1 with at least one focal OCEL1 component having an eQTLGen FDR<0.05 cis-eQTL record, yielding 14 candidate transcripts. For each candidate, the complete released eQTLGen cis set at gene-level FDR<0.05 was evaluated; in this resource, FDR<0.05 corresponds to marginal P<1.829×10⁻⁵ [25]. Variants were harmonized to UKBB337K LD, and independent components were obtained by formal conditional selection using cojopy 1.0.0 (conditional P cutoff 1×10⁻³; COJO collinearity cutoff r²=0.90) [26,27]. For each of the 11 OCEL1 focal components, the maximum UKBB337K LD r² with independently selected candidate components was calculated. Strong component sharing was defined as r²≥0.80 [5]. BABAM1, MRPL34, and NR2F6 each had two strong matches and formed the primary four-exposure model: OCEL1 + NR2F6 + MRPL34 + BABAM1 → heel eBMD (Fig. 4A; Online Resource 1, Table S4). DDA1 had the component-sharing value nearest the 0.80 boundary (maximum r²=0.799005) and was examined in a five-exposure boundary sensitivity analysis. This analysis repeated variant-panel construction, LD estimation, and estimation-error modeling with DDA1 included before refitting OCEL1, NR2F6, MRPL34, BABAM1, and DDA1.

### Instrumental-variable assumptions and assessment

The MR analyses were interpreted within the instrumental-variable framework. The classical IV assumptions are relevance, independence, and exclusion restriction: genetic variants must predict the modeled exposures, be independent of unmeasured determinants of the exposure-outcome relation, and affect the outcome through the modeled exposure pathways. In the present analysis, relevance was addressed through exposure-side association screening and xQTL fine-mapping prefits. CisMRBEEX explicitly models sparse direct genetic effects on the outcome, thereby accommodating horizontal pleiotropic effects rather than requiring every analyzed variant to satisfy a strict no-direct-effect formulation of exclusion restriction [6,7]. Joint modeling of OCEL1, NR2F6, MRPL34, and BABAM1 represented shared local cis regulation, and exposure-association estimation error was corrected under the sparse local genetic-architecture framework. The same model framework was retained across sensitivity analyses while the stated statistical or local exposure-set specification was varied.

### Symmetric cis-variant panel construction and harmonization

Variant selection was symmetric across the four primary exposures. For each candidate SNP, the minimum association P value across OCEL1, NR2F6, MRPL34, and BABAM1 was calculated; variants with Pmin<2×10⁻⁵ were eligible for panel construction. This panel rule was aligned to the eQTLGen FDR<0.05 marginal threshold within a minimum-across-exposures correlated-IV construction [7,25]. Variants were harmonized across exposures and outcome and restricted to heel minor-allele frequency ≥0.01 [28]. Exposure-side association ranking determined the variant panel before outcome harmonization. Retained palindromic variants underwent cross-resource allele-frequency concordance assessment following standard harmonization principles [29], using a prespecified absolute-difference criterion of ≤0.10. A separate sensitivity analysis reconstructed the panel after palindromic variants were excluded before LD pruning.

Eligible variants were ordered by four-exposure Pmin and underwent greedy local LD pruning: after a variant was retained, later variants within ±1 Mb with r²≥0.64 were excluded, following the correlated-instrument construction used by cis-MRBEE/CisMRBEEX [7]. After harmonization, MAF filtering, and LD pruning, the symmetric four-exposure procedure retained 268 correlated cis variants. The independently rebuilt no-palindromic panel contained 263 variants (Online Resource 1, Tables S5A-S6).

No imputation was applied to genetic summary statistics; variants with incomplete information required for harmonization or model fitting were excluded.

### Summary-statistic scaling, LD regularization, and estimation-error correlation

For each exposure, SNP-expression associations were transformed to the standardized summary-statistic scale β*=Z/√N and SE*=1/√N, preserving the original Z statistics, P values, and effect directions. The 268 × 268 UKBB337K LD correlation matrix was regularized using principal orthogonal complement thresholding (POET) [30]. Retained-factor number was selected by the maximum adjacent eigenvalue-gap ratio across candidate K values, and the regularization parameter was the smallest value on a 0.025-0.25 grid that produced a minimum eigenvalue >0.001. This procedure selected K=6 and λ=0.025. The maximum retained pairwise r² was 0.6394; POET increased the minimum eigenvalue from approximately -1.28×10⁻⁴ to 0.00938 and reduced the condition number from approximately 1.02×10⁵ to 1.40×10³ (Fig. 4D).

The eQTLGen exposure meta-analysis and the UK Biobank heel eBMD GWAS were obtained from distinct source resources. Their source publications did not report participant overlap, and the released summary data did not permit an exact participant-level overlap count. Exposure-outcome estimation-error correlations were therefore specified as zero (Online Resource 1, Tables S7A-S7B).

### CisMRBEEX estimation and sensitivity analyses

The primary four-exposure model was fitted with CisMRBEEX, a multivariable cis-MR framework for correlated cis instruments that explicitly addresses weak-instrument bias and horizontal pleiotropy [7]. The model jointly incorporated standardized exposure associations, heel eBMD associations, the POET-regularized LD matrix, and the estimation-error correlation matrix. Exposure-side xQTL prefits used two-stage SuSiE-RSS [31], with an initial model of up to 15 components followed by a refit based on the first-pass credible sets; 95% coverage was used throughout [7,31]. The primary model used a PIP retention threshold of 0.20, a reliability threshold of 0.75, an xQTL PIP threshold of 0.30 [7], and up to 15 xQTL components. All model fits converged under the specified settings. Full computational settings are provided with the reproducibility code in Online Resource 2.

The focal parameter was the conditional OCEL1 coefficient, specified before heel eBMD outcome modeling; its focal test used two-sided P<0.05. OCEL1 was the sole focal estimand in this analysis and its focal test was not multiplicity-adjusted. PIPs and conditional coefficients for BABAM1, NR2F6, and MRPL34 were interpreted as local transcript-attribution outputs of the same joint model rather than as separate confirmatory hypotheses. Robustness analyses evaluated alternative R_X specifications, POET fixed at K=12, a no-palindromic sensitivity analysis, local co-exposure sensitivity analyses, and a five-exposure sensitivity analysis including DDA1 (Online Resource 1, Tables S8-S10).

### Software and reproducibility

MR analyses were performed in R 4.3.2. Estimation-error covariance was calculated with MRBEE 1.0.0, CisMRBEEX fitting used MRBEEX 0.1.0, SuSiE-RSS prefitting used susieR 0.14.2, and local conditional-component selection used cojopy 1.0.0. Exact computational settings, R session information, and derived MR outputs are provided in Online Resource 2.

The study protocol and analysis plan were not preregistered.

### Use of artificial intelligence in manuscript preparation

OpenAI GPT-5.6 was used for English-language polishing and to assist with selected analysis and visualization code. All AI-assisted outputs were reviewed and validated by the authors, who take full responsibility for the final content and analyses.

## Data Availability

Analysis code and derived result tables supporting this study are provided in Online Resource 2. GSE56815 and GSE2208 are publicly available through the NCBI Gene Expression Omnibus. eQTL summary statistics were obtained from eQTLGen, heel eBMD summary statistics from GWAS Catalog study GCST006979, and LD information from the UKBB337K European-ancestry reference. Original eQTL, GWAS, and LD resources remain subject to the access and redistribution terms of their respective providers.

## Supplementary Materials

Online Resource 1. Supplementary Methods, Supplementary Results, and Tables S1-S10 supporting the transcriptomic, machine-learning, local cis-architecture, and multivariable cis-MR analyses.

Online Resource 2. Analysis and plotting code, derived result tables, analysis specifications, and reproducibility documentation supporting the reported analyses.

## Results

### Stage-wise prioritization narrowed the discovery space to 107 common measurable features

The GSE56815 discovery matrix contained 13,237 genes, of which 2,568 had nominal differential-expression evidence at P<0.05. Intersection with the 1,253-gene thematic library retained 190 candidates. Cross-cohort measurability in GSE56815 and the GSE2208 Series Matrix retained 107 genes, which defined the machine-learning feature universe (Fig. 1).

### Three machine-learning models converged on three genes

Linear SVM weight ranking retained 24 genes under the smallest-tie rule: internal subset-selection accuracy reached 0.9833 at 24 and 26 features, and the 24-feature set was selected. LASSO at lambda.1se retained 18 genes, and XGBoost retained 9 genes at the first cumulative feature-frequency threshold of at least 0.50 (Fig. 2). The strict intersection of the three selected sets comprised CPNE1, EZR, and OCEL1 (Fig. 3A).

**Fig. 3.**
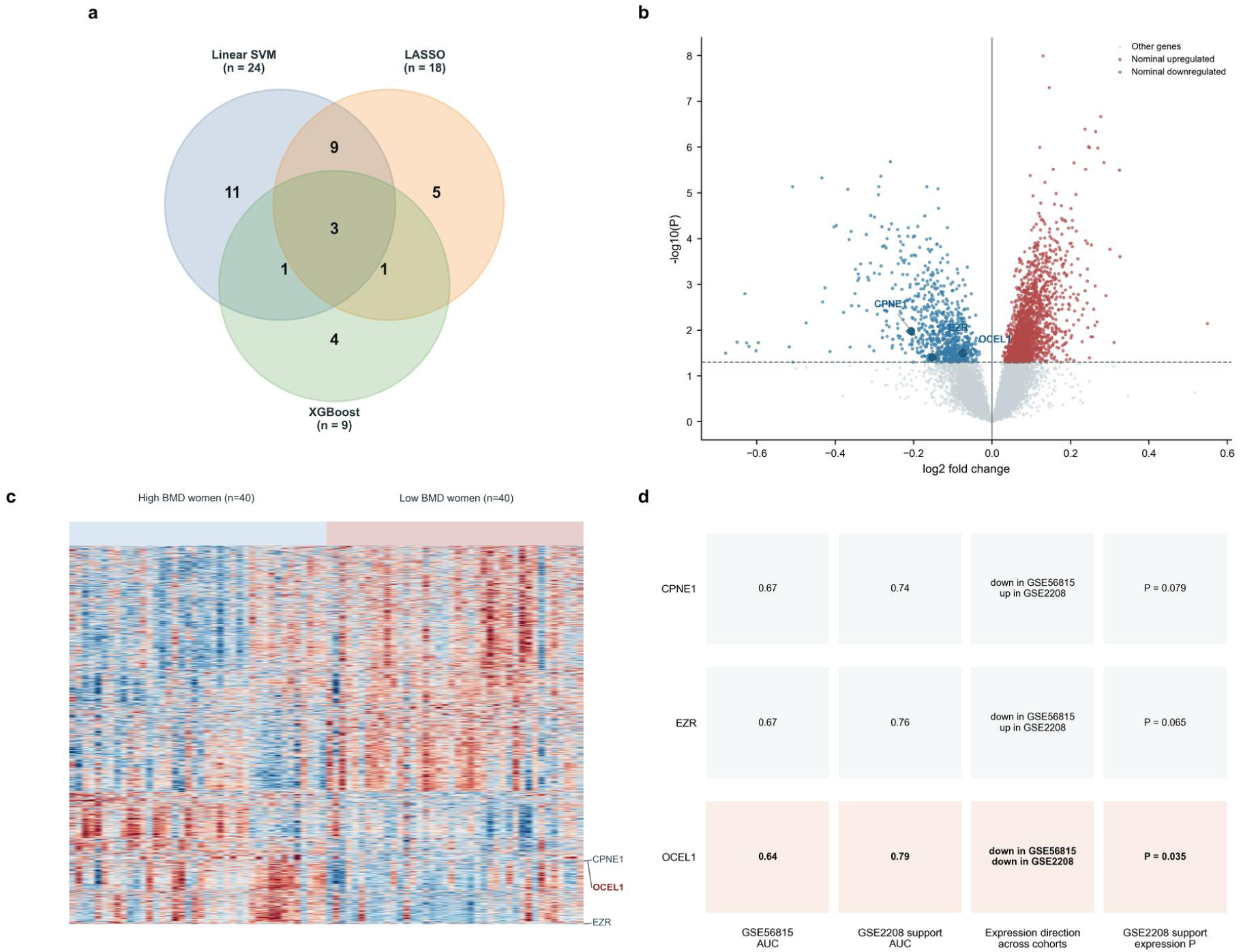
Machine-learning convergence and cross-cohort transcriptomic support. (a) Overlap among the linear-SVM ranking, LASSO, and XGBoost selections, identifying the strict three-gene intersection. (b) Differential-expression landscape in the discovery cohort, with CPNE1, EZR, and OCEL1 labelled. (c) Heatmap of the 3,000 genes with the smallest nominal limma P values; rows show gene-wise z scores across samples, with the three convergent genes indicated. (d) Cross-cohort transcriptomic support for the three overlapping genes, summarizing direction-agnostic AUC magnitude, expression direction, and expression P-value evidence across cohorts; OCEL1 alone met the combined support criteria

### Cross-cohort expression support prioritized OCEL1

OCEL1 was the only convergent gene with concordant cross-cohort expression support. Expression was lower in the low-BMD group in both GSE56815 (Δ=-0.0751, P=0.0376) and GSE2208 (Δ=-0.1289, P=0.0350), with direction-agnostic discrimination magnitudes of 0.635 and 0.7889, respectively. OCEL1 alone met the combined cross-cohort expression and AUC criteria and was taken forward as the focal genetic exposure (Fig. 3D; Online Resource 1, Table S3).

### Local cis architecture defined a four-exposure correlated-instrument model

BABAM1, MRPL34, and NR2F6 each had two strong component-sharing matches to the 11 OCEL1 focal components at r²≥0.80. DDA1 was the transcript nearest the boundary, with maximum r²=0.799005 (Fig. 4A). The primary correlated-instrument model therefore jointly represented OCEL1, NR2F6, MRPL34, and BABAM1.

**Fig. 4.**
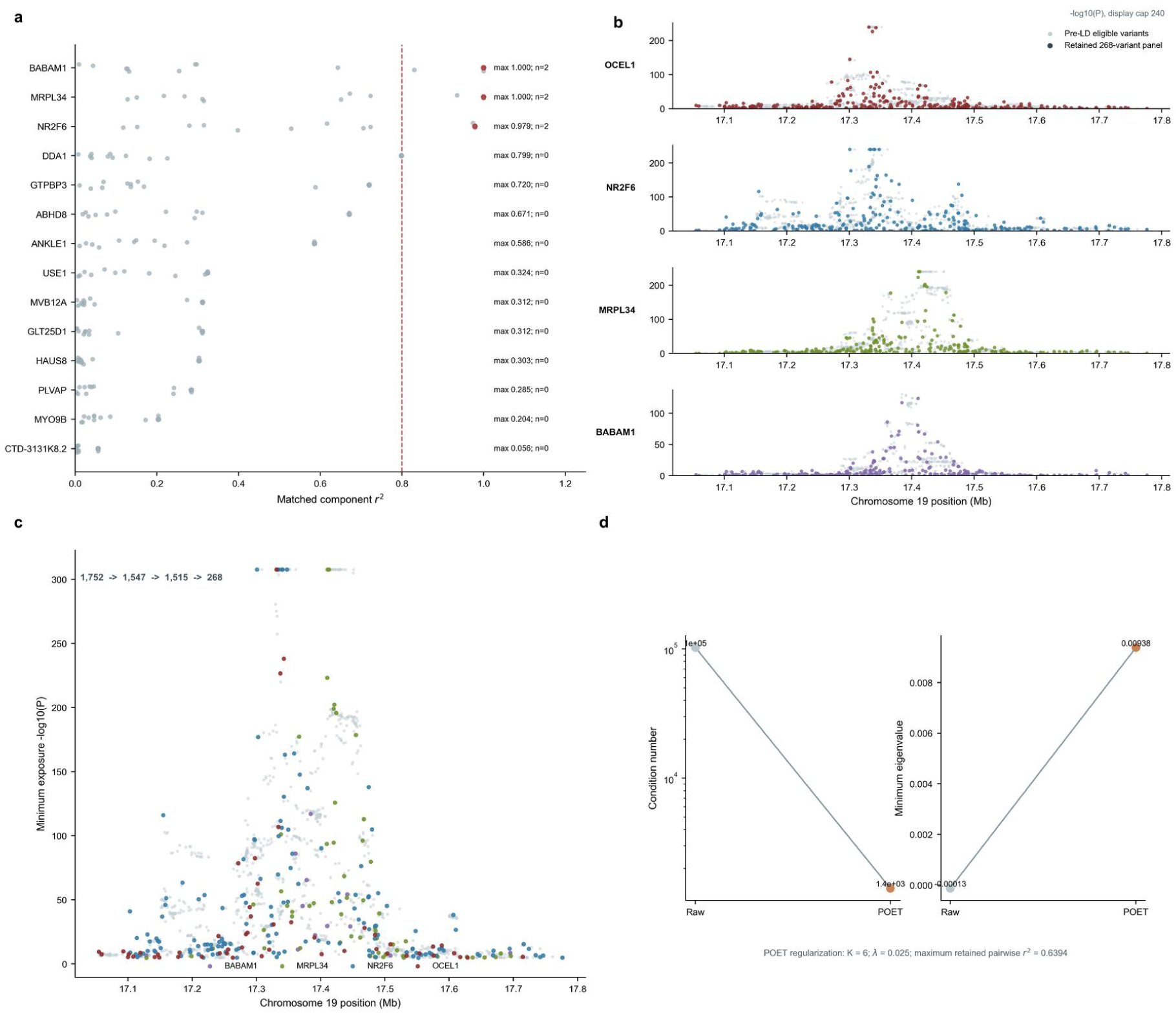
Local cis architecture and instrument construction. (a) Component sharing between 11 focal OCEL1 cis components and 14 neighboring transcripts. BABAM1, MRPL34, and NR2F6 each showed two matches at r²≥0.80, whereas DDA1 was the nearest transcript below this boundary. (b) Regional cis-eQTL tracks for OCEL1, NR2F6, MRPL34, and BABAM1, showing the eligible local-variant landscape and retained panel variants. (c) Symmetric construction of the four-exposure instrument panel, resulting in 268 correlated cis variants. (d) Diagnostics for the raw and POET-regularized LD matrices, including changes in the minimum eigenvalue and matrix conditioning

After harmonization, MAF filtering, and LD pruning, symmetric panel construction retained 268 correlated cis variants (Fig. 4B-C; Online Resource 1, Table S5A). The minimum exposure-side P value was driven by OCEL1 for 51 retained variants, NR2F6 for 148, MRPL34 for 57, and BABAM1 for 12. Maximum retained pairwise r² was 0.6394. POET selected K=6 and λ=0.025 and improved conditioning of the local LD matrix (Fig. 4D).

### Primary multivariable cis-MR retained opposing OCEL1 and BABAM1 components

In the primary 268-variant four-exposure CisMRBEEX model, OCEL1 retained a positive conditional association with heel eBMD (β=0.010293, SE=0.004107, 95% CI 0.002244-0.018342, P=0.01220, PIP=0.4001; Fig. 5A-B). BABAM1 retained an oppositely directed conditional association (β=-0.014290, SE=0.005631, 95% CI - 0.025327 to -0.003252, P=0.01117, PIP=0.4125). NR2F6 and MRPL34 had PIPs of 0.0930 and 0.0945, respectively, below the fixed 0.20 retention threshold, and were not retained by the sparse primary model. Thus, the joint model retained two local transcript-level components with opposing directions.

**Fig. 5.**
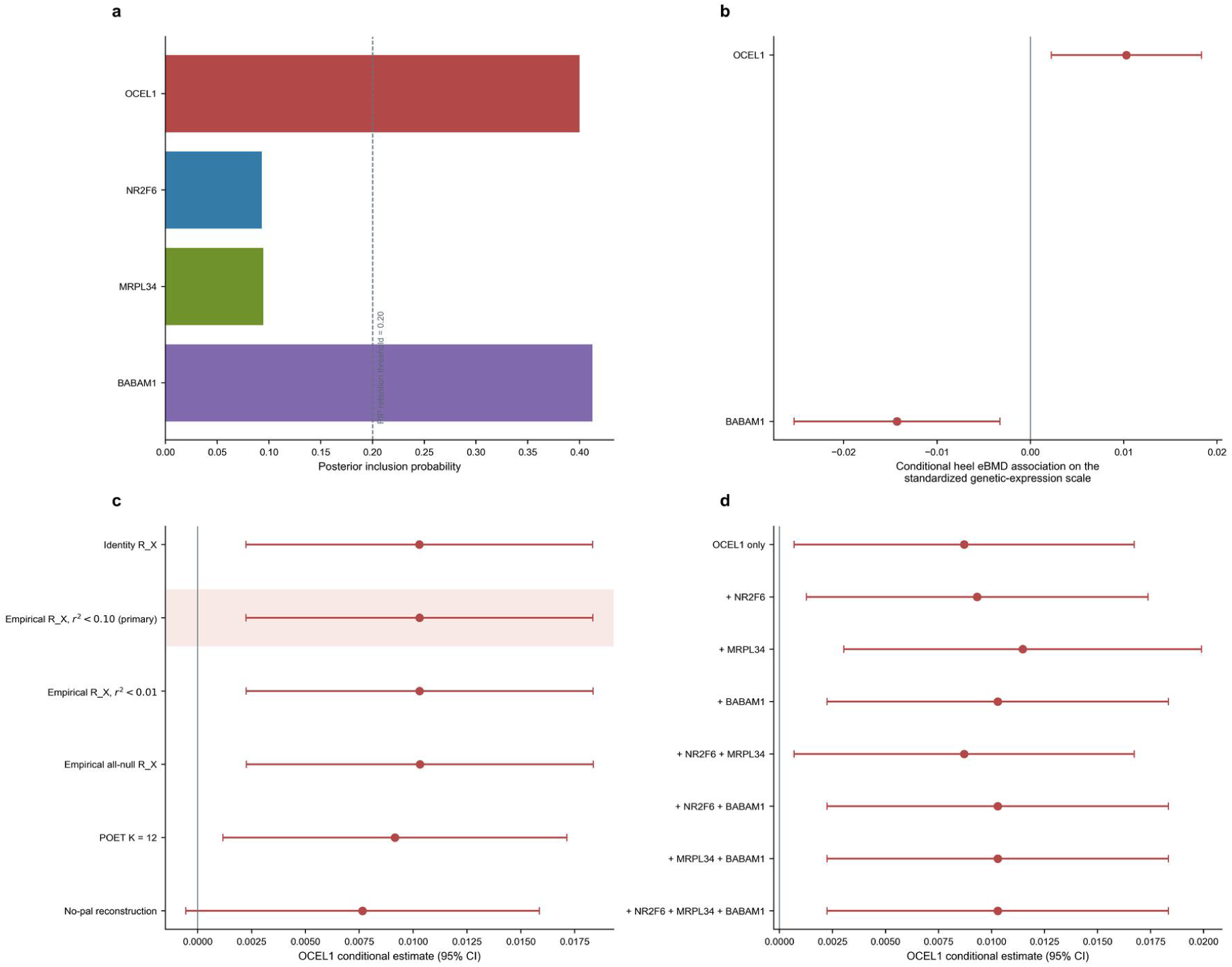
Primary multivariable cis-MR results and robustness. (a) Posterior inclusion probabilities (PIP) for the four primary exposures; the retention threshold was 0.20. (b) Conditional heel eBMD associations from the primary sparse model for retained exposures OCEL1 and BABAM1. (c) OCEL1 conditional estimates across alternative R_X specifications, POET K=12, and an independently reconstructed no-palindromic analysis. (d) OCEL1 estimates across local co-exposure analyses that varied included local co-exposures while retaining OCEL1 as the focal exposure

### The OCEL1 estimate was stable across statistical and local-model specifications

The OCEL1 estimate changed little across estimation-error specifications. Identity, all-null empirical, primary r²<0.10 empirical, and r²<0.01 empirical R_X definitions produced beta values of 0.010283, 0.010312, 0.010293, and 0.010300, respectively, with P values from 0.01208 to 0.01226 (Fig. 5C). Fixing POET at K=12 gave β=0.009155 (P=0.02465). In the independently reconstructed 263-variant no-palindromic analysis, the OCEL1 coefficient remained positive (β=0.007652, 95% CI - 0.000548 to 0.015852, P=0.06739), with wider uncertainty than in the primary analysis. Across eight local co-exposure specifications, the OCEL1 coefficient was positive, ranging from β=0.008710 to β=0.011469 (Fig. 5D). A fully reconstructed five-exposure analysis including DDA1 also produced an essentially unchanged focal estimate (four-exposure β=0.010293; five-exposure β=0.010298). DDA1 was not retained by the sparse five-exposure model (PIP=0.1112). Across most specifications, OCEL1 showed a consistent direction and similar effect magnitude.

## Discussion

This staged transcriptomic-to-genetic framework prioritized OCEL1 in low BMD while representing the local cis-regulatory context of the locus. Transcriptomic analysis narrowed 13,237 discovery genes to a biologically constrained 190-gene candidate set, 107 common measurable features, a three-gene intersection across three machine-learning procedures, and one focal transcript with concordant expression support in an independent monocyte cohort. Genetic follow-up indicated that the OCEL1 expression component remained positively associated with heel eBMD when modeled jointly with three strongly co-regulated neighboring transcripts. BABAM1 contributed an oppositely directed retained component at the same locus, supporting a multi-transcript representation of the regional genetic signal.

The transcriptomic and genetic results were directionally concordant. OCEL1 expression was lower in low-BMD monocytes in both transcriptomic cohorts, whereas the conditional genetic-expression coefficient for OCEL1 was positive for heel eBMD. Thus, lower measured OCEL1 expression accompanied lower BMD in the bulk datasets, while a higher genetically indexed OCEL1 expression component was associated with higher heel eBMD. Monocyte-lineage cells provide the precursor pool for osteoclastogenesis, and NF-κB signaling regulates osteoclast differentiation and resorptive activity [3,4]. Experimental work has also implicated OCEL1 in human inflammatory signaling, including NF-κB-related responses to bacterial infection [8]. These observations provide biological context for the convergent transcriptomic and genetic prioritization of OCEL1.

The locus-aware cis-MR analysis provides a distinct evidentiary layer because several transcripts at the OCEL1 locus share strong cis-regulatory components. Under this architecture, a single-exposure association can attribute a locus-level signal too specifically to one transcript. The joint model estimated the OCEL1 conditional component while incorporating NR2F6, MRPL34, and BABAM1, and retained a positive OCEL1 association with heel eBMD. Simultaneous retention of a negative BABAM1 component indicates that the joint model represented at least two transcript-level components with opposing associations at the locus. At the fixed PIP threshold, sparse-model support was concentrated on OCEL1 and BABAM1.

The focal OCEL1 estimate was stable across several statistical specifications. Alternative estimation-error covariance definitions produced nearly identical estimates, a larger POET factor setting preserved the positive association, all eight local co-exposure specifications retained a positive OCEL1 coefficient, and complete five-exposure reconstruction with DDA1 produced virtually no change in the focal estimate. The independently rebuilt no-palindromic analysis also retained a positive OCEL1 coefficient (β=0.007652, 95% CI -0.000548 to 0.015852, P=0.06739), although with wider uncertainty than the primary model. Across most sensitivity analyses, OCEL1 showed stable direction and similar effect magnitude, with lower precision in the no-palindromic reconstruction.

Several design features support interpretation of the analysis. All machine-learning procedures were trained in GSE56815; the focal transcript was selected before MR; local co-exposures were defined from exposure-side component sharing; and the correlated cis panel was built symmetrically across all four exposures from exposure-side association evidence. The genetic analysis directly modeled local LD, regularized the LD matrix, and estimated exposure-side error correlation from null cis variants.

Several features define the scope of the present evidence. Transcriptomic support was obtained from two relatively small peripheral-monocyte cohorts composed of women. The transcriptomic stage measured peripheral monocytes in BMD-defined cohorts, whereas the genetic stage used predominantly European-ancestry blood/PBMC cis-eQTL data and White British heel eBMD GWAS statistics; generalizability across sex, ancestry, cellular context, and BMD measurement context remains to be established. As with summary-data MR more generally, independence cannot be directly verified from the released data, and correlated pleiotropy or unmodeled local regulatory pathways may influence transcript attribution. CisMRBEEX models a sparse horizontal-pleiotropic direct-effect component and weak-instrument estimation error, while inference remains conditional on its sparse local-genetic-architecture framework. Exact participant overlap between eQTLGen and UK Biobank source studies was unavailable, so exposure-outcome estimation-error correlation was specified as zero. Equivalence between genetically indexed differences in blood transcript expression and experimental or therapeutic modulation of OCEL1 has not been established; the genetic coefficient therefore does not directly quantify an intervention effect. The independently rebuilt no-palindromic analysis remained directionally positive (beta=0.007652, 95% CI - 0.000548 to 0.015852, P=0.06739) but was less precise than the primary model. Overall, the data support cross-cohort transcript prioritization and locus-aware local genetic attribution of OCEL1 in relation to heel eBMD.

## Conclusion

A staged transcriptomic and locus-aware genetic analysis prioritized OCEL1 as a low-BMD-associated transcript. OCEL1 was the only member of the three-gene machine-learning intersection with concordant cross-cohort expression support, showing lower expression in the low-BMD groups of both cohorts. Its positive conditional association with heel eBMD persisted after joint modeling of the strongest locally shared cis transcripts. The opposing BABAM1 component illustrates the complexity of this locus and the value of multivariable local attribution. Together, these findings link cross-cohort transcriptomic prioritization to locus-aware local genetic attribution of OCEL1 within a multi-transcript cis-regulatory locus.

## Supporting information

Online Resource 1: Supplementary Methods, Results, and Tables S1-S10

Online Resource 2: Reproducibility Materials

## Author contributions

JY: Conceptualization, methodology, software, validation, formal analysis, investigation, data curation, visualization, project administration, writing - original draft, and writing - review and editing. QZ: Conceptualization, funding acquisition, resources, and writing - review and editing. Both authors reviewed and approved the final manuscript.

## Acknowledgments

This study received internal research funding from Beijing Weiguang Genome Technology Co., Ltd. Qiong Zhong holds an ownership interest in the company and, as an author, contributed to conceptualization, funding acquisition, resources, and writing - review and editing.

