## Supplementary material for "Transcriptomic and Machine-Learning Prioritization of OCEL1 in Low Bone Mineral Density With Multivariable cis-MR": Online Resource 1: Supplementary Methods, Results, and Tables S1-S10

Supplementary Methods, Results, and Tables for: Transcriptomic and Machine-Learning Prioritization of OCEL1 in Low Bone Mineral Density With Multivariable cis-MR

References cited by number correspond to the main manuscript reference list.

Authors: Jiaheng Yao1*, Qiong Zhong2

1 School of Physical Science and Engineering, College of Life Sciences and Bioengineering, Beijing Jiaotong University, Beijing 100044, China

2 Beijing Weiguang Genome Technology Co., Ltd., Beijing, China

#### Supplementary Methods

##### S1. Transcriptomic preprocessing and differential-expression analysis

Expression values for both cohorts were obtained from GEO Series Matrix files, and phenotype assignments from the corresponding family SOFT metadata. Probes with all values missing were removed, whereas isolated missing values were imputed using the probe-wise median. Expression scale was checked before gene-level mapping. Both datasets were processed with the same GPL96 annotation and probe-to-gene rule: for multiply annotated probes, the first symbol before the GPL96 separator was retained; when multiple probes mapped to the same symbol, the probe with the highest mean expression across samples was selected. This procedure yielded 13,237 gene-symbol rows in GSE56815 and 6,271 in GSE2208. Gene symbols were converted to uppercase and case-duplicate entries collapsed for cross-cohort matching and machine-learning analysis, leaving 107 measurable genes among the 190 thematic candidates.

Before uppercase duplicate collapse, the GSE2208 Series Matrix comprised 6,271 mapped gene-symbol rows from 19 samples, including 10 high-BMD and 9 low-BMD samples. Cross-cohort harmonization identified 107 measurable genes among the 190 thematic candidates.

Differential expression in GSE56815 was estimated with limma [17] using a two-group, no-intercept design and the low-BMD minus high-BMD contrast. Following the source-linked nominal differential-expression strategy [11], nominal $P<0.05$ defined the permissive first-stage expression screen. Complete limma results and the $FDR<0.05$ layer were retained for descriptive reporting and quality control, without a fold-change threshold at this stage.

##### S2. MSigDB thematic candidate library

The predefined thematic library comprised 25 MSigDB gene sets covering bone remodeling and osteoclastogenesis, vitamin D/VDR-related regulation, NF-κB signaling, monocyte differentiation and activation, and monocyte/macrophage state or perturbation signatures. The 25 gene sets were obtained from MSigDB and their union contained 1,253 unique genes. Table S1 lists the terms, collection assignments, and descriptions. Intersecting this fixed universe with the nominal GSE56815 screen yielded 190 candidates.

##### S3. Common feature space and machine-learning procedures

The 190 thematic candidates were limited to genes measurable in both the GSE56815 and GSE2208 Series Matrices, yielding 107 common features. Machine-learning models were fitted in GSE56815.

A linear SVM with internal predictor scaling was fitted to the 107 features. Genes were ordered by absolute linear-SVM weights from a full-data fit. Fixed top-ranked subsets of 2 to 40 genes were evaluated with three repeats of 10-fold cross-validation. The maximum internal subset-selection accuracy, 0.9833, occurred for both 24 and 26 genes; the smaller tied subset of 24 genes was retained.

Logistic LASSO was fitted with 10-fold cross-validation using the lambda.1se rule. XGBoost used a shallow tree-based binary classifier with 5-fold stratified cross-validation and early stopping; feature importance was summarized by occurrence frequency. Features were retained through the first rank at which cumulative frequency reached or exceeded 0.50, yielding 9 genes. The three selected sets shared three genes.

#### S4. Cross-cohort transcriptomic support

The three genes in the strict intersection were evaluated independently in GSE56815 and GSE2208. For each gene, the signed low-BMD minus high-BMD expression difference and two-sided Wilcoxon P value were calculated. Single-gene ROC curves were estimated with pROC [23]. ROC direction was selected automatically, and discrimination was summarized as max(AUC, 1-AUC). AUC support required AUC≥0.60 in both cohorts [24]; expression support required nonzero differences in the same direction and P<0.05 in both cohorts. Genes meeting both requirements were eligible for genetic follow-up.

#### S5. Local component-sharing analysis and primary co-exposure definition

OCEL1 was selected as the focal transcript before heel eBMD outcome modeling. Eleven independently selected OCEL1 cis components defined the focal set. Candidate local transcripts were non-OCEL1 chromosome 19 genes within ±1 Mb of OCEL1 with at least one focal-component eQTLGen FDR<0.05 cis-eQTL record, yielding 14 candidates. The complete released eQTLGen FDR<0.05 cis set was evaluated for each candidate transcript. Variants with marginal P<1.829×10^-5 were harmonized to UKBB337K LD, and independent components were selected with cojopy 1.0.0 using a conditional P cutoff of 1×10^-3 and collinearity cutoff $r^2$=0.90.

For every candidate transcript, the maximum UKBB337K LD $r^2$ between each independently selected component and each of the 11 OCEL1 focal components was calculated. Strong sharing was defined as $r^2$≥0.80. BABAM1, MRPL34, and NR2F6 each had two strong matches and were specified as the three primary local co-exposures. DDA1 was the closest transcript below this boundary (maximum $r^2$=0.799005) and was evaluated in a boundary sensitivity analysis.

#### S6. Symmetric four-exposure cis-variant panel

OCEL1, NR2F6, MRPL34, and BABAM1 were treated symmetrically during panel construction. For each candidate variant, exposure eligibility was defined by the smallest association P value across the four transcripts (Pmin<2×10^-5). Variants were mapped to UKBB337K, harmonized across the four exposures, matched to available harmonizable heel eBMD associations, and required heel MAF≥0.01. Variants were ranked by four-exposure Pmin and greedily pruned: after retention, later variants within ±1 Mb and at $r^2$≥0.64 were excluded. Exposure-side evidence determined ranking and panel construction; heel eBMD associations were added after the panel was defined.

The four-exposure candidate union contained 7,791 variants. After exposure-side eligibility and UKBB mapping, 1,752 variants remained; 1,750 had usable exposure allele/frequency information, 1,547 had available harmonizable heel eBMD associations, 1,515 met the MAF requirement, and 268 remained after greedy correlated-instrument pruning. Among the final variants, the smallest exposure P value arose from OCEL1 for 51 variants, NR2F6 for 148, MRPL34 for 57, and BABAM1 for 12.

Palindromic variants in the primary panel were assessed for cross-resource allele-frequency concordance. All 22 retained palindromic variants had an absolute frequency difference ≤0.10 (maximum 0.06248). For the no-palindromic sensitivity analysis, palindromic variants were excluded before LD pruning and the panel was rebuilt independently, yielding 263 variants.

#### S7. R_X estimation and LD regularization

The eQTLGen exposure meta-analysis and the UK Biobank heel eBMD GWAS were obtained from distinct source resources. Their source publications did not report participant overlap, and the released summary data did not permit an exact participant-level overlap count. Exposure-outcome estimation-error correlations were therefore specified as zero.

The 268×268 UKBB337K LD matrix was regularized with POET [30]. Candidate factor counts were assessed by the adjacent eigenvalue-gap ratio, and the regularization parameter was the smallest value on the 0.025–0.25 grid producing a minimum eigenvalue >0.001. The primary solution used K=6 and λ=0.025. The raw matrix had minimum eigenvalue -1.2831×10^-4 and condition number 102,356; after POET regularization, these were 0.009381 and 1,401.

#### S8. CisMRBEEX estimation

Exposure summary associations were standardized as β*=Z/√N and SE*=1/√N. Exposure-side xQTL prefits used two-stage SuSiE-RSS [31]: the first fit used L=15, at most 2,000 iterations, and 95% coverage; the second used L equal to the number of first-pass credible sets plus one under the same convergence settings. The 95% coverage specification followed the CisMRBEEX/SuSiE fine-mapping framework [7,31]. All four prefits converged.

eQTLGen Phase I combines 37 contributing datasets. The released cis-eQTL source used here provides meta-analytic association statistics and association-specific sample sizes, but not the cohort-specific effect estimates needed to reconstruct between-cohort heterogeneity for individual associations. We therefore did not calculate de novo heterogeneity statistics for the extracted cis-eQTL associations.

Within the 268-variant primary panel, eQTLGen sample size was association-specific. Mean sample size was 25,650.85 for OCEL1, NR2F6, and MRPL34 and 19,885.57 for BABAM1. Variant-specific sample sizes entered directly into the transformations $\beta^*=Z/\sqrt{N}$ and $SE^*=1/\sqrt{N}$. Harmonized exposure and heel eBMD associations, allele orientations, allele-frequency information, and source sample sizes for retained variants are supplied in Online Resource 2.

The primary four-exposure CisMRBEEX model [7] incorporated standardized exposure associations, heel eBMD associations, the POET-regularized LD matrix, and the primary empirical R_X. The primary model used a PIP retention threshold of 0.20, a reliability threshold of 0.75, an xQTL PIP threshold of 0.30 [7], and up to 15 xQTL components. Full computational settings are provided with the reproducibility code in Online Resource 2.

#### S9. Robustness and boundary sensitivity analyses

Robustness analyses varied one statistical or exposure-set feature at a time while retaining the focal OCEL1 estimand. Estimation-error analyses used identity R_X, an empirical all-null specification, the primary $r^2<0.10$ empirical R_X, and the stricter $r^2<0.01$ empirical R_X. A same-panel POET analysis fixed K=12. The no-palindromic sensitivity analysis rebuilt the panel before fitting. Local co-exposure sensitivity analyses evaluated OCEL1 alone and in every combination with NR2F6, MRPL34, and BABAM1. DDA1, the closest transcript below the component-sharing boundary, was included in an independently rebuilt five-exposure sensitivity analysis that repeated panel construction, LD estimation, R_X estimation, and CisMRBEEX fitting.

### Supplementary Results

#### S1. Machine-learning convergence and cross-cohort support

The common feature universe contained 107 genes. Linear-SVM ranking retained 24 genes, LASSO retained 18, and XGBoost retained 9; CPNE1, EZR, and OCEL1 formed their strict intersection. OCEL1 alone satisfied the combined cross-cohort support criteria, with low-BMD minus high-BMD differences of -0.07515 in GSE56815 ($P=0.03763$) and -0.12888 in GSE2208 ($P=0.03499$), and AUC magnitudes of 0.6350 and 0.7889, respectively.

#### S2. Local component-sharing boundary

Across 14 local candidate transcripts, BABAM1 and MRPL34 each had a maximum component $r^2$ of 1.0 and two OCEL1 focal-component matches at $r^2 \geq 0.80$. NR2F6 reached a maximum $r^2$ of 0.978938 and also had two strong matches. No other transcript reached the fixed 0.80 boundary. DDA1 was nearest to the boundary ($r^2=0.799005$) and therefore served as the five-exposure boundary sensitivity rather than a primary co-exposure.

#### S3. Primary panel and allele-frequency audit

The symmetric selection procedure yielded a correlated cis panel of 268 variants. The maximum retained pairwise $r^2$ was 0.6394, consistent with the $r^2<0.64$ blocking rule. Twenty-two retained variants were palindromic, and each met the cross-resource frequency-concordance criterion of an absolute difference $\leq 0.10$.

#### S4. Primary four-exposure model

The formal primary CisMRBEEX fit retained OCEL1 and BABAM1 at the fixed PIP threshold of 0.20. OCEL1 had $\beta=0.010293$, SE=0.004107, 95% CI 0.002244–0.018342, $P=0.01220$, and PIP=0.4001. BABAM1 had $\beta=-0.014290$, SE=0.005631, 95% CI -0.025327 to -0.003252, $P=0.01117$, and PIP=0.4125. NR2F6 and MRPL34 had PIPs of 0.0930 and 0.0945 and were not retained by the sparse model.

#### S5. Robustness of the focal OCEL1 estimate

Across the four R_X definitions, the OCEL1 coefficient was nearly unchanged ($\beta=0.010283$–$0.010312$; $P=0.01208$–$0.01226$). Fixing POET at K=12 gave $\beta=0.009155$ ($P=0.02465$). Across eight local co-exposure specifications, the coefficient remained positive ($\beta=0.008710$–$0.011469$; $P=0.00769$–$0.03335$). The independently rebuilt no-palindromic sensitivity analysis remained positive but was attenuated and did not meet the nominal $P<0.05$ threshold ($\beta=0.007652$, 95% CI -0.000548 to 0.015852, $P=0.06739$).

#### S6. DDA1 boundary sensitivity analysis

The independently reconstructed five-exposure analysis including DDA1 produced an essentially unchanged OCEL1 coefficient ($\beta=0.010298$, SE=0.004107, $P=0.01217$) relative to the primary four-exposure model ($\beta=0.010293$). BABAM1 remained negative ($\beta=-0.014297$, $P=0.01114$). DDA1 had PIP=0.1112 and was not retained at the 0.20 threshold. The five-exposure panel contained 268 variants and used POET K=6 with $\lambda=0.025$.

#### Supplementary Tables

##### Table S1. Predefined 25-gene-set MSigDB thematic library.

| Gene set | Collection | Subcollection | Description |
|---|---|---|---|
| BIOCARTA_VDR_PATHWAY | C2 | CP:BIOCARTA | Control of Gene Expression by Vitamin D Receptor |
| GOBP_BONE_REMODELING | C5 | GO:BP | The continuous turnover of bone matrix and mineral that involves first, an increase in resorption (osteoclastic activity) and later, reactive bone formation (osteoblastic activity). The process of bone remodeling takes place in the adult skeleton at discrete foci. The process ensures the mechanical integrity of the skeleton throughout life and plays an important role in calcium homeostasis. An imbalance in the regulation of bone resorption and bone formation results in many of the metabolic bone diseases, such as osteoporosis. [GOC:curators] |
| GOBP_MONOCYTE_ACTIVATION | C5 | GO:BP | The change in morphology and behavior of a monocyte resulting from exposure to a cytokine, chemokine, cellular ligand, or soluble factor. [GOC:mgi_curators, ISBN:0781735149] |
| GOBP_MONOCYTE_DIFFERENTIATION | C5 | GO:BP | The process in which a relatively unspecialized myeloid precursor cell acquires the specialized features of a monocyte. [GOC:mah] |

| Gene set | Collection | Subcollection | Description |
|---|---|---|---|
| GOBP_MULTINUCLEAR_OSTEOCLAST_DIFFERENTIATION | C5 | GO:BP | The process in which a relatively unspecialized monocyte acquires the specialized features of a multinuclear osteoclast. An osteoclast is a specialized phagocytic cell associated with the absorption and removal of the mineralized matrix of bone tissue. [CL:0000779, GOC:mah, PMID:12713016] |
| GOBP_NEGATIVE_REGULATION_OF_I_KAPPAB_KINASE_NF_KAPPAB_SIGNALING | C5 | GO:BP | Any process that stops, prevents, or reduces the frequency, rate or extent of -kappaB kinase/NF-kappaB signaling. [GOC:jl] |
| GOBP_NEGATIVE_REGULATION_OF_NIK_NF_KAPPAB_SIGNALING | C5 | GO:BP | Any process that stops, prevents or reduces the frequency, rate or extent of NIK/NF-kappaB signaling. [GOC:TermGenie] |
| GOBP_NEGATIVE_REGULATION_OF_OSTEOCLAST_DIFFERENTIATION | C5 | GO:BP | Any process that stops, prevents, or reduces the frequency, rate or extent of osteoclast differentiation. [GOC:go_curators] |
| GOBP_OSTEOCLAST_DIFFERENTIATION | C5 | GO:BP | The process in which a relatively unspecialized monocyte acquires the specialized features of an osteoclast. An osteoclast is a specialized phagocytic cell associated with the absorption and removal of the mineralized matrix of bone tissue. [CL:0000092, GOC:add, ISBN:0781735149, PMID:12161749] |

| Gene set | Collectio n | Subcollection | Description |
|---|---|---|---|
| GOBP_OSTEOCLAST_PROLIFERATION | C5 | GO:BP | The multiplication or reproduction of osteoclasts, resulting in the expansion of an osteoclast cell population. An osteoclast is a specialized phagocytic cell associated with the absorption and removal of the mineralized matrix of bone tissue, which typically differentiates from monocytes. [CL:0000092, GOC:hjd] |
| GOBP_POSITIVE_REGULATION_OF_BONE_RESORPTION | C5 | GO:BP | Any process that activates or increases the frequency, rate or extent of bone resorption. [GOC:go_curators] |
| GOBP_POSITIVE_REGULATION_OF_I_KAPPAB_KINASE_NF_KAPPAB_SIGNALING | C5 | GO:BP | Any process that activates or increases the frequency, rate or extent of I-kappaB kinase/NF-kappaB signaling. [GOC:jl] |
| GOBP_POSITIVE_REGULATION_OF_NIK_NF_KAPPAB_SIGNALING | C5 | GO:BP | Any process that activates or increases the frequency, rate or extent of NIK/NF-kappaB signaling. [GOC:TermGenie] |
| GOBP_POSITIVE_REGULATION_OF_OSTEOCLAST_DIFFERENTIATION | C5 | GO:BP | Any process that activates or increases the frequency, rate or extent of osteoclast differentiation. [GOC:go_curators] |
| GOBP_POSITIVE_REGULATION_OF_VITAMIN_D_RECEPTOR_SIGNALING_PATHWAY | C5 | GO:BP | Any process that activates or increases the frequency, rate or extent of vitamin D receptor signaling pathway activity. [GOC:BHF, GOC:mah] |
| GOBP_REGULATION_OF_MONOCYTE_DIFFERENTIATION | C5 | GO:BP | Any process that modulates the frequency, rate or extent of monocyte differentiation. [GOC:go_curators] |

| Gene set | Collection | Subcollection | Description |
|---|---|---|---|
| GOBP_REGULATION_OF_NIK_NF_KAPPAB_SIGNALING | C5 | GO:BP | Any process that modulates the frequency, rate or extent of NIK/NF-kappaB signaling. [GOC:TermGenie] |
| GOBP_REGULATION_OF_OSTEOCLAST_DIFFERENTIATION | C5 | GO:BP | Any process that modulates the frequency, rate or extent of osteoclast differentiation. [GOC:go_curators] |
| GOBP_REGULATION_OF_VITAMIN_D_RECEPTOR_SIGNALING_PATHWAY | C5 | GO:BP | Any process that modulates the frequency, rate or extent of vitamin D receptor signaling pathway activity. [GOC:BHF, GOC:mah] |
| GOBP_VITAMIN_D_RECEPTOR_SIGNALING_PATHWAY | C5 | GO:BP | The series of molecular signals generated as a consequence of a vitamin D receptor binding to one of its physiological ligands. [GOC:BHF, GOC:mah, PMID:12637589] |
| GOMF_VITAMIN_D_RECEPTOR_BINDING | C5 | GO:MF | Binding to a vitamin D receptor, a nuclear receptor that mediates the action of vitamin D by binding DNA and controlling the transcription of hormone-sensitive genes. [GOC:jl, PMID:12637589] |
| GSE16385_MONOCYTE_VS_12H_IL4_TREATED_MACROPHAGE_DN | C7 | IMMUNESIGDB | Genes down-regulated in monocytes (12h) versus macrophages (12h) treated with IL4 [GeneID=3565]. |
| GSE32034_LY6C_HIGH_VS_LOW_ROSIGLIZATONE_TREATED_MONOCYTE_DN | C7 | IMMUNESIGDB | Genes down-regulated in monocytes treated by rosiglitazone [PubChem=77999]: Ly6C high versus Ly6C low. |
| GSE32034_UNTREATED_VS_ROSIGLIZATONE_TREATED_LY6C_HIGH_MONOCYTE_UP | C7 | IMMUNESIGDB | Genes up-regulated in Ly6C high monocytes: untreated versus rosiglitazone [PubChem=77999]. |

| Gene set | Collectio n | Subcollection | Description |
|---|---|---|---|
| GSE9988_LPS_VS_LOW_LPS_MONOCYTE_DN | C7 | IMMUNESIGD B | Genes down-regulated in comparison of monocytes treated with 5000 ng/ml LPS (TLR4 agonist) versus those treated with 1 ng/ml LPS (TLR4 agonist). |

**Table S2. Genes retained by the three machine-learning procedures.**

| Method | N | Selected genes |
|---|---|---|
| Linear SVM weight ranking | 24 | FAM216A, PTPN1, ZNF281, ABR, CLEC7A, CYTH1, OCEL1, CPNE1, EZR, MAP3K3, NPC1, NPY, PIK3R4, FXN, IL17RA, P2RY2, ANXA2, ADAM9, ATP2C1, MT1G, C12ORF43, PIM1, SMARCC2, SRC |
| LASSO (lambda.1se) | 18 | ABR, ATP2C1, CD33, CHI3L1, CPNE1, EDA, EZR, FAM216A, FXN, IL17RA, LMAN2L, MAP3K3, NCOA1, NPC1, NPY, OCEL1, PTPN1, RC3H2 |
| XGBoost (cumulative frequency ≥0.50) | 9 | EZR, LMAN2L, OCEL1, GATAD2A, NFKBIA, CPT2, ZFP36L1, P2RY2, CPNE1 |
| Strict three-model intersection | 3 | CPNE1, EZR, OCEL1 |

**Table S3. Cross-cohort transcriptomic support for the three convergent genes.**

| Gene | GSE56815 AUC | GSE2208 AUC | GSE56815 Δ | GSE2208 Δ | GSE56815 P | GSE2208 P | Direction concordant | Final support |
|---|---|---|---|---|---|---|---|---|
| CPNE1 | 0.6687 | 0.7444 | -0.20645 | 0.20537 | 0.008991 | 0.0790478 | No | No |
| EZR | 0.6687 | 0.7556 | -0.15207 | 0.29266 | 0.008991 | 0.0652536 | No | No |
| OCEL1 | 0.6350 | 0.7889 | -0.07515 | -0.12888 | 0.0376273 | 0.0349867 | Yes | Yes |

**Table S4. Component sharing between OCEL1 focal components and 14 local candidate transcripts.**

| Candidate transcript | Maximum $r^2$ | Matches $r^2 \geq 0.80$ | Matches $r^2 \geq 0.50$ | Exact matches |
|---|---|---|---|---|
| BABAM1 | 1.000000 | 2 | 3 | 1 |
| MRPL34 | 1.000000 | 2 | 5 | 1 |
| NR2F6 | 0.978938 | 2 | 6 | 0 |
| DDA1 | 0.799005 | 0 | 1 | 0 |
| GTPBP3 | 0.719864 | 0 | 2 | 0 |
| ABHD8 | 0.671113 | 0 | 1 | 0 |
| ANKLE1 | 0.586016 | 0 | 1 | 0 |
| USE1 | 0.324150 | 0 | 0 | 0 |
| MVB12A | 0.312009 | 0 | 0 | 0 |

| Candidate transcript | Maximum r² | Matches r²≥0.80 | Matches r²≥0.50 | Exact matches |
|---|---|---|---|---|
| GLT25D1 | 0.311543 | 0 | 0 | 0 |
| HAUS8 | 0.303486 | 0 | 0 | 0 |
| PLVAP | 0.284591 | 0 | 0 | 0 |
| MYO9B | 0.203866 | 0 | 0 | 0 |
| CTD-3131K8.2 | 0.056382 | 0 | 0 | 0 |

#### Table S5A. Symmetric four-exposure panel-construction flow.

| Panel-construction stage | Variants |
|---|---|
| Four-exposure candidate union | 7791 |
| Exposure-eligible and UKBB-mapped | 1752 |
| Exposure allele/frequency usable | 1750 |
| Heel technically usable | 1547 |
| Heel MAF ≥0.01 | 1515 |
| Primary correlated cis panel | 268 |
| No-palindromic rebuilt panel | 263 |

#### Table S5B. Exposure-side driver of the 268 retained variants.

| Exposure with minimum P among four exposures | Retained variants |
|---|---|
| OCEL1 | 51 |
| NR2F6 | 148 |
| MRPL34 | 57 |
| BABAM1 | 12 |

#### Table S6. Allele-frequency audit of palindromic variants retained in the primary panel.

| SNP | Panel alleles | eQTL A1 frequency | Heel A1 frequency | \|Δ frequency\| | Status |
|---|---|---|---|---|---|
| RS34180454 | A/T | 0.8213 | 0.8158 | 0.0055 | Concordant ≤0.10 |
| RS11086060 | C/G | 0.0874 | 0.0896 | 0.0023 | Concordant ≤0.10 |
| RS62126222 | G/C | 0.9333 | 0.9190 | 0.0144 | Concordant ≤0.10 |
| RS12984127 | G/C | 0.5814 | 0.5771 | 0.0043 | Concordant ≤0.10 |
| RS117558513 | G/C | 0.9596 | 0.9687 | 0.0090 | Concordant ≤0.10 |
| RS10419742 | G/C | 0.9071 | 0.8990 | 0.0081 | Concordant ≤0.10 |
| RS117518483 | C/G | 0.9387 | 0.9365 | 0.0023 | Concordant ≤0.10 |
| RS10420922 | A/T | 0.5633 | 0.5643 | 0.0010 | Concordant ≤0.10 |
| RS76900423 | A/T | 0.9049 | 0.9066 | 0.0016 | Concordant ≤0.10 |
| RS74253216 | C/G | 0.9506 | 0.9526 | 0.0020 | Concordant ≤0.10 |
| RS388484 | A/T | 0.3869 | 0.3824 | 0.0045 | Concordant ≤0.10 |
| RS12984771 | T/A | 0.5334 | 0.5058 | 0.0276 | Concordant ≤0.10 |
| RS76526930 | T/A | 0.9398 | 0.9379 | 0.0019 | Concordant ≤0.10 |
| RS73499195 | G/C | 0.7814 | 0.7983 | 0.0169 | Concordant ≤0.10 |
| RS34761413 | C/G | 0.6259 | 0.5976 | 0.0283 | Concordant ≤0.10 |
| RS6512193 | A/T | 0.0313 | 0.0197 | 0.0116 | Concordant ≤0.10 |
| RS7256385 | G/C | 0.1439 | 0.0814 | 0.0625 | Concordant ≤0.10 |
| RS2099045 | T/A | 0.1171 | 0.0828 | 0.0343 | Concordant ≤0.10 |
| RS62126223 | G/C | 0.9676 | 0.9655 | 0.0021 | Concordant ≤0.10 |
| RS73018481 | T/A | 0.4862 | 0.4626 | 0.0235 | Concordant ≤0.10 |
| RS12979168 | C/G | 0.7930 | 0.7932 | 0.0002 | Concordant ≤0.10 |
| RS6512190 | T/A | 0.0622 | 0.0428 | 0.0194 | Concordant ≤0.10 |

#### Table S7A. Construction of the variant set used for estimation-error correlation.

| R_X null-pool stage | Variants |
|---|---|
| Common harmonized four-exposure cis variants | 6403 |
| After formal selected-variant exclusion | 4805 |
| max\|Z\|≤2 | 3041 |
| Mapped to UKBB337K | 2918 |
| LD-thinned r²<0.10 (primary) | 400 |
| LD-thinned r²<0.01 | 190 |

**Table S7B. Primary empirical estimation-error correlation matrix (R_X with heel eBMD error correlations fixed to zero).**

| Variable | OCEL1 | NR2F6 | MRPL34 | BABAM1 | heel_eBMD |
|---|---|---|---|---|---|
| OCEL1 | 1.0000 | 0.0091 | -0.0054 | -0.0468 | 0.0000 |
| NR2F6 | 0.0091 | 1.0000 | 0.0506 | 0.0407 | 0.0000 |
| MRPL34 | -0.0054 | 0.0506 | 1.0000 | 0.0869 | 0.0000 |
| BABAM1 | -0.0468 | 0.0407 | 0.0869 | 1.0000 | 0.0000 |
| heel_eBMD | 0.0000 | 0.0000 | 0.0000 | 0.0000 | 1.0000 |

**Table S8. Formal primary four-exposure CisMRBEEX results.**

| Exposure | β | SE | 95% CI | P | PIP | Status |
|---|---|---|---|---|---|---|
| OCEL1 | 0.010293 | 0.004107 | 0.002244 to 0.018342 | 0.012198 | 0.4001 | Retained |
| NR2F6 | 0.000000 | 0.000000 | Not applicable | | 0.0930 | Not retained |
| MRPL34 | 0.000000 | 0.000000 | Not applicable | | 0.0945 | Not retained |
| BABAM1 | -0.014290 | 0.005631 | -0.025327 to -0.003252 | 0.011166 | 0.4125 | Retained |

**Table S9A. OCEL1 estimate across R_X, POET, and no-palindromic sensitivity analyses.**

| Specification | OCEL1 β | SE | 95% CI | P |
|---|---|---|---|---|
| Identity R_X | 0.010283 | 0.004106 | 0.002236 to 0.018330 | 0.012257 |
| Empirical all-null R_X | 0.010312 | 0.004109 | 0.002259 to 0.018365 | 0.01208 |
| Empirical $r^2<0.10$ R_X | 0.010293 | 0.004107 | 0.002244 to 0.018342 | 0.012198 |
| Empirical $r^2<0.01$ R_X | 0.010300 | 0.004107 | 0.002250 to 0.018350 | 0.012154 |
| POET K=12 | 0.009155 | 0.004074 | 0.001169 to 0.017140 | 0.024645 |
| No-palindromic rebuilt panel | 0.007652 | 0.004184 | -0.000548 to 0.015852 | 0.067387 |

**Table S9B. OCEL1 estimates across local co-exposure specifications.**

| Local model | OCEL1 β | SE | 95% CI | P |
|---|---|---|---|---|
| OCEL1 only | 0.008710 | 0.004093 | 0.000687 to 0.016733 | 0.033354 |
| OCEL1 + NR2F6 | 0.009322 | 0.004110 | 0.001266 to 0.017378 | 0.023327 |
| OCEL1 + MRPL34 | 0.011469 | 0.004303 | 0.003036 to 0.019902 | 0.0076872 |
| OCEL1 + BABAM1 | 0.010293 | 0.004107 | 0.002244 to 0.018342 | 0.012198 |
| OCEL1 + NR2F6 + MRPL34 | 0.008710 | 0.004093 | 0.000687 to 0.016733 | 0.033354 |
| OCEL1 + NR2F6 + BABAM1 | 0.010293 | 0.004107 | 0.002244 to 0.018342 | 0.012198 |
| OCEL1 + MRPL34 + BABAM1 | 0.010293 | 0.004107 | 0.002244 to 0.018342 | 0.012198 |
| OCEL1 + NR2F6 + MRPL34 + BABAM1 | 0.010293 | 0.004107 | 0.002244 to 0.018342 | 0.012198 |

**Table S10. Independently reconstructed five-exposure DDA1 sensitivity analysis.**

| Exposure | β | SE | 95% CI | P | PIP | Status |
|---|---|---|---|---|---|---|
| OCEL1 | 0.010298 | 0.004107 | 0.002248 to 0.018348 | 0.012166 | 0.3563 | Retained |
| NR2F6 | 0.000000 | 0.000000 | Not applicable | | 0.0862 | Not retained |
| MRPL34 | 0.000000 | 0.000000 | Not applicable | | 0.0873 | Not retained |
| BABAM1 | -0.014297 | 0.005632 | -0.025336 to -0.003258 | 0.011137 | 0.3591 | Retained |
| DDA1 | 0.000000 | 0.000000 | Not applicable | | 0.1112 | Not retained |

**Abbreviations:** AUC, area under the receiver operating characteristic curve; BMD, bone mineral density; eBMD, estimated bone mineral density; LD, linkage disequilibrium; MAF, minor-allele frequency; PIP, posterior inclusion probability; POET, principal orthogonal complement thresholding; R_X, exposure-side estimation-error correlation matrix; SVM, support vector machine.
