## Supplementary material for "Transcriptomic and Machine-Learning Prioritization of OCEL1 in Low Bone Mineral Density With Multivariable cis-MR": Online Resource 2: Reproducibility Materials: 01_unified_gpl96_preprocess.R

```
#!/usr/bin/env Rscript
# Canonical unified GPL96 preprocessing for the Paper 1 bulk transcriptomic
# cohorts. Both cohorts start from locally cached public GEO Series Matrix and
# SOFT files, then use the same probe-to-gene preprocessing functions.

suppressPackageStartupMessages({
  library(limma)
})

args <- commandArgs(trailingOnly = TRUE)
root <- if (length(args)) normalizePath(args[[1]], winslash = "/", mustWork = TRUE) else getwd()
geo_dir <- file.path(root, "data", "geo")
out_dir <- file.path(root, "outputs")
gpl96_source <- Sys.getenv(
  "GPL96_SOURCE",
  unset = file.path(root, "data", "geo", "GPL96_family.soft.gz")
)
dir.create(out_dir, recursive = TRUE, showWarnings = FALSE)

read_gz_lines <- function(path) {
  con <- gzfile(path, open = "rt")
  on.exit(close(con), add = TRUE)
  readLines(con, warn = FALSE)
}

parse_series_matrix <- function(path) {
  x <- read_gz_lines(path)
  begin <- grep("^!series_matrix_table_begin", x)
  end <- grep("^!series_matrix_table_end", x)
  if (length(begin) != 1L || length(end) != 1L || end <= begin) stop("Malformed Series Matrix: ", basename(path))
  tbl <- read.delim(text = paste(x[(begin + 1L):(end - 1L)], collapse = "\n"), header = TRUE,
                    sep = "\t", quote = "\"", check.names = FALSE, stringsAsFactors = FALSE)
  ids <- tbl[[1L]]
  mat <- as.matrix(tbl[, -1L, drop = FALSE])
  storage.mode(mat) <- "numeric"
  rownames(mat) <- ids
  mat
}

parse_soft_groups <- function(path, dataset) {
  x <- read_gz_lines(path)
  starts <- grep("^\\^SAMPLE = ", x)
  if (!length(starts)) stop("No sample blocks in ", basename(path))
  rows <- lapply(seq_along(starts), function(i) {
    block <- x[starts[i]:(if (i < length(starts)) starts[i + 1L] - 1L else length(x))]
    id <- sub("^!Sample_geo_accession = ", "", grep("^!Sample_geo_accession = ", block, value = TRUE)[1L])
    text_lines <- sub("^!Sample_(title|source_name_ch1|characteristics_ch1|description) = ", "",
      grep("^!Sample_(title|source_name_ch1|characteristics_ch1|description) = ", block, value = TRUE))
    text <- paste(text_lines, collapse = " | ")
    # Retain only a line that names one BMD arm. This excludes the GSE2208
    # study-wide description mentioning both arms while accepting the GSE56815
    # title and the GSE2208 sample-specific description.
    text_low <- tolower(text_lines)
    is_one_arm <- xor(grepl("high bmd", text_low, fixed = TRUE), grepl("low bmd", text_low, fixed = TRUE))
    label_lines <- text_lines[is_one_arm]
    label <- if (length(label_lines)) tolower(tail(label_lines, 1L)) else ""
    group <- if (grepl("low bmd", label, fixed = TRUE)) "low_bmd" else if (grepl("high bmd", label, fixed = TRUE)) "high_bmd" else NA_character_
    data.frame(dataset = dataset, sample = id, group = group, title = text, stringsAsFactors = FALSE)
  })
  out <- do.call(rbind, rows)
  out[!is.na(out$group), , drop = FALSE]
}

# The local GSE2208 SOFT contains the GPL96 platform table. This parser follows
# the canonical Paper 1 probe-symbol treatment: first Gene Symbol before any
# " /// " or " // " multi-annotation separator.
parse_gpl96_symbol_map <- function(path, table_cache) {
  if (!file.exists(path)) stop("Missing GPL96 annotation: ", path)
  # The platform file is large. Stream only the first platform table and
  # stop at !platform_table_end after extracting the canonical platform table.
  con <- gzfile(path, open = "rt")
  out <- file(table_cache, open = "wt")
  on.exit({ if (!is.null(con)) close(con); if (!is.null(out)) close(out) }, add = TRUE)
  in_table <- FALSE
  n_written <- 0L
  repeat {
    chunk <- readLines(con, n = 50000L, warn = FALSE)
    if (!length(chunk)) break
    for (line in chunk) {
      if (!in_table) {
        if (identical(line, "!platform_table_begin")) in_table <- TRUE
        next
      }
      if (identical(line, "!platform_table_end")) {
        close(out)
        out <- NULL
        close(con)
        con <- NULL
        break
      }
      writeLines(line, out)
      n_written <- n_written + 1L
    }
    if (is.null(out)) break
  }
  if (n_written == 0L) stop("GPL96 platform table missing from ", basename(path))
  df <- read.delim(table_cache, sep = "\t", header = TRUE,
                   quote = "", comment.char = "", check.names = FALSE, stringsAsFactors = FALSE)
  id_col <- names(df)[1L]
  symbol_candidates <- names(df)[grepl("^gene[ _]?symbol$|gene[ _]?symbol|^symbol$", tolower(names(df)))]
  if (!length(symbol_candidates)) stop("No Gene Symbol column in local GPL96 table")
  map <- data.frame(ID = as.character(df[[id_col]]), symbol = as.character(df[[symbol_candidates[1L]]]), stringsAsFactors = FALSE)
  map$symbol <- trimws(sub(" //.*$", "", sub(" ///.*$", "", map$symbol)))
  map <- map[!is.na(map$ID) & nzchar(map$ID) & !is.na(map$symbol) & nzchar(map$symbol) & map$symbol != "---" & map$symbol != "NA", , drop = FALSE]
  unique(map)
}

# Shared preprocessing begins here. This function is intentionally called for
# both Series Matrix inputs without cohort-specific branches.
normalize_if_needed <- function(expr_probe) {
  q <- as.numeric(quantile(expr_probe, c(0, .25, .5, .75, .99, 1), na.rm = TRUE))
  is_log_scale <- q[5] < 100 && (q[6] - q[1]) < 50
  if (is_log_scale) return(list(expr = expr_probe, action = "already_log_scale"))
  if (min(expr_probe, na.rm = TRUE) < 0) stop("Non-log expression contains negative values")
  list(expr = log2(expr_probe + 1), action = "log2_plus_1")
}

probe_to_gene_highest_mean <- function(expr_probe, map_df) {
  all_na <- rowSums(!is.na(expr_probe)) == 0L
  expr_probe <- expr_probe[!all_na, , drop = FALSE]
  for (i in seq_len(nrow(expr_probe))) {
    miss <- is.na(expr_probe[i, ])
    if (any(miss)) expr_probe[i, miss] <- median(expr_probe[i, !miss], na.rm = TRUE)
  }
  map_df <- map_df[!duplicated(map_df$ID), c("ID", "symbol"), drop = FALSE]
  symbol <- map_df$symbol[match(trimws(rownames(expr_probe)), map_df$ID)]
  keep <- !is.na(symbol) & nzchar(symbol)
  # Preserve GPL96 symbol case until the explicit cross-cohort harmonization step.
  df <- data.frame(symbol = unname(symbol[keep]), expr_probe[keep, , drop = FALSE], check.names = FALSE, stringsAsFactors = FALSE)
  df$mean_expression <- rowMeans(df[, setdiff(names(df), c("symbol", "mean_expression")), drop = FALSE])
  df <- df[order(df$symbol, -df$mean_expression), , drop = FALSE]
  df <- df[!duplicated(df$symbol), , drop = FALSE]
  rownames(df) <- df$symbol
  as.matrix(df[, setdiff(names(df), c("symbol", "mean_expression")), drop = FALSE])
}

limma_result <- function(expr, groups) {
  groups <- groups[match(colnames(expr), groups$sample), , drop = FALSE]
  if (!identical(unname(as.character(groups$sample)), unname(as.character(colnames(expr))))) {
    stop("Expression columns and group metadata are not in the same sample order")
  }
  groups$group <- factor(groups$group, levels = c("high_bmd", "low_bmd"))
  design <- model.matrix(~0 + group, groups)
  colnames(design) <- levels(groups$group)
  fit <- eBayes(contrasts.fit(lmFit(expr, design), makeContrasts(low_bmd - high_bmd, levels = design)))
  res <- topTable(fit, coef = 1, number = Inf, sort.by = "P", adjust.method = "BH")
  data.frame(symbol = rownames(res), res[, c("logFC", "P.Value", "adj.P.Val")], row.names = NULL, check.names = FALSE)
}

datasets <- c("GSE56815", "GSE2208")
platform_table_cache <- file.path(out_dir, "GPL96_PLATFORM_TABLE.tsv")
platform_map <- parse_gpl96_symbol_map(gpl96_source, platform_table_cache)
write.csv(platform_map, file.path(out_dir, "GPL96_PLATFORM_PROBE_SYMBOL_MAP.csv"), row.names = FALSE)
write.csv(data.frame(gpl96_annotation = basename(gpl96_source), platform_table_rows = nrow(platform_map), stringsAsFactors = FALSE), file.path(out_dir, "GPL96_MAPPING_PROVENANCE.csv"), row.names = FALSE)
processed <- list()
groups <- list()
actions <- character()
raw_features <- integer()
for (dataset in datasets) {
  expr_file <- file.path(geo_dir, paste0(dataset, "_series_matrix.txt.gz"))
  soft_file <- file.path(geo_dir, paste0(dataset, "_family.soft.gz"))
  if (!file.exists(expr_file) || !file.exists(soft_file)) stop("Missing local GEO input for ", dataset)
  norm <- normalize_if_needed(parse_series_matrix(expr_file))
  raw_features[dataset] <- nrow(norm$expr)
  write.csv(data.frame(probe_id = rownames(norm$expr), stringsAsFactors = FALSE), file.path(out_dir, paste0(dataset, "_series_matrix_probe_ids.csv")), row.names = FALSE)
  meta <- parse_soft_groups(soft_file, dataset)
  gene <- probe_to_gene_highest_mean(norm$expr, platform_map)
  write.csv(data.frame(sample = colnames(gene), stringsAsFactors = FALSE), file.path(out_dir, paste0(dataset, "_matrix_sample_ids.csv")), row.names = FALSE)
  write.csv(meta, file.path(out_dir, paste0(dataset, "_SOFT_group_ids.csv")), row.names = FALSE)
  keep <- intersect(colnames(gene), meta$sample)
  processed[[dataset]] <- gene[, keep, drop = FALSE]
  groups[[dataset]] <- meta[match(keep, meta$sample), , drop = FALSE]
  actions[dataset] <- norm$action
  write.csv(data.frame(symbol = rownames(processed[[dataset]]), processed[[dataset]], check.names = FALSE), file.path(out_dir, paste0(dataset, "_unified_highest_mean_expr.csv")), row.names = FALSE)
  write.csv(groups[[dataset]], file.path(out_dir, paste0(dataset, "_unified_groups.csv")), row.names = FALSE)
  if (dataset == "GSE56815") {
    write.csv(limma_result(processed[[dataset]], groups[[dataset]]), file.path(out_dir, "GSE56815_unified_limma.csv"), row.names = FALSE)
  }
}

manifest <- data.frame(dataset = datasets, expression_adapter = "local_GEO_series_matrix", metadata_adapter = "local_GEO_family_SOFT", normalization_action = unname(actions[datasets]), probe_features = unname(raw_features[datasets]), final_genes = vapply(processed, nrow, integer(1)), final_samples = vapply(processed, ncol, integer(1)))
write.csv(manifest, file.path(out_dir, "FINAL_PREPROCESS_MANIFEST.csv"), row.names = FALSE)
```
