## Supplementary material for "Transcriptomic and Machine-Learning Prioritization of OCEL1 in Low Bone Mineral Density With Multivariable cis-MR": Online Resource 2: Reproducibility Materials: 00_paths.R

```
script_file <- grep('^--file=', commandArgs(FALSE), value = TRUE)
script_dir <- if (length(script_file)) dirname(normalizePath(sub('^--file=', '', script_file[[1]]), winslash = '/', mustWork = FALSE)) else normalizePath(getwd(), winslash = '/', mustWork = FALSE)
PROJECT_ROOT <- normalizePath(file.path(script_dir, '..'), winslash = '/', mustWork = TRUE)
DATA_ROOT <- normalizePath(file.path(PROJECT_ROOT, '..', '00_DATA_PAPER1'), winslash = '/', mustWork = TRUE)
OUTPUT_ROOT <- file.path(PROJECT_ROOT, 'outputs')
dir.create(OUTPUT_ROOT, recursive = TRUE, showWarnings = FALSE)
```
