## Supplementary material for "Transcriptomic and Machine-Learning Prioritization of OCEL1 in Low Bone Mineral Density With Multivariable cis-MR": Online Resource 2: Reproducibility Materials: 01_gse56815_limma.R

```
source(file.path(dirname(normalizePath(sub('^--file=', '', grep('^--file=', commandArgs(FALSE), value = TRUE)[1]), winslash = '/', mustWork = FALSE)), '00_paths.R'))
suppressPackageStartupMessages(library(limma))
expr <- read.csv(file.path(DATA_ROOT, 'bulk', 'normalized_expression', 'GSE56815_expr_by_symbol.csv'), row.names = 1, check.names = FALSE)
meta <- read.csv(file.path(DATA_ROOT, 'bulk', 'sample_metadata', 'GSE56815_group.csv'), stringsAsFactors = FALSE)
stopifnot(identical(meta$sample, colnames(expr)))
meta$group <- factor(meta$group, levels = c('high_bmd', 'low_bmd'))
design <- model.matrix(~0 + group, meta)
colnames(design) <- levels(meta$group)
fit <- eBayes(contrasts.fit(lmFit(as.matrix(expr), design), makeContrasts(low_bmd - high_bmd, levels = design)))
all_res <- topTable(fit, coef = 1, number = Inf, sort.by = 'P', adjust.method = 'BH')
out <- data.frame(symbol = rownames(all_res), all_res[, c('logFC', 'P.Value', 'adj.P.Val')], row.names = NULL, check.names = FALSE)
dir.create(file.path(OUTPUT_ROOT, '01_bulk'), recursive = TRUE, showWarnings = FALSE)
write.csv(out, file.path(OUTPUT_ROOT, '01_bulk', 'GSE56815_limma_all_genes.csv'), row.names = FALSE)
write.csv(out[out$P.Value < 0.05, ], file.path(OUTPUT_ROOT, '01_bulk', 'GSE56815_nominal_P_lt_0.05.csv'), row.names = FALSE)
write.csv(out[out$adj.P.Val < 0.05, ], file.path(OUTPUT_ROOT, '01_bulk', 'GSE56815_FDR_lt_0.05.csv'), row.names = FALSE)
heatmap_genes <- head(out$symbol, 30L)
heatmap_expr <- expr[heatmap_genes, meta$sample, drop = FALSE]
write.table(data.frame(symbol = rownames(heatmap_expr), heatmap_expr, check.names = FALSE),
            file.path(OUTPUT_ROOT, '01_bulk', 'GSE56815_heatmap_source.tsv'),
            sep = '\t', quote = FALSE, row.names = FALSE)
write.table(data.frame(symbol = rownames(expr), expr, check.names = FALSE),
            file.path(OUTPUT_ROOT, '01_bulk', 'GSE56815_expression_plotting_source.tsv'),
            sep = '\t', quote = FALSE, row.names = FALSE)
write.table(data.frame(symbol = heatmap_genes, display_order = seq_along(heatmap_genes)),
            file.path(OUTPUT_ROOT, '01_bulk', 'GSE56815_heatmap_gene_order.tsv'),
            sep = '\t', quote = FALSE, row.names = FALSE)
write.table(data.frame(sample = meta$sample, group = as.character(meta$group), display_order = seq_len(nrow(meta))),
            file.path(OUTPUT_ROOT, '01_bulk', 'GSE56815_heatmap_sample_metadata.tsv'),
            sep = '\t', quote = FALSE, row.names = FALSE)
message('PASS: GSE56815 limma result generated.')
```
