## Supplementary material for "Transcriptomic and Machine-Learning Prioritization of OCEL1 in Low Bone Mineral Density With Multivariable cis-MR": Online Resource 2: Reproducibility Materials: 02_thematic_candidate_screen.R

```
source(file.path(dirname(normalizePath(sub('^--file=', '', grep('^--file=', commandArgs(FALSE), value = TRUE)[1]), winslash = '/', mustWork = FALSE)), '00_paths.R'))
deg <- read.csv(file.path(OUTPUT_ROOT, '01_bulk', 'GSE56815_nominal_P_lt_0.05.csv'), check.names = FALSE)
library_genes <- read.csv(file.path(DATA_ROOT, 'bulk', 'thematic_library', 'thematic_gene_universe_1253.csv'), check.names = FALSE)
gene_col <- if ('gene' %in% names(library_genes)) 'gene' else names(library_genes)[1]
candidates <- sort(intersect(toupper(deg$symbol), toupper(library_genes[[gene_col]])))
out <- data.frame(gene = candidates)
dir.create(file.path(OUTPUT_ROOT, '02_candidates'), recursive = TRUE, showWarnings = FALSE)
write.csv(out, file.path(OUTPUT_ROOT, '02_candidates', 'thematic_190_candidate_genes.csv'), row.names = FALSE)
write.table(
  data.frame(
    thematic_library_n = length(unique(toupper(library_genes[[gene_col]]))),
    thematic_overlap_candidates_n = nrow(out)
  ),
  file.path(OUTPUT_ROOT, '02_candidates', 'THEMATIC_LIBRARY_SUMMARY.tsv'),
  sep = '\t', row.names = FALSE, quote = FALSE
)
message('PASS: thematic candidate set generated.')
```
