## Supplementary material for "Transcriptomic and Machine-Learning Prioritization of OCEL1 in Low Bone Mineral Density With Multivariable cis-MR": Online Resource 2: Reproducibility Materials: 03_fit_ml_models.R

```
source(file.path(dirname(normalizePath(sub('^--file=', '', grep('^--file=', commandArgs(FALSE), value = TRUE)[1]), winslash = '/', mustWork = FALSE)), '00_paths.R'))
suppressPackageStartupMessages({library(e1071); library(glmnet); library(xgboost)})

# Fit the three machine-learning procedures using the frozen common feature
# universe defined from the two transcriptomic cohorts.
read_expr <- function(path) {
  d <- read.csv(path, row.names = 1, check.names = FALSE)
  m <- as.matrix(d)
  mode(m) <- 'numeric'
  rownames(m) <- toupper(rownames(m))
  m[!duplicated(rownames(m)), , drop = FALSE]
}

expr <- read_expr(file.path(DATA_ROOT, 'bulk', 'normalized_expression', 'GSE56815_expr_by_symbol.csv'))
validation_expr <- read_expr(file.path(DATA_ROOT, 'bulk', 'normalized_expression', 'GSE2208_expr_by_symbol.csv'))
group <- read.csv(file.path(DATA_ROOT, 'bulk', 'sample_metadata', 'GSE56815_group.csv'), stringsAsFactors = FALSE)
group$group <- tolower(trimws(group$group))
candidate_input <- read.csv(file.path(OUTPUT_ROOT, '02_candidates', 'thematic_190_candidate_genes.csv'), stringsAsFactors = FALSE)$gene
candidate_input <- unique(toupper(candidate_input))
candidate <- Reduce(intersect, list(candidate_input, rownames(expr), rownames(validation_expr)))
stopifnot(all(group$sample %in% colnames(expr)), all(group$group %in% c('high_bmd', 'low_bmd')), length(candidate) > 1)
X <- as.data.frame(t(as.matrix(expr[candidate, group$sample, drop = FALSE])), check.names = FALSE)
y <- factor(group$group, levels = c('high_bmd', 'low_bmd'))
out_dir <- file.path(OUTPUT_ROOT, '03_ml_fit')
dir.create(out_dir, recursive = TRUE, showWarnings = FALSE)
write.csv(data.frame(gene = candidate), file.path(out_dir, 'training_feature_universe.csv'), row.names = FALSE)
write.table(data.frame(
  discovery_candidate_n = length(candidate_input),
  discovery_measurable_n = sum(candidate_input %in% rownames(expr)),
  validation_measurable_n = sum(candidate_input %in% rownames(validation_expr)),
  common_train_validation_feature_n = length(candidate),
  validation_labels_used_for_training = FALSE
), file.path(out_dir, 'training_feature_universe_audit.tsv'), sep = '\t', quote = FALSE, row.names = FALSE)

# Fixed seeds and model settings for the canonical Paper 1 training stage.
svm_seed <- 20260326L
lasso_seed <- 8088L
xgb_seed <- 124L

fold_accuracy <- function(train_idx, test_idx, genes) {
  fit <- e1071::svm(x = X[train_idx, genes, drop = FALSE], y = y[train_idx], kernel = 'linear', scale = TRUE)
  mean(predict(fit, X[test_idx, genes, drop = FALSE]) == y[test_idx])
}

set.seed(svm_seed)
svm <- e1071::svm(x = X, y = y, kernel = 'linear', scale = TRUE)
svm_w <- as.vector(t(svm$coefs) %*% svm$SV)
names(svm_w) <- colnames(svm$SV)
svm_ranked <- names(sort(abs(svm_w), decreasing = TRUE))
svm_sizes <- seq(2L, min(40L, length(svm_ranked)), by = 1L)
svm_profile <- do.call(rbind, lapply(svm_sizes, function(n_keep) {
  genes <- head(svm_ranked, n_keep)
  accuracy <- unlist(lapply(seq_len(3L), function(rep_idx) {
    set.seed(svm_seed + rep_idx)
    folds <- sample(rep(seq_len(10L), length.out = nrow(X)))
    vapply(seq_len(10L), function(fold) fold_accuracy(which(folds != fold), which(folds == fold), genes), numeric(1))
  }))
  data.frame(Variables = n_keep, Accuracy = mean(accuracy, na.rm = TRUE), AccuracySD = sd(accuracy, na.rm = TRUE))
}))
write.csv(svm_profile, file.path(out_dir, 'svm_profile.csv'), row.names = FALSE)
write.csv(data.frame(gene = svm_ranked), file.path(out_dir, 'svm_ranked_genes.csv'), row.names = FALSE)
saveRDS(svm, file.path(out_dir, 'svm_linear_full_model.rds'))

set.seed(lasso_seed)
xmat <- as.matrix(X)
ynum <- as.integer(y == 'low_bmd')
lasso <- glmnet::cv.glmnet(xmat, ynum, alpha = 1, family = 'binomial', nfolds = 10, type.measure = 'class')
lasso_coef <- as.matrix(coef(lasso, s = 'lambda.1se'))
lasso_selected <- setdiff(rownames(lasso_coef)[as.numeric(lasso_coef[, 1]) != 0], '(Intercept)')
lambda_table <- data.frame(lambda = as.numeric(lasso$lambda), cvm = as.numeric(lasso$cvm), cvsd = as.numeric(lasso$cvsd))
lambda_table$nonzero <- vapply(lambda_table$lambda, function(lambda) sum(as.numeric(coef(lasso$glmnet.fit, s = lambda)) != 0) - 1L, numeric(1))
write.csv(lambda_table, file.path(out_dir, 'lasso_lambda_table.csv'), row.names = FALSE)
write.csv(data.frame(gene = lasso_selected), file.path(out_dir, 'lasso_lambda_1se_nonzero_genes.csv'), row.names = FALSE)
write.table(data.frame(lambda_min = lasso$lambda.min, lambda_1se = lasso$lambda.1se,
                       selected_lambda = lasso$lambda.1se, selected_nonzero_n = length(lasso_selected),
                       lasso_seed = lasso_seed, selection_rule = 'lambda.1se'),
            file.path(out_dir, 'LASSO_SELECTION_METADATA.tsv'), sep = '\t', quote = FALSE, row.names = FALSE)
coef_path <- as.matrix(lasso$glmnet.fit$beta)
lasso_path <- data.frame(gene = rep(rownames(coef_path), times = ncol(coef_path)),
                         lambda = rep(lasso$lambda, each = nrow(coef_path)),
                         coefficient = as.vector(coef_path), stringsAsFactors = FALSE)
lasso_path$selected_lambda <- abs(log(lasso_path$lambda) - log(lasso$lambda.1se)) < 1e-10
write.table(lasso_path, file.path(out_dir, 'LASSO_COEFFICIENT_PATH.tsv'), sep = '\t', quote = FALSE, row.names = FALSE)
saveRDS(lasso, file.path(out_dir, 'lasso_cv.rds'))

params <- list(objective = 'binary:logistic', eval_metric = 'auc', booster = 'gbtree', eta = 0.05,
               max_depth = 2, min_child_weight = 1, subsample = 1, colsample_bytree = 1, nthread = 1)
set.seed(xgb_seed)
dtrain <- xgboost::xgb.DMatrix(data = xmat, label = ynum)
xgb_cv <- xgboost::xgb.cv(params = params, data = dtrain, nrounds = 300, nfold = 5, stratified = TRUE, early_stopping_rounds = 20, verbose = 0)
xgb_nrounds <- xgb_cv$best_iteration
if (is.null(xgb_nrounds) || !is.finite(xgb_nrounds) || xgb_nrounds < 1) stop('XGBoost cross-validation did not return a valid best_iteration.')
set.seed(xgb_seed)
xgb <- xgboost::xgb.train(params = params, data = dtrain, nrounds = xgb_nrounds, verbose = 0)
xgb_importance <- xgboost::xgb.importance(model = xgb)
if (!nrow(xgb_importance) || !('Feature' %in% names(xgb_importance)) || !('Frequency' %in% names(xgb_importance))) stop('XGBoost produced no usable frequency importance table.')
xgb_importance <- xgb_importance[order(-xgb_importance$Frequency, seq_len(nrow(xgb_importance))), , drop = FALSE]
xgb_importance$frequency_share <- xgb_importance$Frequency / sum(xgb_importance$Frequency)
xgb_importance$cumulative_frequency <- cumsum(xgb_importance$frequency_share)
write.csv(xgb_importance, file.path(out_dir, 'xgb_frequency_importance.csv'), row.names = FALSE)
saveRDS(xgb, file.path(out_dir, 'xgb_model.rds'))

write.table(data.frame(candidate_n = length(candidate), svm_profile_repeats = 3L, svm_profile_folds = 10L,
                       svm_profile_sizes = paste(svm_sizes, collapse = ';'), svm_seed = svm_seed,
                       lasso_rule = 'lambda.1se', lasso_seed = lasso_seed, xgb_seed = xgb_seed,
                       xgb_booster = params$booster, xgb_best_nrounds = xgb_nrounds,
                       xgb_importance = 'frequency',
                       feature_universe = 'discovery_candidates_measurable_in_GSE56815_and_GSE2208',
                       validation_labels_used_for_training = FALSE), file.path(out_dir, 'ML_TRAINING_CONFIG.tsv'), sep = '\t', quote = FALSE, row.names = FALSE)
message('PASS: canonical Paper 1 ML training artifacts were written; selection remains stage 04.')
```
