## Supplementary material for "Transcriptomic and Machine-Learning Prioritization of OCEL1 in Low Bone Mineral Density With Multivariable cis-MR": Online Resource 2: Reproducibility Materials: 04_select_ml_consensus.R

```
source(file.path(dirname(normalizePath(sub('^--file=', '', grep('^--file=', commandArgs(FALSE), value = TRUE)[1]), winslash = '/', mustWork = FALSE)), '00_paths.R'))
fit_dir <- file.path(OUTPUT_ROOT, '03_ml_fit')
svm_profile <- read.csv(file.path(fit_dir, 'svm_profile.csv'), stringsAsFactors = FALSE)
svm_ranked <- read.csv(file.path(fit_dir, 'svm_ranked_genes.csv'), stringsAsFactors = FALSE)$gene
lasso_selected <- read.csv(file.path(fit_dir, 'lasso_lambda_1se_nonzero_genes.csv'), stringsAsFactors = FALSE)$gene
xgb <- read.csv(file.path(fit_dir, 'xgb_frequency_importance.csv'), stringsAsFactors = FALSE)

svm_profile$Variables <- as.integer(svm_profile$Variables)
svm_profile$Accuracy <- as.numeric(svm_profile$Accuracy)
svm_n <- min(svm_profile$Variables[svm_profile$Accuracy == max(svm_profile$Accuracy, na.rm = TRUE)])
if (length(svm_ranked) < svm_n) stop('SVM ranked gene output is shorter than the selected subset size.')
svm_selected <- toupper(svm_ranked[seq_len(svm_n)])

xgb$Frequency <- as.numeric(xgb$Frequency)
xgb$.input_order <- seq_len(nrow(xgb))
xgb <- xgb[order(-xgb$Frequency, xgb$.input_order, method = 'radix'), , drop = FALSE]
xgb$.input_order <- NULL
xgb$frequency_share <- xgb$Frequency / sum(xgb$Frequency)
xgb$cumulative_frequency <- cumsum(xgb$frequency_share)
xgb_n <- which(xgb$cumulative_frequency >= 0.5)[1]
if (is.na(xgb_n)) stop('XGBoost cumulative feature frequency did not reach the prespecified 0.50 threshold.')
xgb$selected_by_rule <- seq_len(nrow(xgb)) <= xgb_n
xgb_selected <- toupper(as.character(xgb$Feature[seq_len(xgb_n)]))

selection <- rbind(data.frame(method = 'Linear_SVM_weight_ranking_max_accuracy_tie', gene = svm_selected), data.frame(method = 'LASSO_lambda_1se', gene = toupper(lasso_selected)), data.frame(method = 'XGBoost_frequency_first_cumulative_ge_0.5', gene = xgb_selected))
strict <- sort(Reduce(intersect, split(selection$gene, selection$method)))
out_dir <- file.path(OUTPUT_ROOT, '04_ml_selection')
dir.create(out_dir, recursive = TRUE, showWarnings = FALSE)
write.csv(svm_profile, file.path(out_dir, 'svm_profile_with_rule.csv'), row.names = FALSE)
write.csv(xgb, file.path(out_dir, 'xgb_frequency_with_rule.csv'), row.names = FALSE)
write.csv(selection, file.path(out_dir, 'ml_selection_after_rules.csv'), row.names = FALSE)
write.csv(data.frame(gene = strict), file.path(out_dir, 'strict_intersection.csv'), row.names = FALSE)
counts <- table(factor(selection$method, levels = unique(selection$method)))
write.csv(data.frame(method = names(counts), n = as.integer(counts)), file.path(out_dir, 'ml_selection_counts.csv'), row.names = FALSE)
message('PASS: ML selection rules and strict intersection were applied.')
```
