## Supplementary material for "Transcriptomic and Machine-Learning Prioritization of OCEL1 in Low Bone Mineral Density With Multivariable cis-MR": Online Resource 2: Reproducibility Materials: 05_validate_selected_intersection.R

```
source(file.path(dirname(normalizePath(sub('^--file=', '', grep('^--file=', commandArgs(FALSE), value = TRUE)[1]), winslash = '/', mustWork = FALSE)), '00_paths.R'))
suppressPackageStartupMessages(library(pROC))
read_data <- function(dataset) {
  expr <- read.csv(file.path(DATA_ROOT, 'bulk', 'normalized_expression', paste0(dataset, '_expr_by_symbol.csv')), row.names = 1, check.names = FALSE)
  grp <- read.csv(file.path(DATA_ROOT, 'bulk', 'sample_metadata', paste0(dataset, '_group.csv')), stringsAsFactors = FALSE)
  list(expr = as.matrix(expr[, grp$sample, drop = FALSE]), group = grp$group)
}
score_one <- function(x, group) {
  y <- factor(group, levels = c('high_bmd', 'low_bmd'))
  delta <- mean(x[y == 'low_bmd']) - mean(x[y == 'high_bmd'])
  p <- wilcox.test(x[y == 'low_bmd'], x[y == 'high_bmd'])$p.value
  auc <- as.numeric(pROC::auc(pROC::roc(y, x, levels = c('high_bmd', 'low_bmd'), direction = 'auto', quiet = TRUE)))
  c(auc = max(auc, 1 - auc), delta = delta, p = p)
}
genes <- read.csv(file.path(OUTPUT_ROOT, '04_ml_selection', 'strict_intersection.csv'), stringsAsFactors = FALSE)$gene
tr <- read_data('GSE56815'); va <- read_data('GSE2208')
out <- do.call(rbind, lapply(genes, function(g) {
  available_train <- g %in% rownames(tr$expr)
  available_validation <- g %in% rownames(va$expr)
  if (!available_train || !available_validation) {
    return(data.frame(gene = g, available_train = available_train, available_validation = available_validation,
                      train_auc = NA_real_, primary_auc = NA_real_, train_delta = NA_real_, primary_delta = NA_real_,
                      train_p = NA_real_, primary_p = NA_real_, auc_gate = FALSE, direction_concordant = FALSE,
                      expression_p_gate = FALSE, final_gate = FALSE, validation_status = 'NOT_MEASURABLE_IN_BOTH_COHORTS'))
  }
  a <- score_one(tr$expr[g, ], tr$group); b <- score_one(va$expr[g, ], va$group)
  direction_concordant <- sign(a['delta']) == sign(b['delta']) && a['delta'] != 0 && b['delta'] != 0
  auc_gate <- a['auc'] >= .60 && b['auc'] >= .60
  expression_p_gate <- a['p'] < .05 && b['p'] < .05
  data.frame(gene = g, available_train = available_train, available_validation = available_validation,
             train_auc = a['auc'], primary_auc = b['auc'], train_delta = a['delta'], primary_delta = b['delta'],
             train_p = a['p'], primary_p = b['p'], auc_gate = auc_gate, direction_concordant = direction_concordant,
             expression_p_gate = expression_p_gate, final_gate = auc_gate && direction_concordant && expression_p_gate,
             validation_status = 'MEASURABLE_IN_BOTH_COHORTS')
}))
dir.create(file.path(OUTPUT_ROOT, '05_validation'), recursive = TRUE, showWarnings = FALSE)
write.csv(out, file.path(OUTPUT_ROOT, '05_validation', 'selected_intersection_two_cohort_metrics.csv'), row.names = FALSE)
write.csv(out[out$final_gate %in% TRUE, , drop = FALSE], file.path(OUTPUT_ROOT, '05_validation', 'FINAL_VALIDATED_GENES.csv'), row.names = FALSE)
message('PASS: two-cohort validation metrics were generated from the selected intersection.')
```
