## Supplementary material for "Transcriptomic and Machine-Learning Prioritization of OCEL1 in Low Bone Mineral Density With Multivariable cis-MR": Online Resource 2: Reproducibility Materials: 00_prepare_frozen_component_inputs.py

```
"""Materialize the explicit Paper 1 local-cis frozen inputs from the accepted grid.

The accepted 154-row grid is used only to materialize the two declared inputs.
The downstream generator reads those inputs rather than a hard-coded transcript list.
"""
from __future__ import annotations

import argparse
from pathlib import Path

import pandas as pd


def main() -> None:
    parser = argparse.ArgumentParser()
    parser.add_argument("--source", type=Path, required=True)
    parser.add_argument("--outdir", type=Path, required=True)
    args = parser.parse_args()
    grid = pd.read_csv(args.source, sep="\t")
    required = {"gene", "OCEL1_focal_SNP"}
    missing = required.difference(grid.columns)
    if missing:
        raise ValueError(f"Missing columns: {sorted(missing)}")
    genes = pd.DataFrame({"gene": sorted(grid["gene"].dropna().astype(str).str.upper().unique())})
    components = pd.DataFrame({"OCEL1_SNP": grid["OCEL1_focal_SNP"].dropna().astype(str).str.upper().drop_duplicates()})
    if len(grid) != 154 or len(genes) != 14 or len(components) != 11:
        raise ValueError("Expected a 14 x 11 frozen component-sharing grid.")
    args.outdir.mkdir(parents=True, exist_ok=True)
    genes.to_csv(args.outdir / "local_cis_candidate_transcripts.tsv", sep="\t", index=False)
    components.to_csv(args.outdir / "ocel1_focal_components.tsv", sep="\t", index=False)


if __name__ == "__main__":
    main()
```
