## Supplementary material for "Transcriptomic and Machine-Learning Prioritization of OCEL1 in Low Bone Mineral Density With Multivariable cis-MR": Online Resource 2: Reproducibility Materials: 01_build_component_sharing.py

```
"""Generate the Paper 1 local cis component-sharing analysis.

Inputs are an eQTLGen cis summary file, allele frequencies, and the
UKBB337K chr19 LD index and sparse matrix.  No outcome or donor-level data
are used.  All input locations are explicit environment variables so this
script can be rerun against public-source material without an installation-
specific path.
"""
from __future__ import annotations

import csv, gzip, hashlib, json, math, os, sys
from pathlib import Path
import numpy as np
import pandas as pd
from scipy import sparse
from scipy.stats import norm as normal_dist
try:
    from cojopy.cojopy import COJO as CojoModel
except Exception as exc:
    raise RuntimeError("cojopy is required for the conditional component selector") from exc

def required_path(name: str) -> Path:
    value = os.environ.get(name)
    if not value:
        raise RuntimeError(f"Set {name} to the required input or output path.")
    path = Path(value).expanduser().resolve()
    if name != "COMPONENT_OUTPUT_ROOT" and not path.exists():
        raise FileNotFoundError(f"{name} does not exist: {path}")
    return path

SOURCE = required_path("EQTLGEN_CIS_SOURCE")
AF = required_path("EQTLGEN_ALLELE_FREQUENCY_SOURCE")
LD_NPZ = required_path("UKBB337K_CHR19_LD_NPZ")
LD_INDEX = required_path("UKBB337K_CHR19_LD_INDEX")
CANDIDATE_TRANSCRIPTS = required_path("LOCAL_CIS_CANDIDATE_TRANSCRIPTS")
FOCAL_COMPONENTS = required_path("OCEL1_FOCAL_COMPONENTS")
OUT = required_path("COMPONENT_OUTPUT_ROOT")
RAW = OUT / "cache" / "candidate_raw_cis"
RES = OUT / "results"
LOG = OUT / "logs"
for d in (RAW, RES, LOG): d.mkdir(parents=True, exist_ok=True)

MARGINAL_P = 1.829e-5
CONDITIONAL_P = 1e-3
COJO_COLLINEAR_R2 = 0.9
MATCH_R2_PRIMARY = 0.8
MATCH_R2_SENS = 0.5
COMPLEMENT = str.maketrans("ACGT", "TGCA")

def snp(x): return str(x).strip().upper()
def allele(x): return str(x).strip().upper()
def comp(a): return allele(a).translate(COMPLEMENT)
def pnum(x):
    try: return float(x)
    except Exception: return np.nan
def sha256(path):
    h=hashlib.sha256()
    with open(path,"rb") as f:
        for b in iter(lambda:f.read(1<<20),b""): h.update(b)
    return h.hexdigest()

def load_af():
    out={}
    path = AF
    with (gzip.open(path,"rt",newline="") if path.suffix==".gz" else open(path,"r",newline="")) as fh:
        reader=csv.DictReader(fh, delimiter="\t")
        for r in reader:
            s=snp(r.get("SNP", r.get("rsid", r.get("variant_id", ""))))
            if not s: continue
            freq=pnum(r.get("freq", r.get("AF", r.get("AlleleB_all", ""))))
            if np.isfinite(freq) and 0 < freq < 1: out[s]=freq
    return out, str(path)

def load_ld_index():
    with gzip.open(LD_INDEX,"rt") as f: return pd.read_csv(f, sep="\t")

def load_frozen_structure():
    """Load the explicit frozen 14-transcript and 11-component input files."""
    candidate_frame = pd.read_csv(CANDIDATE_TRANSCRIPTS, sep="\t")
    component_frame = pd.read_csv(FOCAL_COMPONENTS, sep="\t")
    if "gene" not in candidate_frame or "OCEL1_SNP" not in component_frame:
        raise ValueError("Frozen input columns must be gene and OCEL1_SNP.")
    candidates = candidate_frame["gene"].dropna().astype(str).str.upper().tolist()
    focal = [snp(value) for value in component_frame["OCEL1_SNP"].dropna().tolist()]
    if len(candidates) != 14 or len(set(candidates)) != 14 or len(focal) != 11 or len(set(focal)) != 11:
        raise ValueError("Frozen local-cis inputs must contain 14 unique transcripts and 11 unique components.")
    return candidates, focal

def extract_raw(all_genes):
    existing=[]
    for g in all_genes:
        p=RAW/f"{g}_eqtlgen_raw_cis.tsv.gz"
        if p.exists() and p.stat().st_size>100: existing.append(g)
    if len(existing)==len(all_genes): return {g:RAW/f"{g}_eqtlgen_raw_cis.tsv.gz" for g in all_genes}
    handles={g:gzip.open(RAW/f"{g}_eqtlgen_raw_cis.tsv.gz","wt",newline="") for g in all_genes if g not in existing}
    writers={}; header=None; counts={g:0 for g in all_genes}
    try:
        with gzip.open(SOURCE,"rt",newline="") as fh:
            reader=csv.DictReader(fh, delimiter="\t")
            header=reader.fieldnames
            for g,h in handles.items(): writers[g]=csv.DictWriter(h,fieldnames=header,delimiter="\t"); writers[g].writeheader()
            for r in reader:
                g=str(r.get("GeneSymbol","")).strip()
                if g in writers: writers[g].writerow(r); counts[g]+=1
    finally:
        for h in handles.values(): h.close()
    (LOG/"extraction_counts.tsv").write_text("gene\tn_rows\n"+"\n".join(f"{g}\t{counts[g]}" for g in all_genes), encoding="utf-8")
    return {g:RAW/f"{g}_eqtlgen_raw_cis.tsv.gz" for g in all_genes}

def harmonize(df, ld, afmap):
    idx={snp(x):i for i,x in enumerate(ld["rsid"])}
    rows=[]
    for _,r in df.iterrows():
        s=snp(r.get("SNP")); i=idx.get(s)
        if i is None: continue
        ea,oa=allele(r.get("AssessedAllele")),allele(r.get("OtherAllele"))
        la,lb=allele(ld.iloc[i]["allele1"]),allele(ld.iloc[i]["allele2"])
        orient=None
        if {ea,oa}=={la,lb}: orient=1 if ea==la else -1
        elif {comp(ea),comp(oa)}=={la,lb}: orient=1 if comp(ea)==la else -1
        if orient is None: continue
        z=float(pnum(r.get("Zscore"))); n=float(pnum(r.get("NrSamples"))); p=float(pnum(r.get("Pvalue")))
        eaf=float(afmap.get(s, np.nan))
        if not (np.isfinite(z) and np.isfinite(n) and np.isfinite(eaf)): continue
        se=1/math.sqrt(2*eaf*(1-eaf)*(n+z*z)); beta=z*se*orient; z_signed=z*orient
        rows.append({"SNP":s,"ld_index":int(i),"Pvalue":p,"Zscore":z_signed,"beta":beta,"SE":se,"EAF":eaf,"EA":ea,"OA":oa,"ld_A1":la,"ld_A2":lb,"orientation":orient,"SNPChr":r.get("SNPChr"),"SNPPos":r.get("SNPPos"),"NrSamples":n})
    return pd.DataFrame(rows).drop_duplicates("SNP")

def cond_select(dat, R, label="gene"):
    """COJO-style forward conditional selection using LD and Z statistics."""
    if dat.empty: return dat.assign(selection_status="NO_VALID_INPUT")
    work=dat.reset_index(drop=True).copy(); z=work.Zscore.to_numpy(float); selected=[]; status=np.array(["NOT_SELECTED"]*len(work),dtype=object); condp=np.full(len(work),np.nan); condz=np.full(len(work),np.nan)
    d = OUT / "cache" / "cojopy_runtime"; d.mkdir(parents=True, exist_ok=True)
    ma = d / f"{label}.ma"; ld = d / f"{label}.ld.txt"
    q = work[["SNP","EA","OA","EAF","beta","SE","Pvalue","NrSamples"]].rename(columns={"EA":"A1","OA":"A2","EAF":"freq","beta":"b","SE":"se","Pvalue":"p","NrSamples":"N"})
    q.to_csv(ma, sep="\t", index=False); np.savetxt(ld, R, fmt="%.9g", delimiter="\t")
    model = CojoModel(p_cutoff=CONDITIONAL_P, collinear_cutoff=COJO_COLLINEAR_R2, window_size=10_000_000, maf_cutoff=0.01, diff_freq_cutoff=0.2)
    model.load_sumstats(str(ma), str(ld)); selected_df = model.conditional_selection()
    ss=set(selected_df["SNP"].map(snp)) if not selected_df.empty else set()
    status=np.where(work.SNP.isin(ss), "INDEPENDENT_CONDITIONAL_SIGNAL", "NOT_SELECTED")
    if not selected_df.empty:
        for _,row in selected_df.iterrows():
            i=work.index[work.SNP.eq(snp(row["SNP"]))]
            if len(i):
                ii=int(i[0]); b=float(row.get("joint_beta", np.nan)); se=float(row.get("joint_se", np.nan)); condz[ii]=b/se if np.isfinite(se) and se>0 else np.nan; condp[ii]=float(row.get("joint_p", np.nan))
    work["conditional_Z"]=condz; work["conditional_P"]=condp; work["selection_status"]=status; work["signal_order"]=np.nan
    for k,i in enumerate(np.where(status=="INDEPENDENT_CONDITIONAL_SIGNAL")[0],1): work.loc[i,"signal_order"]=k
    work["conditional_method"]="cojopy_1.0.0_slct"
    return work

def main():
    candidates, frozen_components = load_frozen_structure()
    all_genes = ["OCEL1"] + candidates
    paths=extract_raw(all_genes); afmap,afsrc=load_af(); ld=load_ld_index(); csr=sparse.load_npz(LD_NPZ).tocsr()
    ocel1_comp=frozen_components; idx={snp(x):i for i,x in enumerate(ld["rsid"])}
    oidx=[idx[s] for s in ocel1_comp if s in idx]
    summaries=[]; components=[]; pergene=[]; conditional_by_gene={}
    for g in all_genes:
        raw=pd.read_csv(paths[g],sep="\t",compression="infer")
        raw=raw[pd.to_numeric(raw["Pvalue"],errors="coerce")<MARGINAL_P].copy(); raw_n=len(raw)
        h=harmonize(raw,ld,afmap)
        if h.empty:
            summaries.append({"gene":g,"n_raw_rows":len(raw),"n_usable":0,"n_conditional":0,"status":"NO_USABLE_LD_AF_INPUT"}); continue
        ids=h.ld_index.astype(int).tolist(); sub=csr[ids,:][:,ids].tocsr(); R=(sub+sub.T).tocsr(); R.setdiag(1.0); R.eliminate_zeros()
        sel=cond_select(h,R,g); sel.to_csv(RES/f"{g}_conditional_architecture.tsv",sep="\t",index=False)
        conditional_by_gene[g] = sel
        chosen=sel[sel.selection_status=="INDEPENDENT_CONDITIONAL_SIGNAL"].copy(); chosen["gene"]=g; components.append(chosen)
        # compare each OCEL1 component to the candidate's selected components by max LD r2
        if len(chosen):
            # Candidate .ld_index is local to its Run44 cache matrix; map by
            # SNP identity before querying the shared UKBB337K LD reference.
            cids=[idx[s] for s in chosen.SNP if s in idx]
            cr=csr[oidx,:][:,cids].toarray(); maxr2=np.max(cr*cr,axis=1) if cr.size else np.zeros(len(oidx))
            n08=int(np.sum(maxr2>=MATCH_R2_PRIMARY)); n05=int(np.sum(maxr2>=MATCH_R2_SENS)); meanr=float(np.mean(maxr2)) if len(maxr2) else np.nan; maxr=float(np.max(maxr2)) if len(maxr2) else np.nan
            for s,m in zip(ocel1_comp,maxr2): pergene.append({"gene":g,"OCEL1_SNP":s,"max_component_r2":m,"shared_r2_ge_0.8":m>=.8,"shared_r2_ge_0.5":m>=.5})
        else: n08=n05=0; meanr=maxr=np.nan
        summaries.append({"gene":g,"n_raw_rows":raw_n,"n_usable":len(h),"n_conditional":len(chosen),"cx14_shared_components_r2_ge_0.8":n08,"cx14_shared_components_r2_ge_0.5":n05,"mean_max_component_r2":meanr,"max_component_r2":maxr,"status":"ANALYZED"})
    summary_df=pd.DataFrame(summaries)
    summary_df[summary_df.gene.isin(candidates)].to_csv(RES/"candidate_architecture_summary.tsv",sep="\t",index=False)
    summary_df.to_csv(RES/"all_gene_architecture_summary.tsv",sep="\t",index=False)
    if components: pd.concat(components,ignore_index=True).to_csv(RES/"all_candidates_conditional_components.tsv",sep="\t",index=False)
    pd.DataFrame(pergene).to_csv(RES/"OCEL1_CX14_candidate_component_sharing.tsv",sep="\t",index=False)
    # pairwise TWMR-style Z correlations on shared significant, independent panel SNPs
    allsel=pd.concat(components,ignore_index=True) if components else pd.DataFrame()
    pair=[]
    for g in candidates:
        if allsel.empty or "OCEL1" not in conditional_by_gene: continue
        a=allsel[allsel.gene==g]; oc=pd.read_csv(RES/"OCEL1_conditional_architecture.tsv",sep="\t") if (RES/"OCEL1_conditional_architecture.tsv").exists() else None
        if oc is None: continue
        oc=oc[oc.SNP.isin(ocel1_comp)].copy()
        m=a.merge(oc,on="SNP",suffixes=("_g","_oc")); m=m[(m.Pvalue_g<MARGINAL_P)&(m.Pvalue_oc<MARGINAL_P)]
        r=float(m.Zscore_g.corr(m.Zscore_oc)) if len(m)>=3 else np.nan
        pair.append({"gene1":g,"gene2":"OCEL1","n_shared_significant_independent_snps":len(m),"z_correlation":r,"z_correlation_r2":r*r if np.isfinite(r) else np.nan,"status":"ASSESSABLE" if len(m)>=3 else "COLLINEARITY_NOT_ASSESSABLE"})
    pd.DataFrame(pair).to_csv(RES/"pairwise_twmr_collinearity.tsv",sep="\t",index=False)
    meta={"source":str(SOURCE),"source_sha256":sha256(SOURCE),"af_source":afsrc,"ld_npz":str(LD_NPZ),"ld_sha256":sha256(LD_NPZ),"candidate_transcripts":str(CANDIDATE_TRANSCRIPTS),"candidate_transcripts_sha256":sha256(CANDIDATE_TRANSCRIPTS),"focal_components":str(FOCAL_COMPONENTS),"focal_components_sha256":sha256(FOCAL_COMPONENTS),"marginal_p":MARGINAL_P,"conditional_p":CONDITIONAL_P,"candidate_count":len(candidates),"focal_component_count":len(ocel1_comp),"selector":"cojopy_1.0.0_slct","oneK1K_used":False,"outcome_used":False}
    (RES/"analysis_manifest.json").write_text(json.dumps(meta,indent=2),encoding="utf-8")

if __name__=="__main__": main()
```
