## Supplementary material for "Transcriptomic and Machine-Learning Prioritization of OCEL1 in Low Bone Mineral Density With Multivariable cis-MR": Online Resource 2: Reproducibility Materials: 06_build_primary4_panel.py

```
"""Build the Paper 1 four-exposure panel from declared public source files.

The outcome is opened only after the exposure-side candidate panel has been
defined.  All output is written below ``outputs/06_primary4_panel``.
"""
from __future__ import annotations

import csv
import gzip
import math
from pathlib import Path

import numpy as np
import pandas as pd
from scipy import sparse


PROJECT = Path(__file__).resolve().parents[2]
CLEAN = Path(__file__).resolve().parents[1]
DATA = PROJECT / "00_DATA_PAPER1" / "mr" / "source"
OUT = CLEAN / "outputs" / "06_primary4_panel"
COMPONENT_SHARING_SOURCE = PROJECT / "00_DATA_PAPER1" / "mr" / "competitor_tables" / "OCEL1_new_focal_component_sharing.tsv"
GENES = ("OCEL1", "NR2F6", "MRPL34", "BABAM1")
P_CUTOFF = 2e-5
MAF_CUTOFF = 0.01
LD_R2 = 0.64
WINDOW_BP = 1_000_000
HEEL_N = 426_824.0


def norm(value: object) -> str:
    return str(value).strip().upper()


def comp(value: object) -> str:
    return norm(value).translate(str.maketrans("ACGT", "TGCA"))


def palindromic(a1: object, a2: object) -> bool:
    return {norm(a1), norm(a2)} in ({"A", "T"}, {"C", "G"})


def orient(ea: object, oa: object, a1: object, a2: object) -> tuple[float, str]:
    """Return sign from source effect allele to the UKBB reference A1 allele."""
    ea, oa, a1, a2 = map(norm, (ea, oa, a1, a2))
    isp = palindromic(a1, a2)
    if (ea, oa) == (a1, a2):
        return 1.0, "PAL_EXACT" if isp else "EXACT"
    if (ea, oa) == (a2, a1):
        return -1.0, "PAL_SWAP" if isp else "SWAP"
    if isp:
        return np.nan, "PAL_ORIENTATION_AMBIGUOUS"
    if (comp(ea), comp(oa)) == (a1, a2):
        return 1.0, "COMPLEMENT_EXACT"
    if (comp(ea), comp(oa)) == (a2, a1):
        return -1.0, "COMPLEMENT_SWAP"
    return np.nan, "ALLELE_INCOMPATIBLE"


def read_eqtl() -> dict[str, dict[str, dict[str, object]]]:
    source = DATA / "eqtlgen" / "cis_eqtlgen_phase1.tsv.gz"
    found: dict[str, dict[str, dict[str, object]]] = {gene: {} for gene in GENES}
    with gzip.open(source, "rt", encoding="utf-8-sig", newline="") as handle:
        for row in csv.DictReader(handle, delimiter="\t"):
            gene = norm(row.get("GeneSymbol", ""))
            if gene not in found:
                continue
            try:
                record = {
                    "SNP_norm": norm(row["SNP"]), "position_eqtl": int(float(row["SNPPos"])),
                    "EA": norm(row["AssessedAllele"]), "OA": norm(row["OtherAllele"]),
                    "Z": float(row["Zscore"]), "N": float(row["NrSamples"]),
                    "P": float(row["Pvalue"]), "FDR": float(row["FDR"]),
                }
            except (KeyError, TypeError, ValueError):
                continue
            old = found[gene].get(record["SNP_norm"])
            if old is None or float(record["P"]) < float(old["P"]):
                found[gene][record["SNP_norm"]] = record
    return found


def read_variant_index() -> pd.DataFrame:
    index = pd.read_csv(DATA / "ukbb337k" / "chr19_variant_index.tsv", sep="\t", dtype=str)
    index["SNP_norm"] = index["rsid_norm"].map(norm)
    index["ukbb_index"] = pd.to_numeric(index["ukbb_index"], errors="raise").astype(int)
    index["position"] = pd.to_numeric(index["position"], errors="raise").astype(int)
    index["A1"] = index["allele1"].map(norm)
    index["A2"] = index["allele2"].map(norm)
    return index.drop_duplicates("SNP_norm").set_index("SNP_norm")


def read_af(targets: set[str]) -> dict[str, tuple[str, str, float]]:
    found = {}
    with gzip.open(DATA / "eqtlgen" / "allele_frequency.tsv.gz", "rt", encoding="utf-8-sig", newline="") as handle:
        for row in csv.DictReader(handle, delimiter="\t"):
            snp = norm(row.get("SNP", ""))
            if snp not in targets:
                continue
            try:
                found[snp] = (norm(row["AlleleA"]), norm(row["AlleleB"]), float(row["AlleleB_all"]))
            except (KeyError, TypeError, ValueError):
                pass
    return found


def read_heel(targets: set[str]) -> dict[str, dict[str, str]]:
    found = {}
    with gzip.open(DATA / "heel" / "GCST006979.h.tsv.gz", "rt", encoding="utf-8-sig", newline="") as handle:
        for row in csv.DictReader(handle, delimiter="\t"):
            snp = norm(row.get("hm_rsid", ""))
            if snp in targets:
                found[snp] = row
    return found


def load_ld() -> sparse.csr_matrix:
    matrix = sparse.load_npz(DATA / "ukbb337k" / "chr19_16000001_19000001.npz").tocsr()
    matrix = (matrix + matrix.T).tocsr()
    matrix.setdiag(1.0)
    return matrix


def build_exposure_candidates(rows: dict[str, dict[str, dict[str, object]]], index: pd.DataFrame) -> pd.DataFrame:
    records = []
    for snp in sorted(set().union(*(set(rows[g]) for g in GENES))):
        values = {f"P_{g}": rows[g].get(snp, {}).get("P", np.nan) for g in GENES}
        values.update({f"FDR_{g}": rows[g].get(snp, {}).get("FDR", np.nan) for g in GENES})
        record: dict[str, object] = {"SNP_norm": snp, **values}
        record["min_P_all_exposures"] = np.nanmin([values[f"P_{g}"] for g in GENES])
        record["eligible_min_P_lt_2e_5"] = record["min_P_all_exposures"] < P_CUTOFF
        if snp in index.index:
            record.update(index.loc[snp, ["ukbb_index", "position", "A1", "A2"]].to_dict())
            record["UKBB_match"] = True
        else:
            record.update({"ukbb_index": np.nan, "position": np.nan, "A1": np.nan, "A2": np.nan, "UKBB_match": False})
        records.append(record)
    return pd.DataFrame(records)


def build_complete_candidates(pool: pd.DataFrame, rows: dict[str, dict[str, dict[str, object]]], af: dict[str, tuple[str, str, float]]) -> tuple[pd.DataFrame, pd.DataFrame]:
    kept, audit = [], []
    for _, row in pool.iterrows():
        item, reasons = row.to_dict(), []
        frequency = af.get(norm(row.SNP_norm))
        if frequency is None or not 0 < frequency[2] < 1:
            reasons.append("INVALID_OR_MISSING_EQTL_AF")
        for gene in GENES:
            source = rows[gene].get(norm(row.SNP_norm))
            if source is None:
                reasons.append(f"MISSING_FULL_CIS_{gene}")
                continue
            sign, status = orient(source["EA"], source["OA"], row.A1, row.A2)
            item[f"orientation_{gene}"] = status
            if not np.isfinite(sign):
                reasons.append(f"{gene}_{status}")
                continue
            item[f"Z_{gene}"] = sign * float(source["Z"])
            item[f"N_{gene}"] = float(source["N"])
            item[f"b_{gene}"] = item[f"Z_{gene}"] / math.sqrt(item[f"N_{gene}"])
            item[f"se_{gene}"] = 1 / math.sqrt(item[f"N_{gene}"])
        item["exposure_status"] = "PASS" if not reasons else "|".join(sorted(set(reasons)))
        audit.append(item)
        if not reasons:
            kept.append(item)
    return pd.DataFrame(kept), pd.DataFrame(audit)


def add_outcome(exposure_panel: pd.DataFrame, heel: dict[str, dict[str, str]]) -> tuple[pd.DataFrame, pd.DataFrame]:
    kept, audit = [], []
    for _, row in exposure_panel.iterrows():
        item, reasons = row.to_dict(), []
        source = heel.get(norm(row.SNP_norm))
        if source is None:
            reasons.append("HEEL_NOT_FOUND")
        else:
            sign, status = orient(source.get("hm_effect_allele", ""), source.get("hm_other_allele", ""), row.A1, row.A2)
            item["heel_orientation"] = status
            try:
                beta, se, eaf = float(source["hm_beta"]) * sign, float(source["standard_error"]), float(source["hm_effect_allele_frequency"])
            except (KeyError, TypeError, ValueError):
                beta, se, eaf = np.nan, np.nan, np.nan
            if not np.isfinite(sign): reasons.append(f"HEEL_{status}")
            elif not (np.isfinite(se) and se > 0 and np.isfinite(eaf) and 0 < eaf < 1): reasons.append("HEEL_INVALID_EFFECT_SE_OR_EAF")
            else:
                item.update({"b_heel": beta, "se_heel": se, "Z_heel": beta / se, "N_heel": HEEL_N,
                             "b_heel_std": beta / se / math.sqrt(HEEL_N), "se_heel_std": 1 / math.sqrt(HEEL_N),
                             "heel_effect_allele": norm(source["hm_effect_allele"]), "heel_other_allele": norm(source["hm_other_allele"]),
                             "heel_EAF": eaf, "MAF_heel": min(eaf, 1 - eaf)})
        item["outcome_status"] = "PASS" if not reasons else "|".join(sorted(set(reasons)))
        audit.append(item)
        if not reasons and item["MAF_heel"] >= MAF_CUTOFF:
            kept.append(item)
    return pd.DataFrame(kept), pd.DataFrame(audit)


def greedy_prune(panel: pd.DataFrame, ld: sparse.csr_matrix) -> tuple[pd.DataFrame, pd.DataFrame]:
    table = panel.sort_values(["min_P_all_exposures", "position", "SNP_norm"]).reset_index(drop=True).copy()
    ids, pos = table.ukbb_index.astype(int).to_numpy(), table.position.astype(int).to_numpy()
    blocked, selected, blocker = np.zeros(len(table), bool), [], np.full(len(table), "", dtype=object)
    for i in range(len(table)):
        if blocked[i]: continue
        selected.append(i)
        nearby = np.flatnonzero(np.abs(pos - pos[i]) <= WINDOW_BP)
        corr = ld[ids[i], ids[nearby]].toarray().ravel()
        hits = nearby[np.square(corr) >= LD_R2]
        for j in hits:
            if j != i and not blocked[j]: blocker[j] = str(table.loc[i, "SNP_norm"])
        blocked[hits] = True
    table["selection_status"] = "EXCLUDED_BY_GREEDY_LD"
    table.loc[selected, "selection_status"] = "SELECTED"
    table["blocked_by_SNP"] = blocker
    final = table.iloc[selected].copy().reset_index(drop=True)
    final["selection_order"] = np.arange(1, len(final) + 1)
    return final, table


def main() -> None:
    OUT.mkdir(parents=True, exist_ok=True)
    rows, index = read_eqtl(), read_variant_index()
    pool = build_exposure_candidates(rows, index)
    pool.to_csv(OUT / "01_exposure_candidate_union.tsv", sep="\t", index=False)
    eligible = pool[pool.eligible_min_P_lt_2e_5 & pool.UKBB_match].copy()
    complete, exposure_audit = build_complete_candidates(eligible, rows, read_af(set(eligible.SNP_norm)))
    exposure_audit.to_csv(OUT / "02_exposure_allele_af_qc.tsv", sep="\t", index=False)
    preld, outcome_audit = add_outcome(complete, read_heel(set(complete.SNP_norm)))
    outcome_audit.to_csv(OUT / "03_outcome_technical_maf_qc.tsv", sep="\t", index=False)
    preld["driving_exposure"] = preld.apply(lambda r: min(GENES, key=lambda g: r[f"P_{g}"]), axis=1)
    final, prune_audit = greedy_prune(preld, load_ld())
    prune_audit.to_csv(OUT / "04_ld_pruning_audit.tsv", sep="\t", index=False)
    final.to_csv(OUT / "PRIMARY_268_PANEL.tsv", sep="\t", index=False)
    ids = final.ukbb_index.astype(int).to_numpy()
    ld = load_ld()[ids, :][:, ids].toarray()
    pd.DataFrame(ld, index=final.SNP_norm, columns=final.SNP_norm).to_csv(OUT / "PRIMARY_268_RAW_LD.tsv", sep="\t")
    # This is a construction sensitivity: palindromic variants are removed
    # before greedy LD pruning, so alternatives previously blocked by a
    # palindromic lead may enter the no-palindromic panel.
    no_pal, no_pal_audit = greedy_prune(preld[~preld.apply(lambda r: palindromic(r.A1, r.A2), axis=1)].copy(), load_ld())
    no_pal_audit.to_csv(OUT / "05_no_pal_ld_pruning_audit.tsv", sep="\t", index=False)
    no_pal.to_csv(OUT / "PRIMARY_263_NO_PAL_PANEL.tsv", sep="\t", index=False)
    ii = no_pal.ukbb_index.astype(int).to_numpy()
    pd.DataFrame(load_ld()[ii, :][:, ii].toarray(), index=no_pal.SNP_norm, columns=no_pal.SNP_norm).to_csv(OUT / "PRIMARY_263_NO_PAL_RAW_LD.tsv", sep="\t")
    pal_rows = []
    af = read_af(set(final.SNP_norm))
    for _, row in final[final.apply(lambda r: palindromic(r.A1, r.A2), axis=1)].iterrows():
        aa, ab, fb = af.get(row.SNP_norm, (np.nan, np.nan, np.nan))
        eqtl_a1 = fb if row.A1 == ab else (1 - fb if row.A1 == aa else np.nan)
        heel_a1 = row.heel_EAF if row.heel_orientation == "PAL_EXACT" else 1 - row.heel_EAF
        record = {"SNP_norm": row.SNP_norm, "A1": row.A1, "A2": row.A2, "heel_effect_allele": row.heel_effect_allele, "heel_other_allele": row.heel_other_allele, "heel_orientation": row.heel_orientation, "eqtl_alleleA": aa, "eqtl_alleleB": ab, "eqtl_frequency_A1": eqtl_a1, "heel_frequency_A1": heel_a1, "frequency_absolute_difference": abs(eqtl_a1 - heel_a1) if np.isfinite(eqtl_a1) else np.nan, "frequency_concordance_status": "CONCORDANT_LE_0.10" if np.isfinite(eqtl_a1) and abs(eqtl_a1 - heel_a1) <= .10 else "NOT_ASSESSABLE_OR_DISCORDANT", "retained": True}
        for gene in GENES:
            source = rows[gene][row.SNP_norm]
            record[f"{gene}_EA"] = source["EA"]; record[f"{gene}_OA"] = source["OA"]; record[f"{gene}_orientation"] = row[f"orientation_{gene}"]
        pal_rows.append(record)
    pd.DataFrame(pal_rows).to_csv(OUT / "PALINDROMIC_VARIANT_AUDIT.tsv", sep="\t", index=False)
    driving = final.driving_exposure.value_counts()
    summary = {"candidate_union_n": len(pool), "exposure_eligible_ukbb_n": len(eligible), "exposure_allele_af_usable_n": len(complete), "heel_technically_usable_n": int((outcome_audit.outcome_status == 'PASS').sum()), "maf_ge_0_01_n": len(preld), "primary_panel_n": len(final), "no_pal_panel_n": len(no_pal), "outcome_effect_used_before_selection": False, **{f"driving_{g}_n": int(driving.get(g, 0)) for g in GENES}}
    pd.DataFrame([summary]).to_csv(OUT / "BUILD_SUMMARY.tsv", sep="\t", index=False)
    retained = set(final.SNP_norm)
    selection_map = preld[["SNP_norm", "position", "min_P_all_exposures", "driving_exposure"]].copy()
    selection_map["retained_panel"] = selection_map["SNP_norm"].isin(retained)
    selection_map.to_csv(OUT / "PRIMARY4_PANEL_SELECTION_MAP.tsv", sep="\t", index=False)
    preld_rank = preld["min_P_all_exposures"].rank(method="first").astype(int)
    tracks = []
    for gene in GENES:
        track = preld[["SNP_norm", "position", f"b_{gene}", f"P_{gene}"]].copy()
        track.columns = ["SNP_norm", "position", "effect_estimate", "p_value"]
        track.insert(0, "exposure", gene)
        track.insert(1, "chromosome", 19)
        track["pre_ld_rank_by_min_p"] = preld_rank.to_numpy()
        track["neg_log10_p"] = -np.log10(track["p_value"].clip(lower=np.finfo(float).tiny))
        track["display_neg_log10_p"] = track["neg_log10_p"].clip(upper=80)
        track["retained_panel"] = track["SNP_norm"].isin(retained)
        tracks.append(track)
    pd.concat(tracks, ignore_index=True).to_csv(OUT / "PRIMARY4_CIS_EQTL_TRACKS.tsv", sep="\t", index=False)
    raw_for_plot = ld.copy()
    np.fill_diagonal(raw_for_plot, 0.0)
    pd.DataFrame([{
        "retained_panel_n": len(final),
        "maximum_retained_pairwise_r2": float(np.max(np.square(raw_for_plot))),
        "maximum_retained_absolute_r": float(np.max(np.abs(raw_for_plot))),
    }]).to_csv(OUT / "PRIMARY4_LD_PLOT_SUMMARY.tsv", sep="\t", index=False)
    if not COMPONENT_SHARING_SOURCE.is_file():
        raise FileNotFoundError(f"Missing frozen component-sharing source table: {COMPONENT_SHARING_SOURCE}")
    pd.read_csv(COMPONENT_SHARING_SOURCE, sep="\t").to_csv(OUT / "OCEL1_COMPONENT_SHARING.tsv", sep="\t", index=False)


if __name__ == "__main__":
    main()
```
