## Supplementary material for "Transcriptomic and Machine-Learning Prioritization of OCEL1 in Low Bone Mineral Density With Multivariable cis-MR": Online Resource 2: Reproducibility Materials: 07_build_rxy_null_pool.py

```
"""Construct full-cis, four-exposure null pools for empirical Rxy estimation."""
from __future__ import annotations

import csv
import gzip
from pathlib import Path

import numpy as np
import pandas as pd
from scipy import sparse


PROJECT = Path(__file__).resolve().parents[2]
CLEAN = Path(__file__).resolve().parents[1]
DATA = PROJECT / "00_DATA_PAPER1" / "mr" / "source"
PANEL = CLEAN / "outputs" / "06_primary4_panel"
OUT = CLEAN / "outputs" / "07_rxy_null_pool"
GENES = ("OCEL1", "NR2F6", "MRPL34", "BABAM1")
COMP = str.maketrans("ACGT", "TGCA")


def flip(value: str) -> str:
    return value.translate(COMP)


def compatible(ref_ea: str, ref_oa: str, ea: str, oa: str) -> float | None:
    if {ea, oa} in ({"A", "T"}, {"C", "G"}):
        return None
    if ea == ref_ea: return 1.0
    if oa == ref_ea: return -1.0
    if flip(ea) == ref_ea: return 1.0
    if flip(oa) == ref_ea: return -1.0
    return None


def strict_compatible(ref_ea: str, ref_oa: str, ea: str, oa: str) -> float | None:
    if {ea, oa} in ({"A", "T"}, {"C", "G"}):
        return None
    if (ea, oa) == (ref_ea, ref_oa): return 1.0
    if (ea, oa) == (ref_oa, ref_ea): return -1.0
    if (flip(ea), flip(oa)) == (ref_ea, ref_oa): return 1.0
    if (flip(ea), flip(oa)) == (ref_oa, ref_ea): return -1.0
    return None


def read_four_gene_cis() -> tuple[dict[str, dict[str, dict[str, object]]], dict[str, int]]:
    source = DATA / "eqtlgen" / "cis_eqtlgen_phase1.tsv.gz"
    values: dict[str, dict[str, dict[str, object]]] = {gene: {} for gene in GENES}
    duplicate_rows: dict[str, int] = {gene: 0 for gene in GENES}
    with gzip.open(source, "rt", encoding="utf-8-sig", newline="") as handle:
        for row in csv.DictReader(handle, delimiter="\t"):
            gene = row.get("GeneSymbol", "").strip().upper()
            if gene not in values:
                continue
            try:
                item = {"EA": row["AssessedAllele"].strip().upper(), "OA": row["OtherAllele"].strip().upper(),
                        "Z": float(row["Zscore"]), "N": float(row["NrSamples"]),
                        "chr": int(float(row["SNPChr"])), "pos": int(float(row["SNPPos"]))}
            except (KeyError, TypeError, ValueError):
                continue
            snp = row["SNP"].strip().upper()
            if snp in values[gene]:
                duplicate_rows[gene] += 1
                continue
            values[gene][snp] = item
    return values, duplicate_rows


def thin(matrix: np.ndarray, r2: float) -> np.ndarray:
    keep: list[int] = []
    threshold = np.sqrt(r2)
    for i in range(matrix.shape[0]):
        if not keep or np.max(np.abs(matrix[i, keep])) < threshold:
            keep.append(i)
    return np.asarray(keep, dtype=int)


def main() -> None:
    OUT.mkdir(parents=True, exist_ok=True)
    rows, duplicate_rows = read_four_gene_cis()
    common = sorted(set.intersection(*(set(rows[g]) for g in GENES)))
    candidates = pd.read_csv(PANEL / "01_exposure_candidate_union.tsv", sep="\t")
    formal = pd.read_csv(PANEL / "PRIMARY_268_PANEL.tsv", sep="\t")
    excluded = set(candidates.loc[candidates.eligible_min_P_lt_2e_5, "SNP_norm"].astype(str).str.upper()) | set(formal.SNP_norm.astype(str).str.upper())
    record_list = []
    orientation_qc = []
    for snp in common:
        ref = rows["OCEL1"][snp]
        z = {"OCEL1": float(ref["Z"])}
        valid = True
        for gene in GENES[1:]:
            item = rows[gene][snp]
            sign = compatible(str(ref["EA"]), str(ref["OA"]), str(item["EA"]), str(item["OA"]))
            strict_sign = strict_compatible(str(ref["EA"]), str(ref["OA"]), str(item["EA"]), str(item["OA"]))
            orientation_qc.append({"gene": gene, "SNP_norm": snp, "allele_sign": sign, "strict_pair_sign": strict_sign,
                                   "same_decision": sign == strict_sign})
            if sign is None:
                valid = False
                break
            z[gene] = sign * float(item["Z"])
        if valid:
            record_list.append({"SNP_norm": snp, "chr": ref["chr"], "position": ref["pos"], "ref_EA": ref["EA"], "ref_OA": ref["OA"],
                                "selected_variant_excluded": snp in excluded, **{f"Z_{g}": z[g] for g in GENES}, **{f"N_{g}": rows[g][snp]["N"] for g in GENES}})
    full = pd.DataFrame(record_list)
    pd.DataFrame({"gene": list(duplicate_rows), "duplicate_gene_snp_rows": list(duplicate_rows.values())}).to_csv(OUT / "RXY_DUPLICATE_GENE_SNP_QC.tsv", sep="\t", index=False)
    orientation = pd.DataFrame(orientation_qc)
    orientation.groupby("gene", dropna=False).agg(n_compared=("SNP_norm", "size"), n_decision_mismatch=("same_decision", lambda x: int((~x).sum()))).reset_index().to_csv(OUT / "RXY_ALLELE_ORIENTATION_QC.tsv", sep="\t", index=False)
    full.to_csv(OUT / "01_common_full_cis_harmonized.tsv.gz", sep="\t", index=False, compression="gzip")
    after_exclusion = full.loc[~full.selected_variant_excluded].copy()
    after_exclusion["max_abs_Z"] = after_exclusion[[f"Z_{g}" for g in GENES]].abs().max(axis=1)
    null = after_exclusion.copy()
    null = null.loc[null.max_abs_Z <= 2].copy()
    index = pd.read_csv(DATA / "ukbb337k" / "chr19_variant_index.tsv", sep="\t", dtype=str)
    index["SNP_norm"] = index.rsid_norm.str.upper()
    index_raw_n = len(index)
    index_duplicate_row_n = int(index.duplicated("SNP_norm", keep="first").sum())
    index_duplicate_snp_n = int(index.loc[index.duplicated("SNP_norm", keep=False), "SNP_norm"].nunique())
    index = index.drop_duplicates("SNP_norm", keep="first").copy()
    pd.DataFrame([{
        "ukbb_index_raw_rows": index_raw_n,
        "ukbb_index_unique_SNP_norm": len(index),
        "ukbb_index_duplicate_rows_dropped": index_duplicate_row_n,
        "ukbb_index_duplicate_SNP_norm": index_duplicate_snp_n,
        "deduplication_rule": "drop_duplicates_SNP_norm_keep_first_matches_stage06",
    }]).to_csv(OUT / "UKBB_INDEX_DUPLICATE_AUDIT.tsv", sep="\t", index=False)
    null = null.merge(index[["SNP_norm", "ukbb_index"]], on="SNP_norm", how="inner")
    null["ukbb_index"] = null.ukbb_index.astype(int)
    null.to_csv(OUT / "02_allnull_ld_mapped.tsv.gz", sep="\t", index=False, compression="gzip")
    raw = sparse.load_npz(DATA / "ukbb337k" / "chr19_16000001_19000001.npz").tocsr()
    ld = (raw + raw.T).tocsr(); ld.setdiag(1.0)
    ids = null.ukbb_index.to_numpy()
    sub = ld[ids, :][:, ids].toarray(); sub = (sub + sub.T) / 2; np.fill_diagonal(sub, 1.0)
    outputs = []
    for r2, label in ((0.10, "RXY_R2_010"), (0.01, "RXY_R2_001")):
        kept = null.iloc[thin(sub, r2)].copy(); kept["r2_threshold"] = r2
        kept.to_csv(OUT / f"03_{label}_null_pool.tsv.gz", sep="\t", index=False, compression="gzip")
        outputs.append({"specification": label, "n_null": len(kept), "r2_threshold": r2})
    pd.DataFrame(outputs + [{"specification": "RXY_ALLNULL", "n_null": len(null), "r2_threshold": np.nan}]).to_csv(OUT / "RXY_NULL_POOL_SUMMARY.tsv", sep="\t", index=False)
    pd.DataFrame([{"four_exposure_common_harmonized_n": len(full), "after_formal_variant_exclusion_n": len(after_exclusion), "max_abs_z_le_2_n": int((after_exclusion.max_abs_Z <= 2).sum()), "ukbb_mapped_n": len(null), "r2_010_n": int(outputs[0]["n_null"]), "r2_001_n": int(outputs[1]["n_null"])}]).to_csv(OUT / "RXY_FLOW_SUMMARY.tsv", sep="\t", index=False)


if __name__ == "__main__":
    main()
```
