## Supplementary material for "Transcriptomic and Machine-Learning Prioritization of OCEL1 in Low Bone Mineral Density With Multivariable cis-MR": Online Resource 2: Reproducibility Materials: 08_estimate_empirical_rxy.R

```
source(file.path(dirname(normalizePath(sub('^--file=', '', grep('^--file=', commandArgs(FALSE), value = TRUE)[1]), winslash = '/', mustWork = FALSE)), '00_paths.R'))
if (!requireNamespace('MRBEE', quietly = TRUE)) stop('MRBEE must be installed for empirical exposure-error correlation estimation.')
suppressPackageStartupMessages(library(MRBEE))
genes <- c('OCEL1', 'NR2F6', 'MRPL34', 'BABAM1')
in_dir <- file.path(OUTPUT_ROOT, '07_rxy_null_pool')
out_dir <- file.path(OUTPUT_ROOT, '08_empirical_rxy'); dir.create(out_dir, recursive = TRUE, showWarnings = FALSE)
specs <- c(RXY_ALLNULL='02_allnull_ld_mapped.tsv.gz', RXY_R2_010='03_RXY_R2_010_null_pool.tsv.gz', RXY_R2_001='03_RXY_R2_001_null_pool.tsv.gz')
out <- lapply(names(specs), function(label) {
  dat <- read.delim(gzfile(file.path(in_dir, specs[[label]])), check.names = FALSE)
  z <- as.matrix(dat[, paste0('Z_', genes)])
  set.seed(20260910)
  rx <- MRBEE::errorCov(z, Zscore.cutoff = 2, subsampling.ratio = .1, subsampling.time = 1000)
  rownames(rx) <- colnames(rx) <- genes
  ev <- eigen(rx, symmetric = TRUE, only.values = TRUE)$values
  data.frame(specification=label, pair=as.vector(outer(genes, genes, paste, sep='--')), rho=as.vector(rx), n_null=nrow(dat), min_eigenvalue=min(ev), condition_number=max(ev)/min(ev))
})
identity <- data.frame(specification='RXY_IDENTITY', pair=as.vector(outer(genes, genes, paste, sep='--')), rho=as.vector(diag(4)), n_null=NA_integer_, min_eigenvalue=1, condition_number=1)
write.table(do.call(rbind, c(list(identity), out)), file.path(out_dir, 'ALL_RXY_MATRICES.tsv'), sep='\t', quote=FALSE, row.names=FALSE)
primary <- out[[which(names(specs) == 'RXY_R2_010')]]
rx <- matrix(primary$rho, nrow=4, ncol=4, dimnames=list(genes, genes))
rxy5 <- diag(5); dimnames(rxy5) <- list(c(genes, 'heel_eBMD'), c(genes, 'heel_eBMD')); rxy5[genes, genes] <- rx
write.table(rxy5, file.path(out_dir, 'PRIMARY_RXY_5X5.tsv'), sep='\t', quote=FALSE, col.names=NA)
writeLines(capture.output(sessionInfo()), file.path(out_dir, 'sessionInfo.txt'))
```
