## Supplementary material for "Transcriptomic and Machine-Learning Prioritization of OCEL1 in Low Bone Mineral Density With Multivariable cis-MR": Online Resource 2: Reproducibility Materials: 09_run_primary4_cismrbeex.R

```
source(file.path(dirname(normalizePath(sub('^--file=', '', grep('^--file=', commandArgs(FALSE), value = TRUE)[1]), winslash = '/', mustWork = FALSE)), '00_paths.R'))
mrbeex_rlib <- Sys.getenv('PAPER1_MRBEEX_RLIB', unset = '')
if (nzchar(mrbeex_rlib)) .libPaths(c(mrbeex_rlib, .libPaths()))
if (!requireNamespace('MRBEEX', quietly = TRUE) || !requireNamespace('susieR', quietly = TRUE)) stop('MRBEEX and susieR must be installed before running the four-exposure MR stage.')
suppressPackageStartupMessages({library(MRBEEX); library(susieR)})
if (!exists('susie_ss', envir = asNamespace('susieR'), inherits = FALSE)) {
  # MRBEEX 0.1.0 names the sufficient-statistic SuSiE entry point used before
  # susieR renamed it to susie_suff_stat. This adapter changes no model inputs.
  compat_env <- new.env(parent = asNamespace('MRBEEX'))
  compat_env$susie_ss <- susieR::susie_suff_stat
  ipod_susie <- get('Cis_MRBEE_IPOD_SuSiE', envir = asNamespace('MRBEEX'))
  environment(ipod_susie) <- compat_env
  compat_env$Cis_MRBEE_IPOD_SuSiE <- ipod_susie
  environment(CisMRBEEX) <- compat_env
}
genes <- c('OCEL1', 'NR2F6', 'MRPL34', 'BABAM1')
panel_dir <- file.path(OUTPUT_ROOT, '06_primary4_panel')
panel <- read.delim(file.path(panel_dir, 'PRIMARY_268_PANEL.tsv'), check.names = FALSE)
ld <- as.matrix(read.delim(file.path(panel_dir, 'PRIMARY_268_RAW_LD.tsv'), row.names = 1, check.names = FALSE))
panel <- panel[complete.cases(panel[, c(paste0('b_', genes), paste0('se_', genes), 'b_heel_std', 'se_heel_std')]), ]
ld <- ld[panel$SNP_norm, panel$SNP_norm, drop = FALSE]
rx <- read.delim(file.path(OUTPUT_ROOT, '08_empirical_rxy', 'ALL_RXY_MATRICES.tsv'), check.names = FALSE)
rx <- rx[rx$specification == 'RXY_R2_010', ]
rxy <- diag(5); dimnames(rxy) <- list(c(genes, 'heel_eBMD'), c(genes, 'heel_eBMD'))
for (i in seq_len(nrow(rx))) { pair <- strsplit(rx$pair[i], '--', fixed = TRUE)[[1]]; if (all(pair %in% genes)) rxy[pair[1], pair[2]] <- as.numeric(rx$rho[i]) }
rxy[genes, genes] <- (rxy[genes, genes] + t(rxy[genes, genes])) / 2; diag(rxy) <- 1
poet <- function(R) {
  R[is.na(R)] <- 0; diag(R) <- 1; R[abs(R) < 1e-4] <- 0
  ee <- eigen(R, symmetric = TRUE); d <- ee$values; U <- ee$vectors; km <- min(15L, round(nrow(R) / 2))
  ratios <- vapply(2:km, function(j) (d[j - 1] - d[j]) / (d[j] - d[j + 1]), numeric(1)); k <- which.max(ratios) + 1L
  P <- U[, seq_len(k), drop = FALSE] %*% diag(d[seq_len(k)]) %*% t(U[, seq_len(k), drop = FALSE]); h <- (sum(diag(R)) - sum(d[seq_len(k)])) / (ncol(R) - k)
  E <- R - P; v <- diag(E); v[v < 0] <- max(h, 0.01); diag(E) <- v
  selected <- FALSE
  for (q in seq(0.025, 0.25, 0.025)) {
    regularized <- P + E * (1 - q) + diag(diag(E)) * q
    if (min(eigen(regularized, symmetric = TRUE, only.values = TRUE)$values) > 0.001) {
      selected <- TRUE
      break
    }
  }
  if (!selected) stop('POET regularization did not achieve the required positive-definiteness threshold.')
  list(LD = cov2cor(regularized), K = k, lambda = q)
}
q <- poet(ld); n <- vapply(genes, function(g) mean(panel[[paste0('N_', g)]]), numeric(1)); z <- as.matrix(panel[, paste0('b_', genes)]) / as.matrix(panel[, paste0('se_', genes)])
prefit <- lapply(seq_len(ncol(z)), function(i) {
  first <- susie_rss(z = z[, i], R = q$LD, n = n[i], L = 15, max_iter = 2000, coverage = 0.95)
  if (!isTRUE(first$converged)) stop('Initial SuSiE-RSS prefit did not converge for ', genes[i])
  second <- susie_rss(z = z[, i], R = q$LD, n = n[i], L = length(first$sets$cs) + 1L, max_iter = 2000, coverage = 0.95)
  if (!isTRUE(second$converged)) stop('Final SuSiE-RSS prefit did not converge for ', genes[i])
  second
})
set.seed(20260829)
fit <- CisMRBEEX(causal.pip.thres = 0.2, by = panel$b_heel_std, bX = as.matrix(panel[, paste0('b_', genes)]), byse = panel$se_heel_std, bXse = as.matrix(panel[, paste0('se_', genes)]), LD = q$LD, Rxy = rxy, reliability.thres = 0.75, xQTL.max.L = 15, xQTL.pip.thres = 0.3, xQTL.Nvec = n, tauvec = seq(4.5, 30, 1.5), susie.iter = 500, ridge.diff = 100, ebic.gamma = 0, xQTLfitList = prefit)
if (!('susie.theta' %in% names(fit)) || !isTRUE(fit$susie.theta$converged)) stop('CisMRBEEX causal fit did not converge.')
theta <- as.numeric(fit$theta); se <- as.numeric(fit$theta.se); ztheta <- theta / se
out <- data.frame(exposure = genes, beta = theta, se = se, ci_lower = theta - qnorm(.975) * se, ci_upper = theta + qnorm(.975) * se, p_value = pchisq(ztheta^2, 1, lower.tail = FALSE), theta_pip = as.numeric(fit$theta.pip), theta_pratt = as.numeric(fit$theta.pratt), n_snp = nrow(panel), poet_K = q$K, poet_lambda = q$lambda)
out_dir <- file.path(OUTPUT_ROOT, '09_primary4_mr'); dir.create(out_dir, recursive = TRUE, showWarnings = FALSE)
raw_ev <- eigen(ld, symmetric=TRUE, only.values=TRUE)$values; poet_ev <- eigen(q$LD, symmetric=TRUE, only.values=TRUE)$values
write.table(out, file.path(out_dir, 'PRIMARY4_CISMRBEEX_RESULT.tsv'), sep = '\t', quote = FALSE, row.names = FALSE); write.table(q$LD, file.path(out_dir, 'PRIMARY4_POET_LD.tsv'), sep = '\t', quote = FALSE, col.names = NA); saveRDS(fit, file.path(out_dir, 'PRIMARY4_CISMRBEEX_FIT.rds'))
write.table(data.frame(raw_ld_min_eigen=min(raw_ev), raw_ld_condition_number=kappa(ld, exact=TRUE), poet_ld_min_eigen=min(poet_ev), poet_ld_condition_number=kappa(q$LD, exact=TRUE), poet_K=q$K, poet_lambda=q$lambda), file.path(out_dir, 'PRIMARY4_POET_DIAGNOSTICS.tsv'), sep='\t', quote=FALSE, row.names=FALSE)
write.table(data.frame(exposure=genes, mean_xQTL_N=n, susie_converged=vapply(prefit, function(x) isTRUE(x$converged), logical(1))), file.path(out_dir, 'PRIMARY4_XQTL_N.tsv'), sep='\t', quote=FALSE, row.names=FALSE)
status <- ifelse(is.finite(out$theta_pip) & out$theta_pip >= .2, 'RETAINED_PIP_GE_0.2', 'NOT_RETAINED_PIP_LT_0.2')
write.table(data.frame(exposure=genes, theta_pip=out$theta_pip, beta=out$beta, se=out$se, model_status=status), file.path(out_dir, 'PRIMARY4_MODEL_STATUS.tsv'), sep='\t', quote=FALSE, row.names=FALSE)
write.table(data.frame(causal_susie_converged=if ('susie.theta' %in% names(fit)) as.character(isTRUE(fit$susie.theta$converged)) else 'NOT_REPORTED_BY_CISMRBEEX_OBJECT', fit_object_fields=paste(names(fit), collapse=';')), file.path(out_dir, 'PRIMARY4_CONVERGENCE.tsv'), sep='\t', quote=FALSE, row.names=FALSE)
writeLines(capture.output(sessionInfo()), file.path(out_dir, 'sessionInfo.txt') )
message('PASS: primary four-exposure CisMRBEEX analysis completed.')
```
