## Supplementary material for "Transcriptomic and Machine-Learning Prioritization of OCEL1 in Low Bone Mineral Density With Multivariable cis-MR": Online Resource 2: Reproducibility Materials: 10_run_primary4_mr_sensitivities.R

```
source(file.path(dirname(normalizePath(sub('^--file=', '', grep('^--file=', commandArgs(FALSE), value = TRUE)[1]), winslash = '/', mustWork = FALSE)), '00_paths.R'))
mrbeex_rlib <- Sys.getenv('PAPER1_MRBEEX_RLIB', unset = '')
if (nzchar(mrbeex_rlib)) .libPaths(c(mrbeex_rlib, .libPaths()))
if (!requireNamespace('MRBEEX', quietly = TRUE) || !requireNamespace('susieR', quietly = TRUE)) stop('MRBEEX and susieR must be installed.')
suppressPackageStartupMessages({library(MRBEEX); library(susieR)})
genes <- c('OCEL1', 'NR2F6', 'MRPL34', 'BABAM1'); panel_dir <- file.path(OUTPUT_ROOT, '06_primary4_panel'); out_dir <- file.path(OUTPUT_ROOT, '10_mr_sensitivities'); dir.create(out_dir, recursive = TRUE, showWarnings = FALSE)
rx_table <- read.delim(file.path(OUTPUT_ROOT, '08_empirical_rxy', 'ALL_RXY_MATRICES.tsv'), check.names = FALSE)
make_rxy <- function(spec) { x <- rx_table[rx_table$specification == spec, ]; m <- diag(5); dimnames(m) <- list(c(genes, 'heel_eBMD'), c(genes, 'heel_eBMD')); for (i in seq_len(nrow(x))) { p <- strsplit(x$pair[i], '--', fixed = TRUE)[[1]]; if (all(p %in% genes)) m[p[1], p[2]] <- as.numeric(x$rho[i]) }; m[genes, genes] <- (m[genes, genes] + t(m[genes, genes])) / 2; diag(m) <- 1; m }
poet <- function(R, K = NA_integer_) {
  R[is.na(R)] <- 0
  diag(R) <- 1
  R[abs(R) < 1e-4] <- 0
  decomposition <- eigen(R, symmetric = TRUE)
  values <- decomposition$values
  vectors <- decomposition$vectors
  max_k <- min(15L, round(nrow(R) / 2))
  ratios <- vapply(2:max_k, function(j) (values[j - 1] - values[j]) / (values[j] - values[j + 1]), numeric(1))
  k <- if (is.na(K)) which.max(ratios) + 1L else K
  low_rank <- vectors[, seq_len(k), drop = FALSE] %*% diag(values[seq_len(k)]) %*% t(vectors[, seq_len(k), drop = FALSE])
  residual <- R - low_rank
  residual_variance <- diag(residual)
  residual_variance[residual_variance < 0] <- max((sum(diag(R)) - sum(values[seq_len(k)])) / (ncol(R) - k), 0.01)
  diag(residual) <- residual_variance
  selected <- FALSE
  for (lambda in seq(0.025, 0.25, 0.025)) {
    regularized <- low_rank + residual * (1 - lambda) + diag(diag(residual)) * lambda
    if (min(eigen(regularized, symmetric = TRUE, only.values = TRUE)$values) > .001) {
      selected <- TRUE
      break
    }
  }
  if (!selected) stop('POET regularization did not yield a positive-definite LD matrix.')
  list(LD = cov2cor(regularized), K = k, lambda = lambda)
}

read_panel <- function(panel_path, ld_path) {
  data <- read.delim(panel_path, check.names = FALSE)
  required <- c(paste0('b_', genes), paste0('se_', genes), 'b_heel_std', 'se_heel_std')
  data <- data[complete.cases(data[, required]), ]
  raw_ld <- as.matrix(read.delim(ld_path, row.names = 1, check.names = FALSE))
  list(data = data, LD = raw_ld[data$SNP_norm, data$SNP_norm, drop = FALSE])
}

fit_exposure_susie <- function(beta, se, n, ld) {
  first <- susie_rss(z = beta / se, R = ld, n = n, L = 15, max_iter = 2000, coverage = .95)
  if (!isTRUE(first$converged)) stop('Initial SuSiE fit failed to converge.')
  final <- susie_rss(z = beta / se, R = ld, n = n, L = length(first$sets$cs) + 1L, max_iter = 2000, coverage = .95)
  if (!isTRUE(final$converged)) stop('Final SuSiE fit failed to converge.')
  final
}

fit_one <- function(label, input, selected_genes, rxy, K = NA_integer_) {
  poet_fit <- poet(input$LD, K)
  data <- input$data
  beta_x <- as.matrix(data[, paste0('b_', selected_genes), drop = FALSE])
  se_x <- as.matrix(data[, paste0('se_', selected_genes), drop = FALSE])
  sample_size <- vapply(selected_genes, function(gene) mean(data[[paste0('N_', gene)]]), numeric(1))
  xqtl_fits <- lapply(seq_len(ncol(beta_x)), function(index) fit_exposure_susie(beta_x[, index], se_x[, index], sample_size[index], poet_fit$LD))
  set.seed(20260829)
  fit <- CisMRBEEX(causal.pip.thres = .2, by = data$b_heel_std, bX = beta_x, byse = data$se_heel_std, bXse = se_x, LD = poet_fit$LD,
                   Rxy = rxy[c(selected_genes, 'heel_eBMD'), c(selected_genes, 'heel_eBMD'), drop = FALSE], reliability.thres = .75,
                   xQTL.max.L = 15, xQTL.pip.thres = .3, xQTL.Nvec = sample_size, tauvec = seq(4.5, 30, 1.5), susie.iter = 500,
                   ridge.diff = 100, ebic.gamma = 0, xQTLfitList = xqtl_fits)
  if (is.null(fit$susie.theta) || !isTRUE(fit$susie.theta$converged)) stop('CisMRBEEX causal fit failed to converge.')
  theta <- as.numeric(fit$theta)
  se <- as.numeric(fit$theta.se)
  data.frame(run = label, exposure = selected_genes, beta = theta, se = se, ci_lower = theta - qnorm(.975) * se,
             ci_upper = theta + qnorm(.975) * se, p_value = pchisq((theta / se)^2, 1, lower.tail = FALSE),
             theta_pip = as.numeric(fit$theta.pip), theta_pratt = as.numeric(fit$theta.pratt), n_snp = nrow(data),
             poet_K = poet_fit$K, poet_lambda = poet_fit$lambda)
}

primary <- read_panel(file.path(panel_dir, 'PRIMARY_268_PANEL.tsv'), file.path(panel_dir, 'PRIMARY_268_RAW_LD.tsv'))
specs <- c('RXY_IDENTITY', 'RXY_ALLNULL', 'RXY_R2_010', 'RXY_R2_001')
rxy_out <- do.call(rbind, lapply(specs, function(s) fit_one(s, primary, genes, make_rxy(s))))
write.table(rxy_out, file.path(out_dir, 'PRIMARY4_RXY_SPECIFICATION_SENSITIVITY.tsv'), sep = '\t', quote = FALSE, row.names = FALSE)
models <- list(O = 'OCEL1', O_N = c('OCEL1', 'NR2F6'), O_M = c('OCEL1', 'MRPL34'), O_B = c('OCEL1', 'BABAM1'), O_N_M = c('OCEL1', 'NR2F6', 'MRPL34'), O_N_B = c('OCEL1', 'NR2F6', 'BABAM1'), O_M_B = c('OCEL1', 'MRPL34', 'BABAM1'), O_N_M_B = genes)
lattice <- do.call(rbind, Map(function(nm, g) fit_one(nm, primary, g, make_rxy('RXY_R2_010')), names(models), models))
write.table(lattice, file.path(out_dir, 'PRIMARY4_MODEL_LATTICE.tsv'), sep = '\t', quote = FALSE, row.names = FALSE)
k12 <- fit_one('RXY_R2_010_K12', primary, genes, make_rxy('RXY_R2_010'), 12L)
write.table(k12, file.path(out_dir, 'PRIMARY4_K12_DIAGNOSTIC.tsv'), sep = '\t', quote = FALSE, row.names = FALSE)
nopal <- read_panel(file.path(panel_dir, 'PRIMARY_263_NO_PAL_PANEL.tsv'), file.path(panel_dir, 'PRIMARY_263_NO_PAL_RAW_LD.tsv'))
write.table(fit_one('RXY_R2_010_NO_PAL', nopal, genes, make_rxy('RXY_R2_010')), file.path(out_dir, 'PRIMARY4_NO_PAL_SENSITIVITY.tsv'), sep = '\t', quote = FALSE, row.names = FALSE)
message('PASS: MR specification, lattice, K12, and no-palindromic sensitivity tables were generated.')
```
