## Supplementary material for "Transcriptomic and Machine-Learning Prioritization of OCEL1 in Low Bone Mineral Density With Multivariable cis-MR": Online Resource 2: Reproducibility Materials: build_publication_figures.py

```
"""Create the Paper 1 publication figure library from final statistical layer tables."""
from __future__ import annotations

import os
from pathlib import Path

import matplotlib.patches as patches
import matplotlib.pyplot as plt
import numpy as np
import pandas as pd
from scipy.cluster.hierarchy import leaves_list, linkage


ROOT = Path(__file__).resolve().parents[1]
STATISTICAL_LAYER = Path(os.environ.get(
    "STATISTICAL_LAYER_OUTPUTS",
    ROOT.parent / "06_STATISTICAL_LAYER_KEY_RESULTS",
))
PANELS = ROOT / "03_PANEL_OUTPUTS"
FIGURES = ROOT / "05_COMPOSITE_FIGURES"

COLORS = {
    "ink": "#28343b", "muted": "#9caeb7", "light": "#e9eff2",
    "red": "#b34848", "blue": "#347ba8", "green": "#6f922d",
    "purple": "#8169ae", "orange": "#c98255", "darkred": "#8e2929",
}

plt.rcParams.update({
    "font.family": ["Arial", "Helvetica", "Liberation Sans", "DejaVu Sans"], "font.size": 9, "axes.labelsize": 9,
    "axes.titlesize": 10, "axes.titleweight": "bold", "axes.linewidth": .8,
    "xtick.labelsize": 8, "ytick.labelsize": 8,
})


def read(relative: str) -> pd.DataFrame:
    path = STATISTICAL_LAYER / relative
    return pd.read_csv(path, sep="\t" if path.suffix == ".tsv" else ",")


def label(ax: plt.Axes, value: str) -> None:
    ax.text(-.12, 1.08, value, transform=ax.transAxes, fontsize=14,
            fontweight="bold", va="top", ha="left")


def clean(ax: plt.Axes) -> None:
    ax.spines[["top", "right"]].set_visible(False)


def save(fig: plt.Figure, folder: Path, stem: str) -> None:
    folder.mkdir(parents=True, exist_ok=True)
    fig.savefig(folder / f"{stem}.png", dpi=400, bbox_inches="tight", facecolor="white")
    fig.savefig(folder / f"{stem}.pdf", bbox_inches="tight", facecolor="white")
    plt.close(fig)


def standalone(stem: str, draw, figsize=(7.2, 5.5)) -> None:
    fig, ax = plt.subplots(figsize=figsize)
    draw(ax)
    save(fig, PANELS, stem)


def draw_f2a(ax: plt.Axes) -> None:
    path = read("03_ml_fit/LASSO_COEFFICIENT_PATH.tsv")
    meta = read("03_ml_fit/LASSO_SELECTION_METADATA.tsv").iloc[0]
    selected = list(read("03_ml_fit/lasso_lambda_1se_nonzero_genes.csv")["gene"])
    palette = plt.get_cmap("tab20")(np.linspace(0, 1, max(1, len(selected))))
    selected_colors = dict(zip(selected, palette))
    for gene, block in path.groupby("gene", sort=False):
        if gene in selected_colors:
            ax.plot(np.log(block["lambda"]), block["coefficient"], color=selected_colors[gene], lw=1.0, alpha=.9)
        else:
            ax.plot(np.log(block["lambda"]), block["coefficient"], color="#c8d2d6", lw=.4, alpha=.28)
    ax.axvline(np.log(meta["selected_lambda"]), color=COLORS["green"], ls="--", lw=1)
    ax.set(xlabel="log(lambda)", ylabel="Coefficient")
    clean(ax)


def draw_f2b(ax: plt.Axes) -> None:
    d = read("04_ml_selection/xgb_frequency_with_rule.csv").head(16).iloc[::-1].copy()
    selected = d["selected_by_rule"].astype(str).str.upper().eq("TRUE") | d["selected_by_rule"].eq(True)
    colors = np.where(selected, "#4c94be", "#bedceb")
    ax.barh(d["Feature"], d["frequency_share"], color=colors, edgecolor="none")
    ax.set(xlabel="Split frequency")
    clean(ax)


def draw_f2c(ax: plt.Axes) -> None:
    d = read("04_ml_selection/svm_profile_with_rule.csv")
    n = int(read("04_ml_selection/ml_selection_counts.csv").query("method == 'Linear_SVM_weight_ranking_max_accuracy_tie'").iloc[0]["n"])
    chosen = d[d["Variables"] == n].iloc[0]
    ax.plot(d["Variables"], d["Accuracy"], "o-", color=COLORS["blue"], ms=3, lw=1.2)
    ax.scatter(chosen["Variables"], chosen["Accuracy"], marker="D", s=36, color=COLORS["red"], zorder=3)
    ax.axvline(n, color=COLORS["red"], ls="--", lw=.9)
    ax.axhline(chosen["Accuracy"], color=COLORS["red"], ls="--", lw=.9)
    ax.set(xlabel="Number of selected features", ylabel="Repeated-CV accuracy")
    # Keep the selected 24-feature solution visible without the crowded 24/25
    # pair introduced by Matplotlib's automatic integer tick locator.
    ax.set_xticks(sorted({2, 5, 10, 15, 20, n, 30, 35, 40}))
    ax.set_yticks(sorted(set(ax.get_yticks()).union({float(chosen['Accuracy'])})))
    for tick, value in zip(ax.get_xticklabels(), ax.get_xticks()):
        if np.isclose(value, n): tick.set_color(COLORS["red"]); tick.set_fontweight("bold")
    for tick, value in zip(ax.get_yticklabels(), ax.get_yticks()):
        if np.isclose(value, chosen["Accuracy"]): tick.set_color(COLORS["red"]); tick.set_fontweight("bold")
    ax.text(.97, .05, f"Selected: {n} features; accuracy = {chosen['Accuracy']:.3f}", transform=ax.transAxes, ha="right", color=COLORS["red"], fontweight="bold", fontsize=7.5)
    clean(ax)


def draw_f2d(ax: plt.Axes) -> None:
    cv = read("03_ml_fit/lasso_lambda_table.csv").sort_values("lambda", ascending=False)
    meta = read("03_ml_fit/LASSO_SELECTION_METADATA.tsv").iloc[0]
    x = np.log(cv["lambda"])
    ax.errorbar(x, cv["cvm"], yerr=cv["cvsd"], fmt="o-", color=COLORS["blue"], ms=2.5, lw=.8)
    ax.axvline(np.log(meta["lambda_min"]), color=COLORS["muted"], ls="--", lw=.9, label="lambda.min")
    ax.axvline(np.log(meta["lambda_1se"]), color=COLORS["green"], ls="--", lw=1.1, label="lambda.1se")
    ax.set(xlabel="log(lambda)", ylabel="Cross-validated error")
    ax.legend(frameon=False, fontsize=7, loc="upper left"); ax.text(.97, .05, f"Selected: {int(meta['selected_nonzero_n'])} genes", transform=ax.transAxes, ha="right", color=COLORS["green"], fontweight="bold")
    clean(ax)


def draw_f3a(ax: plt.Axes) -> None:
    d = read("04_ml_selection/ml_selection_after_rules.csv")
    groups = {method: set(block["gene"].str.upper()) for method, block in d.groupby("method")}
    svm = next(value for key, value in groups.items() if key.startswith("Linear_SVM"))
    lasso = next(value for key, value in groups.items() if key.startswith("LASSO"))
    xgb = next(value for key, value in groups.items() if key.startswith("XGBoost"))
    only = lambda a, b, c: len(a - b - c)
    pair = lambda a, b, c: len((a & b) - c)
    ax.axis("off")
    ax.set(xlim=(0.06, 0.94), ylim=(0.05, 0.94), aspect="equal")

    # Fixed equal-radius circles make the seven disjoint regions readable.
    # Set labels live outside the geometry so they cannot collide with counts.
    circles = [
        (0.38, 0.57, "#4C78A8"),
        (0.62, 0.57, "#F58518"),
        (0.50, 0.39, "#54A24B"),
    ]
    for x, y, color in circles:
        ax.add_patch(plt.Circle((x, y), 0.255, facecolor=color, edgecolor=color, alpha=0.24, linewidth=1.35))

    label_style = dict(ha="center", va="center", fontsize=9, fontweight="bold", color="#263238")
    ax.text(0.30, 0.885, "Linear SVM\n(n = 24)", **label_style)
    ax.text(0.70, 0.885, "LASSO\n(n = 18)", **label_style)
    ax.text(0.50, 0.095, "XGBoost\n(n = 9)", **label_style)

    counts = [
        (0.275, 0.595, only(svm, lasso, xgb)),
        (0.725, 0.595, only(lasso, svm, xgb)),
        (0.500, 0.270, only(xgb, svm, lasso)),
        (0.500, 0.695, pair(svm, lasso, xgb)),
        (0.390, 0.425, pair(svm, xgb, lasso)),
        (0.610, 0.425, pair(lasso, xgb, svm)),
        (0.500, 0.515, len(svm & lasso & xgb)),
    ]
    for x, y, value in counts:
        ax.text(x, y, str(value), ha="center", va="center", fontsize=12, fontweight="bold", color="#111111")


def draw_f3b(ax: plt.Axes) -> None:
    d = read("01_bulk/GSE56815_limma_all_genes.csv").copy()
    d["symbol"] = d["symbol"].str.upper()
    five = set(read("04_ml_selection/strict_intersection.csv")["gene"].str.upper())
    y = -np.log10(d["P.Value"].clip(lower=np.finfo(float).tiny))
    nominal = d["P.Value"].lt(.05)
    up = nominal & d["logFC"].gt(0)
    down = nominal & d["logFC"].lt(0)
    other = ~(up | down)
    ax.scatter(d.loc[other, "logFC"], y[other], s=5, color="#c8d2d6", alpha=.55, linewidths=0, label="Other genes")
    ax.scatter(d.loc[up, "logFC"], y[up], s=6, color=COLORS["red"], alpha=.72, linewidths=0, label="Nominal upregulated")
    ax.scatter(d.loc[down, "logFC"], y[down], s=6, color=COLORS["blue"], alpha=.72, linewidths=0, label="Nominal downregulated")
    focal = d[d["symbol"].isin(five)].copy()
    focal_colors = np.where(focal["logFC"].gt(0), COLORS["darkred"], "#1b5f8a")
    ax.scatter(focal["logFC"], -np.log10(focal["P.Value"]), s=30, color=focal_colors, edgecolor="#1d2930", linewidth=.35, zorder=3)
    offsets = {"CPNE1":(-28,18), "EZR":(12,20), "FAM216A":(12,-18), "MAP3K3":(-35,-18), "OCEL1":(12,8)}
    for _, row in focal.iterrows():
        dx, dy = offsets.get(row["symbol"], (8, 8))
        label_color = COLORS["darkred"] if row["logFC"] > 0 else "#1b5f8a"
        ax.annotate(row["symbol"], xy=(row["logFC"], -np.log10(row["P.Value"])), xytext=(dx, dy), textcoords="offset points", fontsize=7.5, color=label_color, fontweight="bold", arrowprops={"arrowstyle":"-", "lw":.55, "color":"#73828a"})
    ax.axvline(0, color="#64747d", lw=.8)
    ax.axhline(-np.log10(.05), color="#64747d", lw=.8, ls="--")
    ax.set(xlabel="log2 fold change", ylabel="-log10(P)")
    ax.legend(frameon=False, fontsize=6.8, loc="upper right", handletextpad=.3, borderpad=.1)
    clean(ax)


def draw_f3c(ax: plt.Axes) -> None:
    expr = read("01_bulk/GSE56815_expression_plotting_source.tsv").set_index("symbol")
    limma = read("01_bulk/GSE56815_limma_all_genes.csv").copy()
    limma["symbol"] = limma["symbol"].str.upper()
    expr.index = expr.index.astype(str).str.upper()
    five = list(read("04_ml_selection/strict_intersection.csv")["gene"].str.upper())
    ordered = limma.sort_values(["P.Value", "adj.P.Val", "symbol"], kind="stable")["symbol"].tolist()
    # This panel is strictly the top 3,000 genes ranked by nominal P value.
    # Strict-intersection genes are annotated only when they naturally fall in that set.
    genes = ordered[:3000]
    genes = [gene for gene in genes if gene in expr.index]
    samples = read("01_bulk/GSE56815_heatmap_sample_metadata.tsv").sort_values("display_order")
    heat = expr.loc[genes, samples["sample"]].astype(float)
    heat = heat.sub(heat.mean(axis=1), axis=0).div(heat.std(axis=1).replace(0, np.nan), axis=0).fillna(0.0)
    row_order = leaves_list(linkage(heat.to_numpy(), method="average", metric="euclidean"))
    heat = heat.iloc[row_order]
    heat_xlim_right = heat.shape[1] + 2.0
    heat_panel_frac = heat.shape[1] / (heat_xlim_right + .5)
    group_labels = np.where(samples["group"].eq("high_bmd"), "High BMD women", "Low BMD women")
    group_colors = np.where(samples["group"].eq("high_bmd"), "#DCE9F5", "#E8CFCF")
    strip = ax.inset_axes([0.00, .94, heat_panel_frac, .06])
    strip.imshow(np.arange(len(group_colors))[None, :], cmap=plt.matplotlib.colors.ListedColormap(group_colors.tolist()), aspect="auto")
    strip.set_xticks([]); strip.set_yticks([])
    for spine in strip.spines.values(): spine.set_visible(False)
    label_ax = ax.inset_axes([0.00, 1.01, heat_panel_frac, .08])
    label_ax.set_axis_off()
    for group in pd.unique(group_labels):
        idx = np.flatnonzero(group_labels == group)
        xpos = (idx.min() + idx.max()) / 2.0 / max(1, len(group_labels) - 1)
        label_ax.text(xpos, .58, f"{group} (n={len(idx)})", transform=label_ax.transAxes, ha="center", va="center", fontsize=7.8, color="#223039")
    ax.imshow(heat.to_numpy(), cmap="RdBu_r", aspect="auto", vmin=-2.2, vmax=2.2)
    ax.set(xticks=[], yticks=[])
    positions = {gene: int(np.flatnonzero(heat.index.to_numpy() == gene)[0]) for gene in five if gene in heat.index}
    ordered_labels = sorted(positions.items(), key=lambda item: item[1])
    min_sep = 210.0 if heat.shape[0] >= 2500 else 38.0
    label_positions = {}
    placed_y = []
    for gene, y in ordered_labels:
        y_text = float(y)
        if placed_y and y_text - placed_y[-1] < min_sep:
            y_text = placed_y[-1] + min_sep
        label_positions[gene] = y_text
        placed_y.append(y_text)
    if placed_y:
        overflow = placed_y[-1] - (heat.shape[0] - 8)
        if overflow > 0:
            label_positions = {gene: value - overflow for gene, value in label_positions.items()}
    x_mark0, x_mark1, x_label = heat.shape[1] - .5, heat.shape[1] + .18, heat.shape[1] + .92
    for gene, y in ordered_labels:
        color = COLORS["darkred"] if gene == "OCEL1" else "#344b5f"
        y_text = label_positions[gene]
        ax.plot([x_mark0, x_mark1], [y, y], color="#2E3A43", lw=.55, solid_capstyle="butt", zorder=4)
        ax.plot([x_mark1, x_label-.10], [y, y_text], color="#2E3A43", lw=.55, solid_capstyle="butt", zorder=4)
        ax.text(x_label, y_text, gene, ha="left", va="center", fontsize=7.2, color=color, fontweight="bold" if gene == "OCEL1" else "normal", zorder=5)
    ax.set_xlim(-.5, heat_xlim_right)
    for spine in ax.spines.values(): spine.set_visible(False)


def draw_f3d(ax: plt.Axes) -> None:
    d = read("05_validation/selected_intersection_two_cohort_metrics.csv").sort_values("gene").reset_index(drop=True)
    columns = ["GSE56815\nAUC", "GSE2208 support\nAUC", "Expression direction\nacross cohorts", "GSE2208 support\nexpression P"]
    ax.set(xlim=(-.5, len(columns)-.5), ylim=(-.5, len(d)-.5), xticks=range(len(columns)), xticklabels=columns, yticks=range(len(d)), yticklabels=d["gene"])
    def direction(value: float) -> str:
        return "down" if value < 0 else "up"
    def ptext(value: float) -> str:
        return "P < 0.001" if value < .001 else f"P = {value:.3f}"
    for i, row in enumerate(d.itertuples()):
        highlight = row.gene == "OCEL1"
        for j in range(len(columns)):
            ax.add_patch(patches.Rectangle((j-.47, i-.42), .94, .84, facecolor="#fbede7" if highlight else "#f5f8f9", edgecolor="white"))
        values = [f"{row.train_auc:.2f}", f"{row.primary_auc:.2f}", f"{direction(row.train_delta)} in GSE56815\n{direction(row.primary_delta)} in GSE2208", ptext(row.primary_p)]
        for j, text in enumerate(values): ax.text(j, i, text, ha="center", va="center", fontsize=7.4, fontweight="bold" if highlight else "normal")
    ax.invert_yaxis()
    for spine in ax.spines.values(): spine.set_visible(False)
    ax.tick_params(length=0)


def draw_f4a(ax: plt.Axes) -> None:
    sharing = read("06_primary4_panel/OCEL1_COMPONENT_SHARING.tsv")
    summary = (sharing.groupby("gene", as_index=False)
               .agg(max_r2=("max_r2", "max"), strong_matches=("shared_r2_ge_0.8", "sum"))
               .sort_values("max_r2", ascending=False).reset_index(drop=True))
    y = np.arange(len(summary))
    rng = np.random.default_rng(20260912)
    for yi, row in enumerate(summary.itertuples()):
        points = sharing.loc[sharing["gene"] == row.gene, "max_r2"].astype(float).to_numpy()
        ax.scatter(points, yi + rng.uniform(-.13, .13, len(points)), s=16, color="#9eafb7", alpha=.74, linewidths=0)
    # Keep the summary point on the same visual scale as the component points;
    # red denotes candidates with at least one strong component match.
    summary_colors = np.where(summary["max_r2"].ge(.8), COLORS["red"], "#9eafb7")
    ax.scatter(summary["max_r2"], y, color=summary_colors, s=22, linewidths=0, zorder=3)
    for yi, row in zip(y, summary.itertuples()):
        ax.text(1.025, yi, f"max {row.max_r2:.3f}; n={int(row.strong_matches)}", va="center", fontsize=6.8)
    ax.axvline(.8, color=COLORS["red"], lw=1, ls="--")
    ax.set(xlim=(0, 1.25), yticks=y, yticklabels=summary["gene"], xlabel="Matched component $r^2$")
    ax.invert_yaxis(); clean(ax)


def draw_f4b(ax: plt.Axes) -> None:
    tracks = read("06_primary4_panel/PRIMARY4_CIS_EQTL_TRACKS.tsv")
    order = ["OCEL1", "NR2F6", "MRPL34", "BABAM1"]
    palette = {"OCEL1": COLORS["darkred"], "NR2F6": COLORS["blue"], "MRPL34": COLORS["green"], "BABAM1": COLORS["purple"]}
    fig = ax.figure
    parent = ax.get_subplotspec(); ax.remove()
    grid = parent.subgridspec(4, 1, hspace=.08)
    axes = [fig.add_subplot(grid[i, 0]) for i in range(4)]
    for current, gene in zip(axes, order):
        d = tracks[tracks["exposure"] == gene]
        bg, retained = d[~d["retained_panel"]], d[d["retained_panel"]]
        cap = max(200, int(np.ceil(np.quantile(tracks["neg_log10_p"], .985) / 10) * 10))
        current.scatter(bg["position"]/1e6, bg["neg_log10_p"].clip(upper=cap), s=5, color="#c5d0d5", alpha=.5, linewidths=0)
        current.scatter(retained["position"]/1e6, retained["neg_log10_p"].clip(upper=cap), s=9, color=palette[gene], alpha=.82, linewidths=0)
        current.text(-.075, .5, gene, transform=current.transAxes, rotation=0, va="center", ha="right", fontweight="bold", fontsize=8)
        clean(current); current.set_ylim(bottom=0)
    axes[-1].set_xlabel("Chromosome 19 position (Mb)")
    axes[0].text(.99, 1.08, f"-log10(P), display cap {cap}", transform=axes[0].transAxes, ha="right", fontsize=7, color="#59686f")
    axes[0].scatter([], [], s=8, color="#c5d0d5", label="Pre-LD eligible variants")
    axes[0].scatter([], [], s=12, color="#3e4a50", label="Retained 268-variant panel")
    axes[0].legend(frameon=False, loc="upper right", fontsize=7, ncol=1)
    label(axes[0], "b")


def draw_f4c(ax: plt.Axes) -> None:
    build = read("06_primary4_panel/BUILD_SUMMARY.tsv").iloc[0]
    d = read("06_primary4_panel/PRIMARY4_PANEL_SELECTION_MAP.tsv")
    colors = {"OCEL1": COLORS["darkred"], "NR2F6": COLORS["blue"], "MRPL34": COLORS["green"], "BABAM1": COLORS["purple"]}
    bg, retained = d[~d["retained_panel"]], d[d["retained_panel"]]
    bg_score = -np.log10(bg["min_P_all_exposures"].clip(lower=np.finfo(float).tiny))
    ax.scatter(bg["position"]/1e6, bg_score, s=7, color="#c5d0d5", alpha=.55, linewidths=0)
    for gene, block in retained.groupby("driving_exposure"):
        score = -np.log10(block["min_P_all_exposures"].clip(lower=np.finfo(float).tiny))
        ax.scatter(block["position"]/1e6, score, s=14, color=colors[gene], alpha=.88, linewidths=0, label=gene)
    ax.set(xlabel="Chromosome 19 position (Mb)", ylabel="Minimum exposure -log10(P)")
    ax.text(.01, .93, f"{int(build.exposure_eligible_ukbb_n):,}  ->  {int(build.heel_technically_usable_n):,}  ->  {int(build.maf_ge_0_01_n):,}  ->  {int(build.primary_panel_n):,}", transform=ax.transAxes, fontsize=8, fontweight="bold", color="#43545d")
    ax.legend(frameon=False, ncol=4, loc="lower center", fontsize=7)
    clean(ax)


def draw_f4d(ax: plt.Axes) -> None:
    poet = read("09_primary4_mr/PRIMARY4_POET_DIAGNOSTICS.tsv").iloc[0]
    ld = read("06_primary4_panel/PRIMARY4_LD_PLOT_SUMMARY.tsv").iloc[0]
    ax.axis("off")
    cond = ax.inset_axes([.05,.18,.42,.62]); eig = ax.inset_axes([.55,.18,.40,.62])
    for sub, raw, regularized, title, logscale in [
        (cond, poet["raw_ld_condition_number"], poet["poet_ld_condition_number"], "Condition number", True),
        (eig, poet["raw_ld_min_eigen"], poet["poet_ld_min_eigen"], "Minimum eigenvalue", False),
    ]:
        sub.plot([0,1], [raw, regularized], color="#8a9aa1", lw=1.2, zorder=1)
        sub.scatter([0,1], [raw, regularized], color=["#b9c7cd", COLORS["orange"]], s=42, zorder=2)
        sub.set(xticks=[0,1], xticklabels=["Raw", "POET"], ylabel=title)
        if logscale: sub.set_yscale("log")
        sub.text(0, raw, f"{raw:.2g}", ha="center", va="bottom", fontsize=7)
        sub.text(1, regularized, f"{regularized:.3g}", ha="center", va="bottom", fontsize=7)
        clean(sub)
    ax.text(.5,.05, f"POET regularization: K = {int(poet.poet_K)}; $\\lambda$ = {poet.poet_lambda:.3f}; maximum retained pairwise $r^2$ = {ld.maximum_retained_pairwise_r2:.4f}", transform=ax.transAxes, ha="center", fontsize=7.5, color="#4e5d64")


def draw_f5a(ax: plt.Axes) -> None:
    d = read("09_primary4_mr/PRIMARY4_CISMRBEEX_RESULT.tsv").set_index("exposure").reindex(["OCEL1","NR2F6","MRPL34","BABAM1"])
    y=np.arange(4); ax.barh(y,d["theta_pip"],color=[COLORS["red"],COLORS["blue"],COLORS["green"],COLORS["purple"]])
    ax.axvline(.2,color="#64747d",lw=.9,ls="--"); ax.text(.205,.04,"PIP retention threshold = 0.20",transform=ax.get_xaxis_transform(),fontsize=7,color="#59686f",rotation=90,va="bottom")
    ax.set(yticks=y,yticklabels=d.index,xlabel="Posterior inclusion probability")
    ax.invert_yaxis(); clean(ax)


def draw_f5b(ax: plt.Axes) -> None:
    d = read("09_primary4_mr/PRIMARY4_CISMRBEEX_RESULT.tsv").set_index("exposure").reindex(["OCEL1","NR2F6","MRPL34","BABAM1"])
    d = d[d["se"].fillna(0).gt(0)].copy()
    y = np.arange(len(d)); retained = np.ones(len(d), dtype=bool)
    ax.errorbar(d.loc[retained,"beta"],y[retained],xerr=[d.loc[retained,"beta"]-d.loc[retained,"ci_lower"],d.loc[retained,"ci_upper"]-d.loc[retained,"beta"]],fmt="o",color=COLORS["red"],capsize=3,zorder=3)
    ax.axvline(0,color="#64747d",lw=.8); ax.set(yticks=y,yticklabels=d.index,xlabel="Conditional heel eBMD association on the\nstandardized genetic-expression scale")
    ax.invert_yaxis(); clean(ax)


def draw_f5c(ax: plt.Axes) -> None:
    sens=read("10_mr_sensitivities/PRIMARY4_RXY_SPECIFICATION_SENSITIVITY.tsv")
    k12=read("10_mr_sensitivities/PRIMARY4_K12_SENSITIVITY.tsv")
    npal=read("10_mr_sensitivities/PRIMARY4_NO_PAL_SENSITIVITY.tsv")
    lookup={"RXY_IDENTITY":"Identity R_X","RXY_R2_010":"Empirical R_X, $r^2 < 0.10$ (primary)","RXY_R2_001":"Empirical R_X, $r^2 < 0.01$","RXY_ALLNULL":"Empirical all-null R_X"}
    rows=[]
    for key in ["RXY_IDENTITY","RXY_R2_010","RXY_R2_001","RXY_ALLNULL"]:
        r=sens[(sens["run"]==key)&(sens["exposure"]=="OCEL1")].iloc[0]; rows.append((lookup[key],r.beta,r.ci_lower,r.ci_upper,key=="RXY_R2_010"))
    for title, table in [("POET K = 12",k12),("No-pal reconstruction",npal)]:
        r=table[table["exposure"]=="OCEL1"].iloc[0]; rows.append((title,r.beta,r.ci_lower,r.ci_upper,False))
    y=np.arange(len(rows)); b=np.array([r[1] for r in rows]); lo=np.array([r[2] for r in rows]); hi=np.array([r[3] for r in rows])
    ax.errorbar(b,y,xerr=[b-lo,hi-b],fmt="o",color=COLORS["red"],capsize=3)
    for yi,row in enumerate(rows):
        if row[4]: ax.axhspan(yi-.38,yi+.38,color="#f8e9e6",zorder=-2)
    ax.axvline(0,color="#64747d",lw=.8); ax.set(yticks=y,yticklabels=[r[0] for r in rows],xlabel="OCEL1 conditional estimate (95% CI)")
    ax.invert_yaxis(); clean(ax)


def draw_f5d(ax: plt.Axes) -> None:
    lattice=read("10_mr_sensitivities/PRIMARY4_MODEL_LATTICE.tsv"); d=lattice[lattice["exposure"]=="OCEL1"].copy()
    order=["O","O_N","O_M","O_B","O_N_M","O_N_B","O_M_B","O_N_M_B"]
    names=["OCEL1 only","+ NR2F6","+ MRPL34","+ BABAM1","+ NR2F6 + MRPL34","+ NR2F6 + BABAM1","+ MRPL34 + BABAM1","+ NR2F6 + MRPL34 + BABAM1"]
    d["run"]=pd.Categorical(d["run"],order,ordered=True); d=d.sort_values("run"); y=np.arange(len(d))
    ax.errorbar(d["beta"],y,xerr=[d["beta"]-d["ci_lower"],d["ci_upper"]-d["beta"]],fmt="o",color=COLORS["red"],capsize=3)
    ax.axvline(0,color="#64747d",lw=.8); ax.set(yticks=y,yticklabels=names,xlabel="OCEL1 conditional estimate (95% CI)")
    ax.invert_yaxis(); clean(ax)


PANELS_DRAW = [
    ("Figure2A_lasso_coefficient_path", draw_f2a, (7, 5.6)), ("Figure2B_xgboost_importance", draw_f2b, (7, 5.6)),
    ("Figure2C_linear_svm_ranking", draw_f2c, (7, 5.6)), ("Figure2D_lasso_cv", draw_f2d, (7, 5.6)),
    ("Figure3A_three_model_overlap", draw_f3a, (7, 5.6)), ("Figure3B_discovery_volcano", draw_f3b, (7, 5.6)),
    ("Figure3C_discovery_heatmap", draw_f3c, (7, 5.6)), ("Figure3D_validation_decision", draw_f3d, (7, 5.6)),
    ("Figure4A_component_sharing", draw_f4a, (7, 5.6)), ("Figure4B_regional_cis_eqtl_tracks", draw_f4b, (8.5, 7)),
    ("Figure4C_panel_construction", draw_f4c, (7, 5.6)), ("Figure4D_ld_poet_diagnostics", draw_f4d, (7, 5.6)),
    ("Figure5A_posterior_inclusion", draw_f5a, (7, 5.6)), ("Figure5B_primary_conditional_effects", draw_f5b, (7, 5.6)),
    ("Figure5C_specification_stability", draw_f5c, (7, 5.6)), ("Figure5D_local_model_lattice", draw_f5d, (7, 5.6)),
]


def composite(stem: str, functions, figsize=(13,10), ratios=None) -> None:
    fig, axes = plt.subplots(2,2,figsize=figsize,constrained_layout=True,gridspec_kw=ratios or {})
    for ax, (letter, func) in zip(axes.ravel(), functions):
        func(ax)
        if func is not draw_f4b: label(ax, letter)
    save(fig, FIGURES, stem)


def main() -> None:
    for folder in (PANELS, FIGURES):
        folder.mkdir(parents=True, exist_ok=True)
        for suffix in ("*.png", "*.pdf"):
            for path in folder.glob(suffix):
                path.unlink()
    for stem, function, size in PANELS_DRAW:
        standalone(stem, function, size)
    composite("Figure2_machine_learning_selection", [("a",draw_f2a),("b",draw_f2b),("c",draw_f2c),("d",draw_f2d)])
    composite("Figure3_machine_learning_mapping_to_bulk_evidence", [("a",draw_f3a),("b",draw_f3b),("c",draw_f3c),("d",draw_f3d)])
    composite("Figure4_local_cis_architecture_and_instrument_construction", [("a",draw_f4a),("b",draw_f4b),("c",draw_f4c),("d",draw_f4d)], figsize=(14,12))
    composite("Figure5_primary_multivariable_cisMR_results_and_robustness", [("a",draw_f5a),("b",draw_f5b),("c",draw_f5c),("d",draw_f5d)], figsize=(14,11))
    print(f"Wrote 16 panels to {PANELS} and four composite figures to {FIGURES}")


if __name__ == "__main__":
    main()
```
