## Supplementary material for "Transcriptomic and Machine-Learning Prioritization of OCEL1 in Low Bone Mineral Density With Multivariable cis-MR": Online Resource 2: Reproducibility Materials: README.md

# Online Resource 2: Reproducibility Materials

Article: Transcriptomic and Machine-Learning Prioritization of OCEL1 in Low Bone Mineral Density With Multivariable cis-MR

Authors: Jiaheng Yao; Qiong Zhong

Corresponding author: Jiaheng Yao, School of Physical Science and Engineering, College of Life Sciences and Bioengineering, Beijing Jiaotong University, Beijing 100044, China;

This archive contains the statistical analysis code, plotting code, and compact derived result tables supporting the analyses reported in the manuscript.

## Upstream resources

GSE56815 and GSE2208 are available from NCBI GEO; blood cis-eQTL summary statistics are available from eQTLGen; heel eBMD summary statistics are available as GWAS Catalog study GCST006979; and the European-ancestry LD reference is UKBB337K. These third-party resources are not redistributed in this archive.
